# Brain Structure in Acutely Underweight and Partially Weight-Restored Individuals with Anorexia Nervosa - A Coordinated Analysis by the ENIGMA Eating Disorders Working Group

**DOI:** 10.1101/2021.10.18.21264728

**Authors:** Esther Walton, Fabio Bernardoni, Victoria-Luise Batury, Klaas Bahnsen, Sara Larivière, Giovanni Abbate-Daga, Susana Andres-Perpiña, Lasse Bang, Amanda Bischoff-Grethe, Samantha J Brooks, Iain C Campbell, Giammarco Cascino, Josefina Castro-Fornieles, Enrico Collantoni, Federico D’Agata, Brigitte Dahmen, Unna N. Danner, Angela Favaro, Jamie D Feusner, Guido KW Frank, Hans-Christoph Friederich, John L Graner, Beate Herpertz-Dahlmann, Andreas Hess, Stefanie Horndasch, Allan S Kaplan, Lisa-Katrin Kaufmann, Walter H Kaye, Sahib S Khalsa, Kevin S LaBar, Luca Lavagnino, Luisa Lazaro, Renzo Manara, Amy E Miles, Gabriella F. Milos, Alessio Maria Monteleone, Palmiero Monteleone, Benson Mwangi, Owen O’Daly, Jose Pariente, Julie Roesch, Ulrike H Schmidt, Jochen Seitz, Megan E Shott, Joe J Simon, Paul A.M. Smeets, Christian K Tamnes, Elena Tenconi, Sophia I. Thomopoulos, Annemarie A. van Elburg, Aristotle N Voineskos, Georg G von Polier, Christina E Wierenga, Nancy L Zucker, Joseph A King, Paul M. Thompson, Laura A Berner, Stefan Ehrlich

## Abstract

The pattern of structural brain abnormalities in anorexia nervosa (AN) is still not well understood. While several studies report substantial deficits in grey matter volume and cortical thickness in acutely underweight patients, others find no differences, or even increases in patients compared with healthy controls. Recent weight regain before scanning may explain some of this heterogeneity across studies. To clarify the extent, magnitude, and dependencies of grey matter changes in AN, we conducted a prospective, coordinated meta-analysis of multicenter neuroimaging data. We analyzed T_1_-weighted structural MRI scans assessed with standardized methods from 685 female AN patients and 963 female healthy controls across 22 sites worldwide. In addition to a case-control comparison, we conducted a three-group analysis comparing healthy controls to acutely underweight AN patients (n = 466), and to those in treatment and partially weight-restored (n = 251). In AN, reductions in cortical thickness, subcortical volumes, and, to a lesser extent, cortical surface area, were sizable (Cohen’s *d* up to 0.95), widespread and co-localized with hub regions. Highlighting the effects of undernutrition, these deficits associated with lower BMI in the AN sample and were less pronounced in partially weight-restored patients. Notably, the effect sizes observed for cortical thickness deficits in acute AN are the largest of any psychiatric disorder investigated in the ENIGMA consortium to date. These results confirm the importance of considering weight loss and renutrition in biomedical research on AN and underscore the importance of treatment engagement to prevent potentially long-lasting structural brain changes in this population.

## Introduction

Anorexia nervosa (AN) is an eating disorder characterized by low weight, severe restrictive eating, and a high mortality rate due to complications of starvation (1). Although the underlying mechanisms are unknown, biological underpinnings are widely recognized (2). In acutely underweight patients with AN, sulcal widening and grey matter thinning are sometimes visible on computed tomography or magnetic resonance images (MRI). However, the spatial distribution and extent (and even direction, see e.g. (3,4)) of grey matter alterations in AN varies across study samples, further complicating efforts to identify the neurobiological basis of structural neural changes in AN (5,6). Possible reasons for these heterogeneous findings include different analytic approaches (e.g. voxel-versus vertex-based morphometry), small sample sizes (typically between 20 to 40 individuals per group; (6)) and clinical heterogeneity among study participants (6). Recent work suggests that weight gain is closely linked to the normalization of grey matter reductions, and that adolescents show normalized brain structure even after partial weight restoration (7,8). Therefore, grey matter changes in AN may reflect nutritional status (as opposed to a trait-level alteration) and time (i.e., weight gain) between initiating weight-gain focused treatment and MRI scanning may substantially affect the extent and magnitude of structural brain changes.

To characterize grey matter differences in AN several metrics from T_1_-weighted MRI scans can be derived, including regional cortical thickness, cortical surface area, and grey matter volume. Although cortical thickness and surface area were reported to have distinct genetic origins and life span trajectories (9), to date, only three studies have investigated surface area in AN (4,10,11).

Over the last decade, large-scale coordinated research efforts have facilitated the investigation of structural brain abnormalities, such as cortical thickness, surface area or subcortical grey matter volume in multiple psychiatric disorders (12). Through these big international collaborations, researchers have been able to carry out prospective meta-analyses — i.e. analyses that were designed a priori using harmonized protocols and did not rely on published findings, hence reducing the risk of publication bias. By including an unprecedented number of international research sites without the need to share individual-level data, these efforts can generate more generalizable and rigorous findings compared to a single large study. For example, combining data from several thousand patients with schizophrenia and unaffected controls, two recent prospective coordinated meta-analyses reported small deficits in subcortical volumes, widespread (small to moderate) cortical thinning and equally widespread, although weaker reductions in surface area (13,14). Similar, but often smaller and more localized effects were reported for major depression, bipolar disorder, PTSD, and other psychiatric disorders (12,15).

We formed the ENIGMA Eating Disorders Working Group (http://enigma.ini.usc.edu/ongoing/enigma-eating-disorders/) to characterize brain alterations in eating disorders (e.g. AN and bulimia nervosa) using the same imaging analysis, quality control and statistical analysis methods across a large number of independently collected case-control samples across the globe. Given the aforementioned heterogeneity of AN studies and the possible significant effect of weight gain, we used two complementary approaches in the current AN study: 1) a case-control two-group comparison maximizing sample size and 2) a three-group comparison. In the latter, we subcategorized participants with AN into acutely underweight and partially weight-restored patients (i.e., patients who, at the time of their MRI scan, had already been in treatment for some time and/or gained some weight). In order to better characterize patterns of altered cortical thickness we applied multiscale neural contextualization using the ENIGMA Toolbox (16). Based on prior studies (7,17,18), we predicted that grey matter volume, cortical thickness and surface area reductions would be apparent in acute AN, but would be less pronounced in partially weight-restored AN.

## Methods

### Study samples

#### Overview

Twenty-two cohorts contributed data to the AN arm of the ENIGMA Eating Disorder Working Group. Patients in the two-group case-control comparison had to be female and meet DSM IV-TR, DSM-5, or ICD-10 criteria for AN including a body mass index (BMI, kg/m^2^) <17.5 (adults) or <10th age-adjusted BMI percentile (adolescents). Control participants were healthy females with a BMI >17.5 (adults) or >10th age-adjusted BMI percentile (and no current or lifetime diagnosis of any eating disorder). For all participants, including the subgroups described below, the following exclusion criteria were applied: current comorbid severe psychiatric disorders (such as schizophrenia/schizoaffective, bipolar disorder, substance dependence), current severe neurological disorders, significant current or chronic medical illness that can explain most of the weight loss, significant lifetime neurological illness/ accidents that may have affected the brain, preterm birth < 30 weeks, and/or developmental disorders. All participating cohorts obtained approval from local institutional review boards and ethics committees, and all study participants provided written informed consent (SM section 1.1).

To disentangle the impact of weight gain from diagnosis on brain structure, we also carried out analyses based on three groups: underweight patients acutely ill with AN (‘acAN’), partially weight-restored patients with AN (‘pwrAN’) and healthy controls (HC_3_). The inclusion of the pwrAN group allowed us to assess the effect of partial/short-term weight gain on structural brain measures. Of note, the sample size of controls in the three-group comparison (denoted as HC_3_) differed from the sample size of controls for the two-group comparison (HC_2_), as not all cohorts contributed data to the three-group analysis stream (see SM Table 1). In contrast to the ‘AN’ group from the two-group comparison, ‘acAN’ cases were defined using more stringent criteria.

Specifically, in addition to the BMI criteria detailed above, acute patients had to be either (a) within the 1st week of inpatient treatment, independent of weight changes; (b) within 2 weeks of inpatient treatment but with a weight gain of <1 kg since admission; or (c) in an outpatient/ partial hospitalization program with a weight gain of <2 kg in the last 4 weeks. The pwrAN group included patients with AN currently undergoing some form of treatment (that included weight gain as a goal) who had not yet reached their target weights. Therefore, BMI was either < 18.5 (adults) or < 10th age-adjusted percentile (adolescents). If BMI was between 18.5 - 19.5 (adults) or between the 10th and 25th age-adjusted percentile, then patients had to be engaged in regular therapy for AN, have no regular menses (unless on birth control medications) and still show significant eating disorder symptoms.

#### Case-control (two-group comparison)

We aggregated data from 22 cohorts with a combined sample size of n=685 patients with AN and n=963 HC_2_ (SM Table 1 and SM Figure 1). Sample size-weighted mean age across cohorts was 21 years (range of mean ages: 15 to 27 years). Patients with AN were significantly younger compared to HC_2_ in six of the 22 cohorts. Weighted mean BMI was 15.40 kg/m^2^ (range: 14.32 to 16.91) in AN and 21.61 kg/m^2^ in HC_2_ (range: 20.81 to 23.48). In all 22 cohorts, BMI was significantly lower among participants with AN than among controls. Age-adjusted BMI, available in 15 cohorts, was also consistently lower in AN (weighted mean group difference -2.83; range: -3.83 to -1.61) compared to HC_2_ (weighted mean -0.20; range: -0.09 to 0.61). Mean age of AN onset was 16 years (range: 13 to 18 years). Mean duration of illness was 5 years (range: 1 to 13 years).

#### Acutely ill, partially weight-restored patients and controls (three-group comparison)

In up to 12 cohorts, data from n=251 pwrAN were available and contrasted with n=874 HC_3_ and n=466 acAN (SM Table 1 and SM Figure 1). pwrAN were on average 20 years old (range: 14 to 33 years) and did not differ from acAN in age, but were younger than HC_3_ in three of the 12 cohorts. The difference in weighted mean BMI was larger between pwrAN and HC_3_ than between pwrAN and acAN (see SM Table 1 and SM section 2.1, SM Figure 2). In pwrAN, mean age of onset was 15 years (range: 13 to 16 years). Mean duration of illness was 5 years (range: 1 to 20 years).

### Image acquisition and processing

All sites processed T_1_-weighted structural brain scans using FreeSurfer (http://surfer.nmr.mgh.harvard.edu; (19)) and extracted, per hemisphere, subcortical volumes for eight regions (see SM Table 3), and cortical thickness and surface area for 34 Desikan-Killiany atlas regions (20), as well as left and right hemisphere mean thickness and total surface area (see SM Table 4 and 4). Measures for the 8 subcortical and 34 cortical regions were averaged across the left and right hemispheres. Cohort-specific details on the number of scanners, vendor, strength, sequence, acquisition parameters, and FreeSurfer version run are provided in SM Table 2. The ENIGMA protocol for quality assurance was performed at each site prior to analysis, and included visual checks of the cortical segmentations and region-by-region removal of values for segmentations found to be incorrect (http://enigma.usc.edu/protocols/imaging-protocols). Histograms of all regions’ values for each site/ sample were also produced for visual inspection.

### Statistical meta-analyses and follow up analyses

Group differences for each of the 42 regions within each sample were examined using univariate linear regression. We used R’s linear model function *lm* for the two-group contrast and the *glht* function from the multcomp R package to assess all pairwise contrasts for the three-group comparison using the ‘Tukey’ method. Bilateral ROI mean volume, mean cortical thickness or total surface area measures were predicted by group (AN vs HC_2_), controlling for linear and quadratic effects of age (and intracranial volume when the outcome was subcortical volumes; model A, SM Table 15). To further assess whether group differences in cortical thickness and surface area showed regional specificity, the analyses were repeated including global mean cortical thickness or total cortical surface area as covariates in addition to age and age^2^ (model B). To test for potential effects of (partial) weight restoration, we also included models using three groups (acAN, pwrAN and HC_3_), covarying for age and age^2^ (model C). In patients (separately in AN, acAN and pwrAN), we also analyzed effects of BMI on brain structure, correcting for linear and quadratic effects of age (model D).

Analysis of multi-scanner cohorts (n=5) included binary dummy covariates for n-1 scanners. Each site conducted analyses of their sample’s individual subject data using R code created within the ENIGMA collaboration. Per model, only individuals with complete data were analysed. Random-effects meta-analyses of the Cohen’s *d* statistics and partial correlation effect sizes for each of the 42 brain regions were performed in R (version 3.5.1) using the metafor package (version 2.1-0). Throughout the manuscript, we report FDR-corrected results separately for each modality (i.e., volume, thickness and surface area) and Bonferroni-corrected results across all brain regions (i.e., corrected for 42 brain regions; *p*=0.0012). Sub-/cortical maps were generated using models provided by ‘brainder’ (brainder.org/research/brain-for-blender). Working group members had access to the statistical results from each site.

Lastly, cortical thickness findings were contextualized across micro- and macroscales using the ENIGMA Toolbox ((16), see SM section 1.2).

## Results

### Widespread reductions in brain volumes and cortical thickness, but weaker alterations in cortical surface area of patients with AN compared to controls *(two-group comparison)*

#### Subcortical brain volumes

We observed volume alterations in all eight subcortical structures (model A; SM Table 3 and Figure 1A), with largest effects in the thalamus (Cohen’s *d* = -0.69; 95% CI: [-0.86; -0.52]). The lateral ventricles were the only structures enlarged in AN with all other areas showing lower volume in AN. Mean absolute effect size across these brain regions was *d* = 0.42 (SD = 0.15).

**Figure 1.**
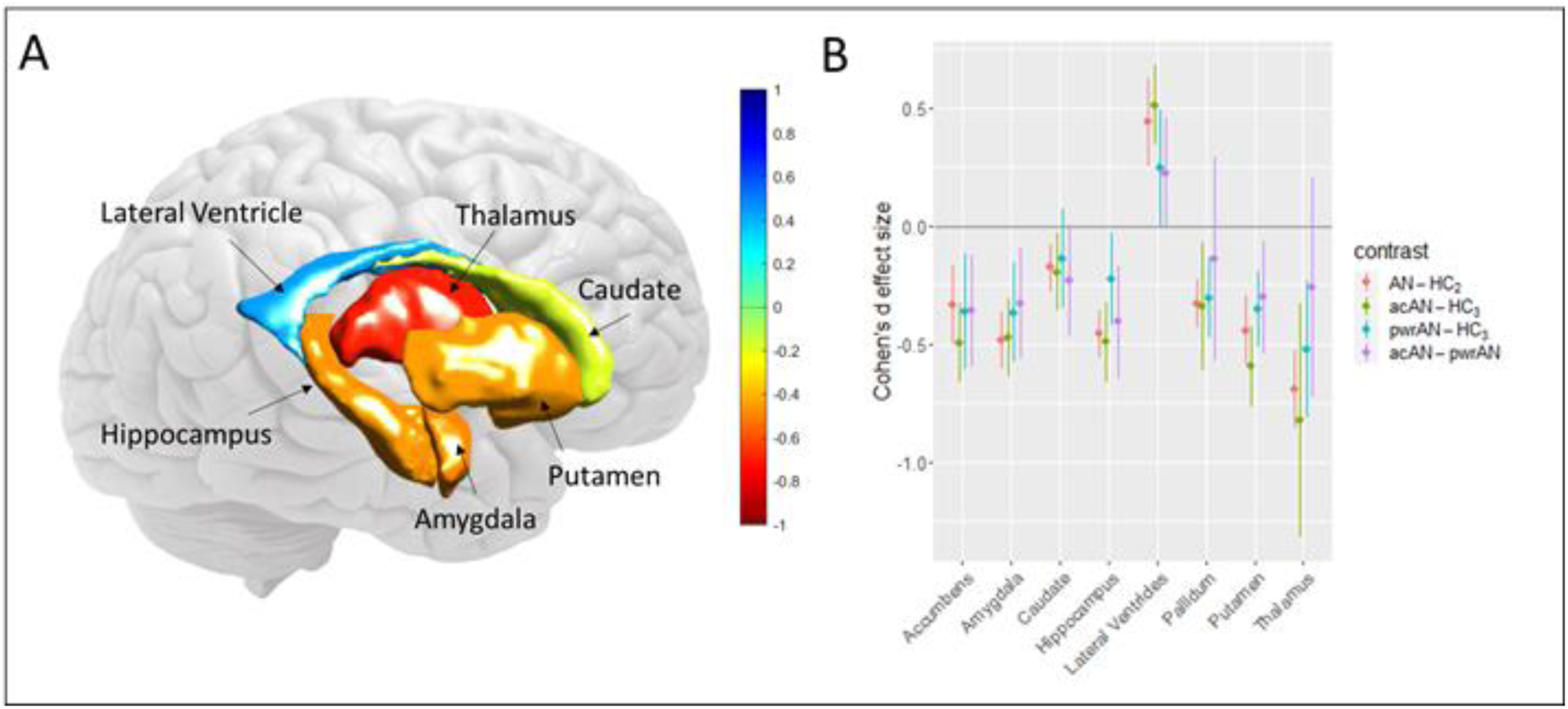
Subcortical volume reductions in anorexia nervosa. Differences (Cohen’s *d*) between A) patients with Anorexia Nervosa (AN) and healthy controls (HC_2_) and B) all groups, including acute (acAN) and partially weight-restored patients (pwrAN). Color scale: Red indicates lower volumes (Cohen’s *d*; averaged across the left and right hemispheres, but depicted on the right side of the brain) in patients compared to controls. Error bars are 95% confidence intervals.

#### Cortical thickness

We also observed widespread reductions in cortical thickness in 29 regions passing Bonferroni correction (and 30 regions passing FDR-correction; model A; SM Table 4 and Figure 2A). Largest effects were found in the superior (*d* = -0.95; 95% CI: [-1.20; -0.69]) and inferior parietal gyrus (*d* = -0.94; 95% CI: [-1.20; -0.67]). Mean effect size across these 29 regions was *d* = -0.65 (SD = 0.18). When additionally correcting for global mean thickness (model B), only 12 regions showed significant differences after Bonferroni adjustment (n=18 regions with FDR correction; SM section 2.2 and SM Table 6), suggesting that differences in region-specific cortical thickness between patients and controls were to some extent related to global thickness reductions.

**Figure 2.**
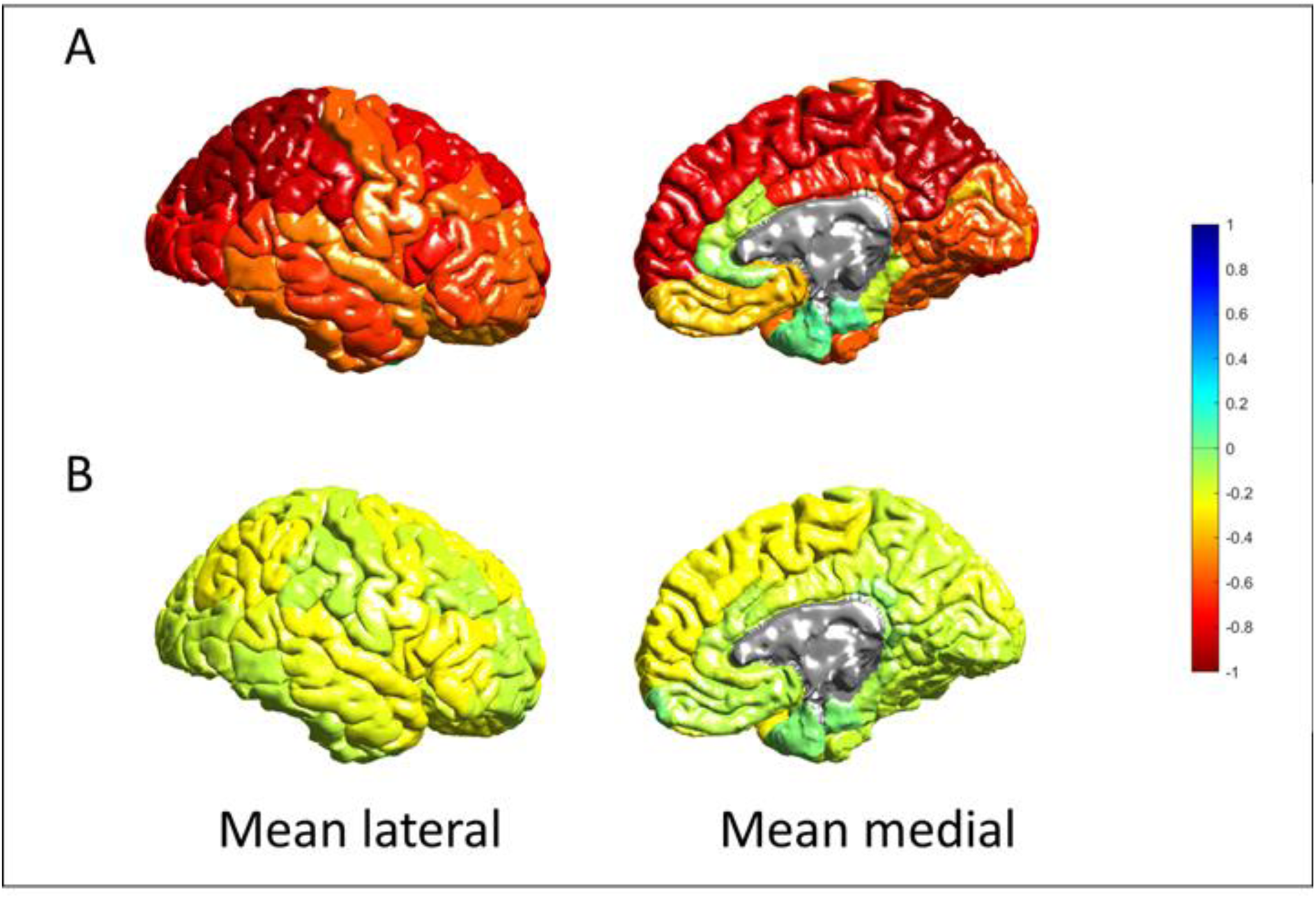
Reductions in A) cortical thickness and B) surface area between patients with Anorexia Nervosa (AN) and healthy controls (HC_2_). Color scale: Warmer colors indicate reductions (Cohen’s *d* effect size; averaged across the left and right hemispheres, but depicted on the right side of the brain) in patients compared to controls.

#### Cortical surface area

We also observed reductions in cortical surface area in 16 regions passing Bonferroni correction (n=16 with FDR-correction; model A; SM Table 5 and Figure 2B). Largest effects were found in the transverse temporal gyrus (d = -0.29; 95% CI: [-0.42; -0.15]) and pars opercularis (d = - 0.28; 95% CI: [-0.38; -0.17]). Mean effect size across these 16 regions was d = -0.23; roughly a third of that observed for reductions in cortical thickness and half of that found for volumetric reductions. When additionally correcting for global mean surface area (model B), only the paracentral and transverse temporal gyrus showed a significant difference (SM section 2.2 and SM Table 7), suggesting that differences in region-specific cortical surface area between AN and HC_2_ were to a large extent driven by global reductions in surface area.

### Reductions in volume, thickness and surface area are less severe in partially weight-restored patients than in acutely ill patients *(three-group comparison)*

#### Subcortical brain volumes

Compared to the volumetric differences between acAN and HC_3_ (mean d_acAN-HC_ = 0.49; SD = 0.18), the differences between pwrAN and HC_3_ were reduced by 36% (mean d_pwrAN-HC_ = 0.31; SD = 0.12), suggesting that volume reductions in pwrAN were smaller than in acAN (model C). pwrAN also had larger subcortical volumes than acAN (mean d_acAN-pwrAN_ = 0.28; SD = 0.09; Figure 1B, SM section 2.3 and SM Table 8). Overall, these findings suggest that reductions in subcortical volumes in pwrAN were smaller than those observed between acAN and HC_3_.

#### Cortical thickness

While reductions in thickness in acAN remained significant in only 11 of the original 29 cortical regions found significant in the AN-HC_2_ comparison (likely because of lower power in the smaller sample), effect sizes correlated strongly between these analyses (model C; SM section 2.3). Compared to the thickness reductions in acAN (d_acAN-HC_ = 0.67; SD = 0.15), differences between pwrAN and HC_3_ were reduced by 36% (d_pwrAN-HC_ = 0.43; SD = 0.17; Figure 3A). Cortical thickness in pwrAN was also larger than in acAN (d_acAN-pwrAN_ = 0.49; SD = 0.14). This suggests again that cortical thickness reductions in pwrAN were less severe than in acAN compared to HC_3_ (i.e., indicating partial normalization of thickness during weight restoration). The reductions appeared to be driven to a large extent by global reductions in cortical thickness. Once controlled for global thickness, effects were reduced by 75% (acAN-HC_3_), 63% (pwrAN-HC_3_) and 86% (acAN-pwrAN; SM Table 9).

**Figure 3.**
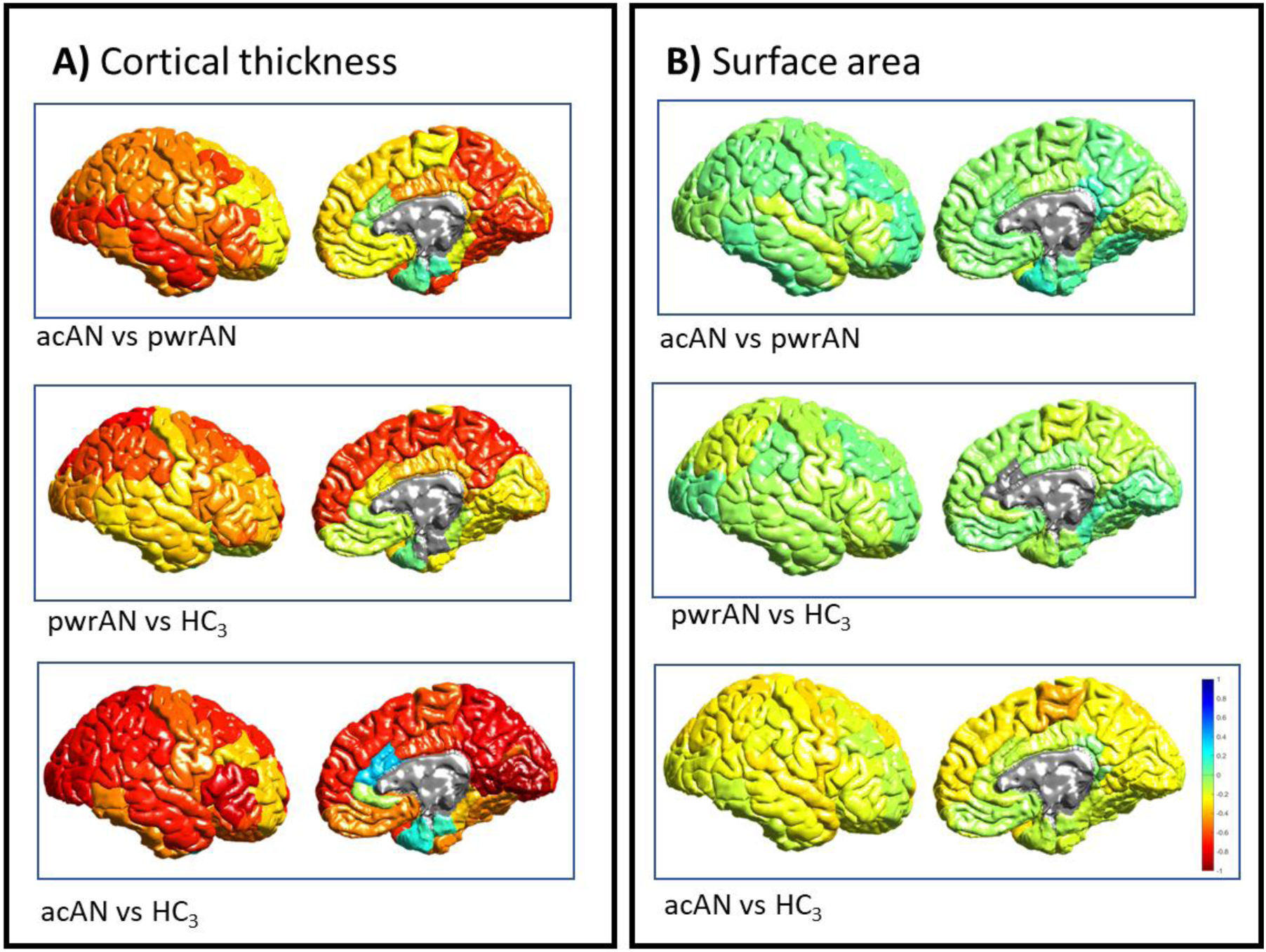
Pairwise reductions, shown as Cohen’s d effect sizes, in A) cortical thickness and B) surface area between acute patients with Anorexia Nervosa (acAN), partially weight-restored patients (pwrAN) and healthy controls (HC_3_). Color scale: Red indicates reductions (Cohen’s d; averaged across the left and right hemispheres, but depicted on the right side of the brain).

#### Cortical surface area

Differences in surface area in seven of the original 16 cortical regions found significant in the AN-HC_2_ comparison, remained significant in acAN with a very good agreement in point estimates across analyses, but a weaker correlation of effect sizes compared to subcortical volume and cortical thickness (model C, SM section 2.3). Effect sizes for reductions in surface area were on average 52% smaller contrasting pwrAN to HC_3_ (d_pwrAN-HC_ = 0.10; SD = 0.05) compared to reductions observed in acAN (d_AN-HC_ = 0.26; SD = 0.07; Figure 3B). This suggests again that cortical surface area reductions in pwrAN were less severe than in acAN compared to HC_3_ (i.e. indicating partial normalization). These reductions seem to be driven to a large extent by global reductions in cortical area. Controlling for global surface area reduced effects sizes from d = 0.26 to d = 0.08 for acAN-HC_3_, from d = 0.10 to 0.05 for pwrAN-HC_3_ and increased only very slightly from d = 0.07 to 0.08 for acAN-pwrAN (SM Table 10).

#### Multiscale neural contextualization

Having established robust patterns of atrophy, we evaluated whether these abnormalities were associated with micro- and macroscale brain organization (see SM section 1.1). Patterns of cortical thickness reductions in AN corresponded to regions with greater, and more evenly distributed (across the layers), cellular densities ((21); Figure 4A), particularly converging in parietal and frontal cytoarchitectonic classes ((22); Figure 4B). Leveraging connectivity data from the Human Connectome Project (23), AN-related atrophy implicated functional and structural cortico-cortical hub regions more strongly than nonhub (i.e., locally connected) regions (Figure 4C and 4D).

**Figure 4.**
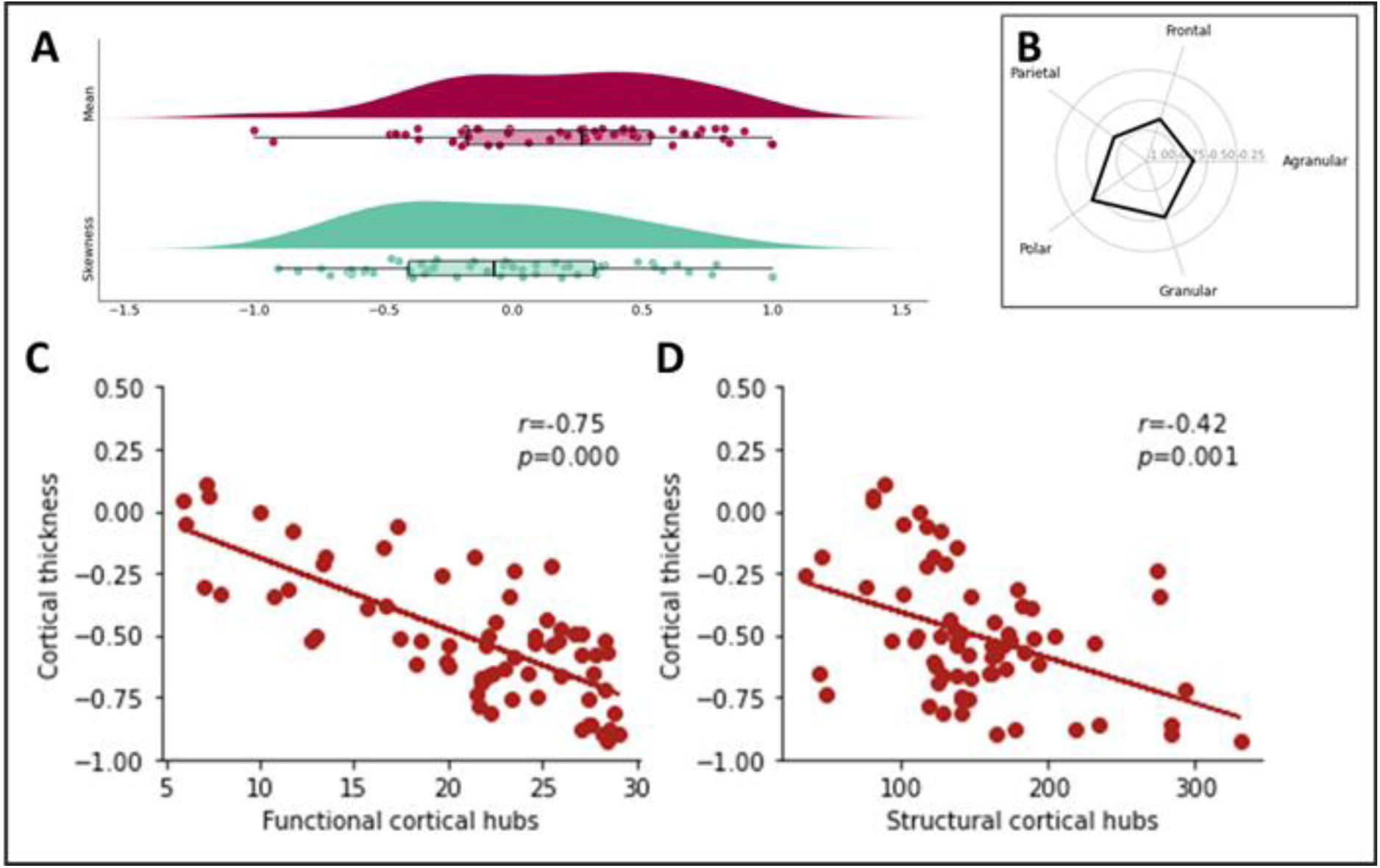
Neural contextualization of cortical thickness case-control differences. Cohen’s d effect sizes in the context of: A) regional cytoarchitecture, specifically overall cellular density (A; top panel) and laminar differentiation (A; lower panel); B) cytoarchitectonic classes based on postmortem work by von Economo and Koskinas; and C) functional as well as D) structural connectivity hubs.

### Reductions in grey matter volume and thickness are associated with BMI

In patients with AN, BMI was positively associated with volumes in the thalamus, putamen, amygdala and hippocampus after Bonferroni-correction (as well as the accumbens and pallidum after FDR-correction; model D; SM Table 11). The mean effect was r = 0.20 (SD = 0.03) with largest associations in the amygdala (r = 0.23; 95% CI [0.12; 0.35]).

Compared to the volumetric findings, associations between BMI and cortical thickness were larger with a mean effect of r = 0.32 (SD = 0.06), significant across 25 regions (n=28 after FDR correction; SM Table 12).

For surface area, associations with BMI were the weakest (r = 0.18; SD = 0.04) with only 5 significant regions (n=7 after FDR; SM Table 13). Together, these findings suggest that cortical thickness (and subcortical volumes and surface area, albeit to a lesser extent) in AN might be related to BMI, and therefore weight status.

Effects were similar – and in the case of volume and cortical thickness slightly stronger – when using an age-adjusted measure of BMI (SM Tables 11-13).

### Moderator effects of additional variables

The two-group differences in volume, thickness and surface area remained stable when also covarying for the proportion of antidepressant or antipsychotic medication use, AN subtype (restrictive or binge-purge), depressive symptoms, duration of illness or tesla field strength of the MRI scanner (SM File 2). Furthermore, almost none of these clinical or technical variables showed significant moderating effects after FDR correction (SM File 2 and SM Table 14). Samples with a larger proportion of patients with a restrictive subtype were characterized by reduced thickness in the insula and reduced volume in the putamen and nucleus accumbens. These subcortical volumes were also larger for samples with a larger proportion of patients with a binge-purge subtype.

## Discussion

In this prospective coordinated meta-analysis combining scans from 685 patients with AN from around the world (total sample size n=1,648, including controls), we found widespread and sizable reductions in cortical thickness and subcortical brain volume in the underweight state of AN as compared to HC. Surface area was also reduced but the effect sizes were smaller. Comparison of patients acutely ill with AN, partially weight-restored patients with AN, and healthy controls indicated a substantial positive effect of partial weight gain on all three structural brain metrics. This represents the largest structural neuroimaging study in AN to date. Taken together, results suggest that AN is associated with global reductions in grey matter (and no increases) and that these reductions might be highly state-dependent, i.e. related to lower BMI.

In line with some, but not all, previous (smaller) studies (3–5,7,10,17), reductions in cortical thickness and subcortical volume in AN were on average moderate (mean Cohen’s d of 0.65 and 0.42, respectively), and between two to four times larger than in other psychiatric disorders that are often comorbid with AN, including depression and obsessive-compulsive disorder (effect sizes between 0.10 to 0.31; (12), see also SM Figure 3). In fact, until now the largest effects among all ENIGMA studies in psychiatric disorders (apart from the 22q11 deletion syndrome, which is characterized by significant hypertrophy) have been found in schizophrenia with Cohen’s d effect sizes ranging between 0.12 to 0.46 for subcortical structures and up to 0.53 for cortical thickness (12–14). Although smaller than the effects observed in neurodegenerative diseases such as Alzheimer’s disease (24), the effects found in AN in the current study are higher than those found in schizophrenia, and can therefore be considered the largest among all psychiatric disorders. Embedding our findings within a multiscale framework (25) revealed that patterns of cortical thickness reductions primarily affected regions with greater cellular densities as well as densely connected hub regions. In line with previous psychiatric and neurological disorders (26,27), the high metabolic demands and increased connectional flow of hub regions may account for their selective vulnerability in the manifestation of AN-related atrophy.

Unlike our findings of larger alterations in volume and cortical thickness compared to other disorders, surface area reductions were similar in size (mean Cohen’s d, 0.23) compared to those observed in other psychiatric disorders such as OCD or schizophrenia (Cohen’s d between 0.16 to 0.33) and slightly smaller than those reported for depression (0.26 to 0.57; (12)). Even though effects for surface area were smaller, they followed a similar pattern as for cortical thickness.

In line with this and previous studies (28), BMI showed small to moderate associations with subcortical volumes and cortical thickness (and to a smaller degree with surface area). The moderating effect of AN subtype may also be related to this, since patients with a restrictive subtype are often characterized by more rapid and extensive weight loss (29–31). Interestingly, abnormally high body weight has also been also associated with lower grey matter and bariatric surgery seems to reverse some of these effects (32). Underlining the importance of state effects such as weight loss and subsequent weight gain, our three-group comparison showed that partial weight recovery was associated with an attenuated reduction in all three grey matter metrics seen in acutely underweight AN (36-52% smaller differences compared to patients that were at the very beginning of treatment). Reversibility of “pseudoatrophy” in AN, i.e. increases in grey matter volume and cortical thickness (and even gyrification) following weight restoration, has been reported in previous cross-sectional structural neuroimaging investigations of long-term weight recovered former AN patients (10,17,33– 35) and a small number of longitudinal studies (7,36–38). However, based on a recent study comparing different age groups, normalization seems to be easier to achieve in younger patients (18). Importantly, however, the current findings go beyond previous studies by supporting these effects across many cohorts in a coordinated and harmonized meta-analytic research design. Overall, these findings highlight the need to control for the clinical state (acute vs. already gaining weight) of the disorder when studying AN, i.e., the drastic impact of AN on the brain is strongly related to undernutrition and therefore rapidly changes with weight gain or treatment.

Results should be interpreted in the light of the following limitations: first, based on the neuroimaging method employed, microstructural changes cannot be detected. Therefore, our study cannot exclude the persistence of irreversible scars after weight restoration at the microstructural level. This is important, as recent studies have shown elevated neuronal and glial damage markers in AN (39,40). Second, we aggregated data from different study sites, but differences in MRI scanners and acquisition protocols can introduce non-biological variations (41). However, we did covary for potential scanner differences (within each study site) and found little evidence for moderating effects of scanner across study sites. Third, we did not assess or control for the impact of comorbidities (e.g. OCD, depression), but prior studies indicated generally smaller effects of these psychiatric disorders on brain structure (15,42). Hence, it is unlikely that our findings were better accounted for by comorbid conditions. Fourth, our study included healthy controls with a BMI as low as 17.5. It is possible that these individuals also showed some subthreshold eating disorder symptoms and were therefore more similar to pwrAN than to the control group. However, our analysis indicated that BMI had a similar association with brain structure as diagnostic group, suggesting that biases due to misclassification were unlikely. Last, we assume that differences on T_1_- weighted MRI measurements relate to true variations in brain morphology rather than measurement errors or artifacts.

In summary, based on the largest and most representative sample to date, the current results indicate that acutely underweight individuals with AN have sizable and widespread reductions of subcortical volumes and cortical thickness and, to a lesser extent, cortical surface area. Effect sizes for reductions in cortical thickness are the largest detected to date among all psychiatric disorders (12). These effects are attenuated in partially weight restored patients and all metrics of structural brain changes (especially cortical and subcortical grey matter) associate with current BMI, which mirrors the clinical state of AN. Our findings underline the importance of taking weight loss and renutrition into consideration in biomedical research on AN and the importance of effective early intervention and treatment engagement to prevent long-lasting structural brain changes in AN.

## Supporting information

Supplemental Materials File 2 (Moderator analyses)

STROBE checklist

## Data Availability

All data produced in the present study are available upon reasonable request to the authors.

## Acknowledgements

This work was supported by the Carus Promotionskolleg (KB), the Ministerio de Igualdad, Spain (grant number 234/09) and by the Generalitat de Catalunya (2009 SGR 1119) (SA), NIH R21MH86017, NIH R01MH113588 (ABG), Biomedical Research Centre (BRC) UK (ICC), the Alicia Koplowitz Foundation (FAK) (DN040546) and by the Generalitat de Catalunya 2017SGR4881 (JCF), German Ministry for Education and Research (grants 01GV0602 and 01GV0623) (BD), NIMH R01MH105662, NIMH R01MH093535 (JDF), NIMH K23MH080135, R01MH096777 (GKWF), NIH RC1MH088678 (JLG), German Ministry for Education and Research (grants 01GV0602 and 01GV0623) (BHD), the CAMH AFP Innovation Fund (ASK), Swiss Anorexia Nervosa Foundation (project no. 19-12), the Palatin Foundation, and the Gottfried and Julia Bangerter-Rhyner-Foundation (LKK), NIH R01MH042984-17A1, Price Foundation, NIH R01MH113588 (WHK), NIMH K23MH112949 (SSK), NIH RC1MH088678 (KSL), the Carlos III Research Institute of the Spanish Ministry of Health, FIS PI040829 and by the Generalitat de Catalunya (2009 SGR 1119) (LL), the CAMH AFP Innovation Fund (AEM), Swiss Anorexia Nervosa Foundation (project no. 19-12), the Palatin Foundation, and the Gottfried and Julia Bangerter-Rhyner-Foundation (GFM), Biomedical Research Centre (BRC) UK (OOD), Biomedical Research Centre (BRC) UK (UHS), German Ministry for Education and Research (grants 01GV0602 and 01GV0623) (JS), NIMH K23MH080135, R01MH096777 (MES), DFG: SI 2087/2-1, BR 4852/1-1, Swiss Anorexia Nervosa Foundation: 57-16 (JJS), Research Council of Norway (#288083, #223273); South-Eastern Norway Regional Health Authority (#2019069, #2021070, #500189) (CKT), the CAMH AFP Innovation Fund (ANV), German Ministry for Education and Research (grants 01GV0602 and 01GV0623) (GGvP), NIH R21MH86017, R01MH113588 (CEW), NIH RC1MH088678 (NLZ), NIH RC1MH088678 (JAK), a National Institute of Health Research (NIHR) Senior Investigator Award (US), the NIHR Mental Health Biomedical Research Centre at the South London and Maudsley NHS Foundation Trust and King’s College London (ICC, US and OOD), K23MH118418; NARSAD Young Investigator Grant from the Brain & Behavior Research Foundation (LAB), and SFB 940, DFG: EH 367/5-1, EH 367/7-1 and the Swiss Anorexia Nervosa Foundation (SE). This work is further supported by the European Union’s Horizon 2020 research and innovation programme (EarlyCause, grant n° 848158, to EW). The ENIGMA Working Group acknowledges the NIH Big Data to Knowledge (BD2K) award for foundational support and consortium development (U54 EB020403 to Paul M. Thompson).

## Data availability

All data produced in the present study are available upon reasonable request to the authors.

