## Supplemental Materials for "Brain Structure in Acutely Underweight and Partially Weight-Restored Individuals with Anorexia Nervosa - A Coordinated Analysis by the ENIGMA Eating Disorders Working Group"

**Supplementary Materials**

SM 1 Methods

SM 1.1 Ethical approval

All participating cohorts obtained approval from local institutional review boards and ethics committees, and all study participants provided written informed consent. AN_BDD_UCLA and AN_Reward_UCLA: All procedures performed in studies involving human participants were in accordance with the ethical standards of the institutional and/or national research committee and with the 1964 Helsinki declaration and its later amendments or comparable ethical standards. The UCLA Institutional Review Board (IRB) approved the study (UCLA IRB#10-001767). AN_Oslo: This study is approved by the Norwegian Regional Committee for Medical and Health Research Ethics. Written informed consent was obtained from all participants (2015/793). Denver: The study was approved by the Colorado Multiple Institutional Review Board. We obtained written assent from the participant and consent from the legal guardian prior to conducting any of the study procedures. Erlangen: The Ethics Committee of the University Hospital of Erlangen gave approval for the study and it was conducted in accordance with the Declaration of Helsinki (220_15B). Heidelberg: The medical ethics committee of the Medical Faculty Heidelberg at the Ruprecht-Karls-University in Heidelberg, Germany, approved this study, and written informed consent was obtained from all participants (protocol numbers S-373/2014, S-592/2015, S-125/201). IDIBAPS: The ethics committee of clinical research of the Hospital Clínic approved the study protocol. All subjects gave written informed consent after a detailed description of the study. The investigation was carried out in accordance with the latest version of the Declaration of Helsinki (Seoul, Republic of Korea, October 2008); (HCB/2016/0061). LIBR_AN: This study was approved by the Western Institutional Review Board, and all methods were carried out in accordance with relevant guidelines and regulations. Prior to the experiment, each participant provided written informed consent, and informed consent was obtained from a parent and/or legal guardian for the participants under 18 years of age (Trial Registration: ClinicalTrials.gov #NCT03758326). Napoli_Salerno: The study was approved by the Ethics Committee of the University of Campania ‘Luigi Vanvitelli’ and performed in accordance with the ethical standards laid down in the 1964 Declaration of Helsinki and its later amendments (454/2013). Padova: Ethical permission was obtained from the ethics committee of the Padova Hospital. After completely describing the study to the subjects, written informed consent was obtained (ID 1598P). Torino: The study was approved by the local Ethics Committee [Comitato Etico Interaziendale A.O.U. Città della Salute e della Scienza di Torino—A.O. Ordine Mauriziano—A.S.L. Città di Torino; approval #12042010]. UCSC_AdolAN and UCSC_sMRI: The study was conducted according to the IRB regulations of the University of California, San Diego Human Research Protections Program (090008). Zurich: The study was approved by the local ethics committee and the study protocol complied with the Declaration of Helsinki. All participants gave their written informed consent prior to study enrollment (KEK-ZH-Nr. 2013-0273). BRCACE and PREDICTA: The study received ethical approval from the London—City Road and Hampstead Research Ethics Committee and the King's College London Psychiatry, Nursing and Midwifery Research Ethics Subcommittee (References: 15/LO/0196 and HR-15/16-2836).

SM 1.2 Microstructural and macrostructural contextualization

Microstructural and macrostructural contextualization of cortical thickness was carried out using the ENIGMA Toolbox, as described in detail in Larivière et al. (1) and briefly summarized here.

Neural microstructure: Regional cytoarchitecture was characterized using statistical moments of staining profiles, based on data from The BigBrain Project, an ultra-high-resolution 3D reconstruction of a sliced and stained human brain (2) from a 65-year-old male.

For the ENIGMA Toolbox, the highest resolution full brain volume was used (100μm isotropic voxels) to generate 50 equivolumetric surfaces between the pial and white matter surfaces. Next, staining intensity profiles, representing neuronal density and soma size by cortical depth, were sampled along 327,684 surface points in the direction of cortical columns. Vertex-wise cytoarchitecture was characterized by taking two central moments of the staining intensity profiles (mean and skewness). The Desikan-Killiany atlas was then nonlinearly transformed to the BigBrain histological surfaces and central moments were averaged within each parcels, excluding outlier vertices with values more than three scaled median absolute deviations away from the parcel median.

Mean intracortical staining across the mantle allows inferences on overall cellular density, whereas analysis of profile skewness indexes the distribution of cells across upper and lower layers of the cortex—a critical dimension of laminar differentiation.

The cytoarchitectonic atlas of von Economo and Koskinas (3,4) was mapped to cortical surface templates. Cytoarchitectonic class labels from the original five different structural types of cerebral cortex (agranular, frontal, parietal, polar, granular) were manually assigned to each parcellation region and subsequently mapped to vertex-wise space. To stratify cortex-wide effects according to the five cytoarchitectonic classes, parcellated data (e.g., disease-related atrophy map on the Desikan-Killiany atlas) was mapped to vertexwise space and values were iteratively averaged from all vertices within each class.

Neural macrostructure: connectivity data were selected from a group of unrelated healthy adults (n=207; 83 males, mean age±SD=28.73±3.73 years, range=22-36 years) from the Human Connectome Project (HCP) dataset (5). High-resolution resting-state functional and diffusion MRI structural data were parcellated according to the Desikan-Killiany atlas. Normative functional connectivity matrices were generated by computing pairwise correlations between the time series of all cortical regions; negative connections were set to zero. Subject-specific connectivity matrices were then z-transformed and aggregated across participants to construct a group-average functional connectome. Normative structural connectivity matrices were generated from preprocessed diffusion MRI data. Weighted degree centrality maps were derived from functional (or structural) connectivity data by computing the sum of all weighted cortico-cortical connections for every region, with higher degree centrality denoting hub regions.

Spatial permutation tests

The significance of the spatial correlation in two given brain maps may be inflated due to the intrinsic spatial smoothness. We therefore used spin permutation tests to assess statistical significance of these spatial correlations. Within this framework, null models of overlap between cortical maps are generated by projecting the spatial coordinates of cortical data onto the surface spheres, applying randomly sampled rotations (10,000 repetitions), and reassigning connectivity values. The empirical (i.e., original) correlation coefficients are then compared against the null distributions determined by the ensemble of correlation coefficients comparing spatially permuted cortical maps.

SM 2 Results

SM 2.1 Sample setup

For our main two-group case-control comparison, we aggregated data from 22 cohorts with a combined sample size of n=685 patients with AN and n=963 healthy controls. We also carried out three-group comparisons using data of n=251 partially weight-restored patients, n=874 healthy controls and n=466 acutely ill patients, with at least two of these groups available in up to 12 cohorts (SM Figure 1).

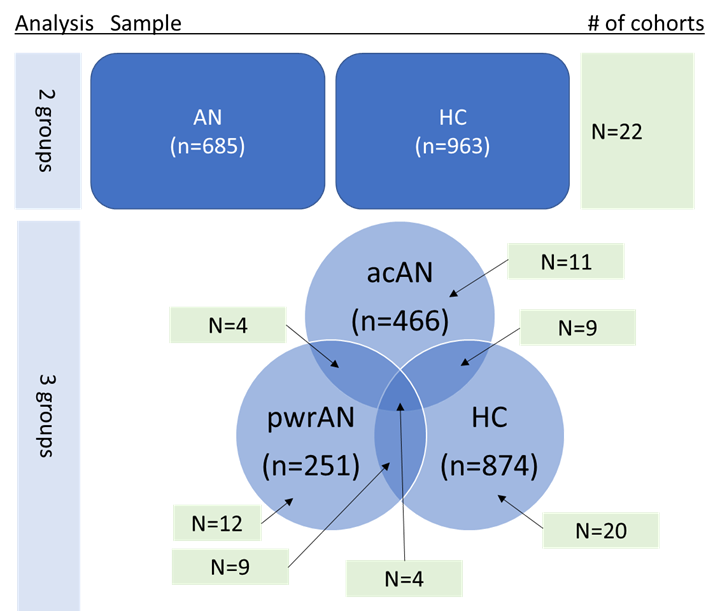

SM Figure 1. Sample setup.

Weighted mean BMI was 16.58 kg/m^2^ in partially weight-restored patients (range: 14.32 to 18.33) compared to 15.36 kg/m^2^ (range: 12.82 to 15.91) in acutely ill patients and 21.57 kg/m^2^ in controls (range: 20.81 to 23.48). While all pairwise contrasts in BMI between the three groups were statistically significant in all cohorts, overall the difference between controls and partially weight-restored patients was larger (beta=4.91; 95% CI: [4.06; 5.75]) than the difference between acutely ill and partially weight-recovered patients (beta=-2.60; 95% CI: [-3.36; -1.83]). Results were similar when using BMI-SDS (SM Table 1 and SM Figure 2).

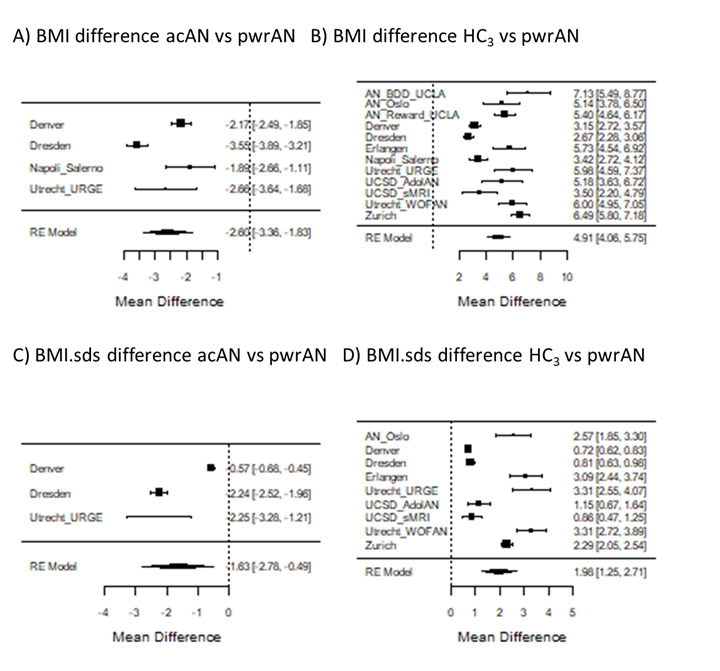

SM Figure 2. Pairwise contrasts in BMI and age-adjusted BMI (BMI.sds; standard deviation scores).

SM 2.2 AN patients and healthy controls (two-group comparison)

Cortical thickness

When additionally correcting for global mean thickness (model B), only 12 regions showed significant differences after Bonferroni adjustment (n=18 regions at FDR correction). The direction of effect reversed for two of these regions (fusiform and lateral orbitofrontal; both with significant differences before and after global thickness correction; SM Table 4 and SM Table 6), suggesting that these regions were less severely affected in patients compared to controls, but only after accounting for global thinning. Across all 12 regions, effect sizes were on average reduced by 53% (mean absolute d_n=12_ before global thickness correction = 0.60 versus mean absolute d_n=12_ after correction = 0.28). This suggests that differences in region-specific cortical thickness between patients and controls were to some extent driven by global thickness reductions.

Surface area

When additionally correcting for global mean surface area (model B), only the paracentral and transverse temporal gyrus showed significant differences after Bonferroni adjustment (n=4 regions at FDR correction; SM Table 7). These findings suggest that differences in region-specific cortical surface area between patients and controls were to a large extent driven by global reductions in surface area.

SM 2.3. Acutely ill, partially weight-recovered patients and controls (three group comparison)

Subcortical volume

Contrasting controls to acutely ill AN patients (i.e. using a more stringent ‘case’ definition here referred to as ‘acAN’, see methods) in a subset of five cohorts, we were able to confirm our original findings, which were based on a more inclusive ‘case’ definition (here, referred to as ‘AN’). While only six of the original eight subcortical regions found significant in the AN-HC_2_ comparison remained significant, effect sizes correlated strongly between these analyses (rho=0.99; p=5.72*10-6). Larger confidence intervals and a smaller sample size (SM Table 1, SM Figure 1, SM Table 8 and Figure 1B) in the acAN analysis together with a strong correlation in effect sizes and agreement in point estimates across analyses (mean d_AN-HC_ = 0.42; SD = 0.15 versus mean d_acAN-HC_ = 0.49; SD = 0.18) suggest that a reduction in the number of significant findings were largely driven by a lack of power rather than a lack of differences between acAN and HC.

Cortical thickness

While only 11 of the original 29 cortical regions found significant in the AN-HC_2_ comparison remained significant based on a more stringent acAN case definition (SM Table 4 and SM Table 9), effect sizes correlated strongly between these analyses (rho=0.85; p=1.16*10^-10^) with an additional agreement in point estimates across analyses (mean d_AN-HC_ = 0.66; SD = 0.17 versus mean d_acAN-HC_ = 0.67; SD = 0.15). This suggests – as before – that a reduction in the number of significant findings was largely driven by a lack of power rather than a lack of differences between acAN and HC_3_.

Surface area

Seven of the original 16 cortical regions found significant in the AN-HC_2_ comparison remained significant based on a more stringent acAN case definition (SM Table 5 and SM Table 10). Although effect sizes between these analyses correlated less strongly than for the two other metrics (rho=0.71; p=2.16*10^-6^), agreement in point estimates across analyses was very good (mean d_AN-HC_ = 0.23; SD = 0.04 versus mean d_acAN-HC_ = 0.26; SD = 0.07). This suggests that patient groups based on either definition showed very similar patterns in surface area reductions when compared to controls.

SM 2.4 Cortical thickness reductions across four psychiatric disorders

Reductions in cortical thickness in AN compared to those observed in other psychiatric disorders, such as schizophrenia (6) and disorders often comorbid with AN, including depression (7) and obsessive-compulsive disorder (8).

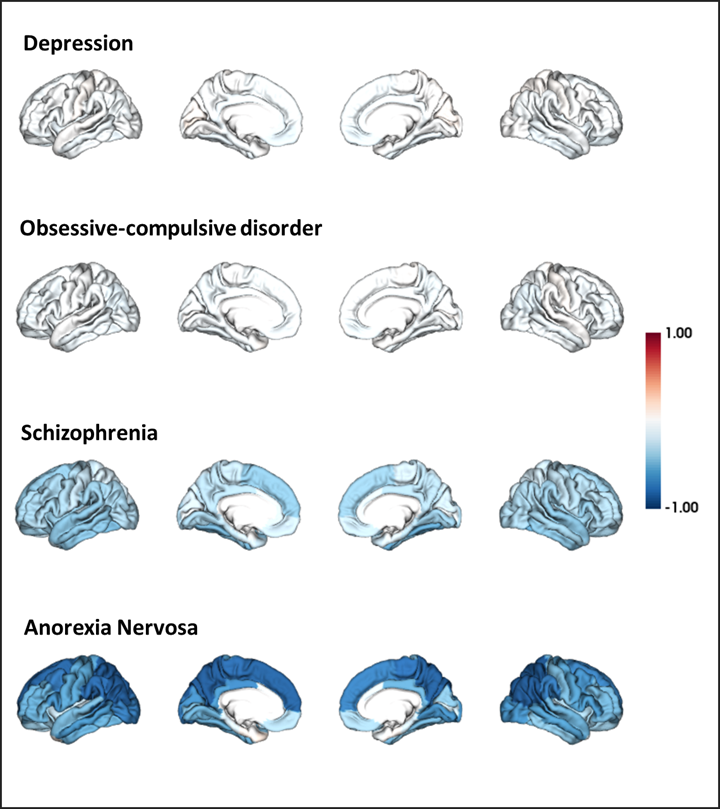

SM Figure 3. Cortical thickness reductions (uncorrected), shown as Cohen’s d effect sizes, across four psychiatric disorders. Images were created using the ENIGMA Toolbox (Larivière et al. (1)).
