## Supplemental Materials File 2 (Moderator analyses) for "Brain Structure in Acutely Underweight and Partially Weight-Restored Individuals with Anorexia Nervosa - A Coordinated Analysis by the ENIGMA Eating Disorders Working Group"

Overview moderator* analyses

**Page 2: Subcortical**

Two-group case-control differences in a meta-regression with medication, AN subtype, depressive symptoms, duration of illness or tesla strength as an additional moderator

**Page 3: Subcortical**

Effect of the moderator

**Page 4-8: Surface area**

Two-group case-control differences in a meta-regression with medication, AN subtype, depressive symptoms, duration of illness or tesla strength as an additional moderator

**Page 9-13: Surface area**

Effect of the moderator

**Page 14-18: Cortical thickness**

Two-group case-control differences in a meta-regression with medication, AN subtype, depressive symptoms, duration of illness or tesla strength as an additional moderator

**Page 19-23: Cortical thickness**

Effect of the moderator

* medication defined as the proportion of patients on antidepressive or antipsychotic medication;

Subtype = restrictive or binge-purge subtype;

Depressive symptoms as a continuous variable;

Duration of illness in years;

Tesla field strength either 3 or 1.5 tesla.

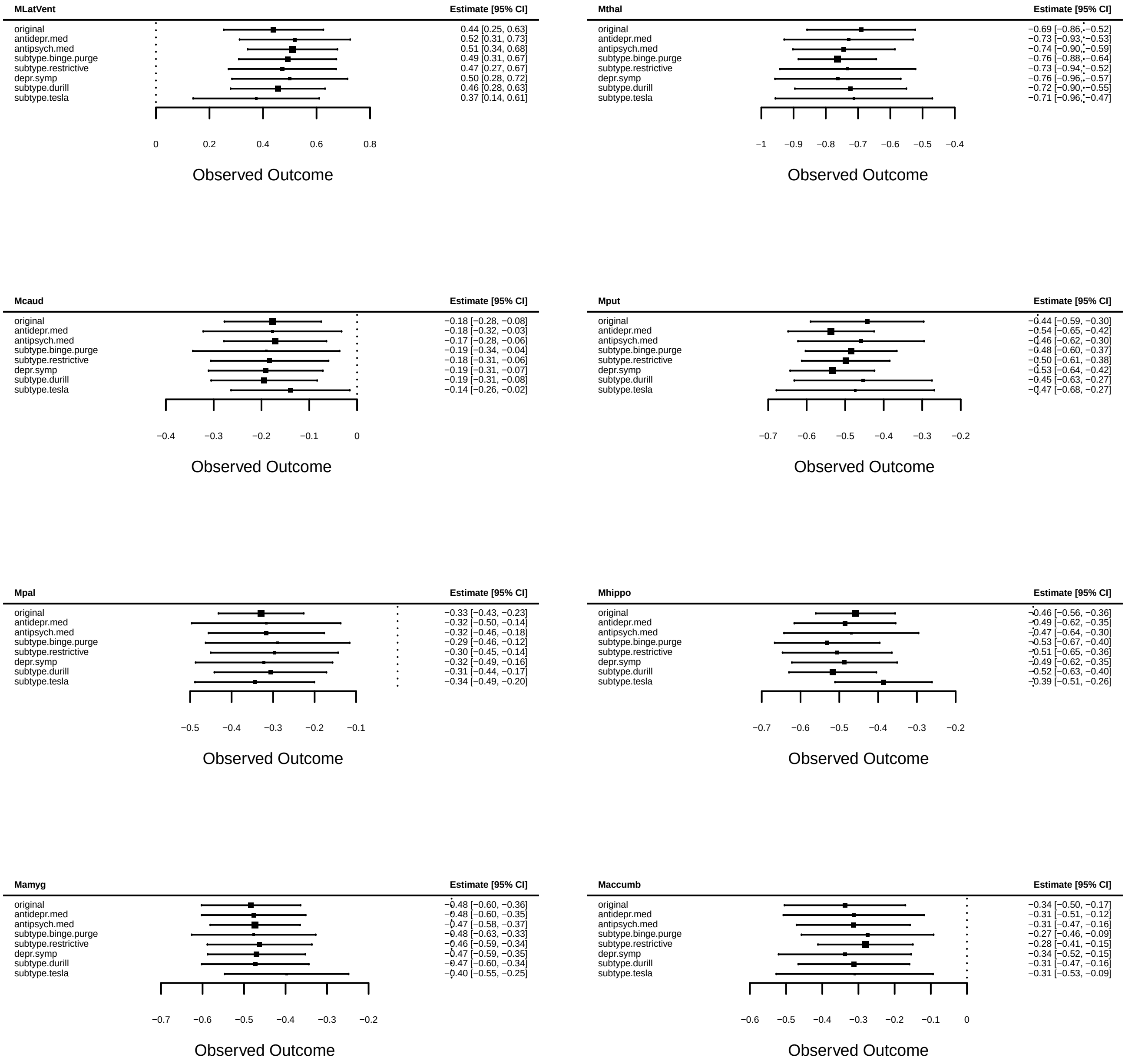
MLatVent
Estimate [95% CI]
original 0.44 [0.25, 0.63]
antidepr.med 0.52 [0.31, 0.73]
antipsych.med 0.51 [0.34, 0.68]
subtype.binge.purge 0.49 [0.31, 0.67]
subtype.restrictive 0.47 [0.27, 0.67]
depr.symp 0.50 [0.28, 0.72]
subtype.durill 0.46 [0.28, 0.63]
subtype.tesla 0.37 [0.14, 0.61]
0 0.2 0.4 0.6 0.8
Observed Outcome
Mthal
Estimate [95% CI]
original −0.69 [−0.86, −0.52]
antidepr.med −0.73 [−0.93, −0.53]
antipsych.med −0.74 [−0.90, −0.59]
subtype.binge.purge −0.76 [−0.88, −0.64]
subtype.restrictive −0.73 [−0.94, −0.52]
depr.symp −0.76 [−0.96, −0.57]
subtype.durill −0.72 [−0.90, −0.55]
subtype.tesla −0.71 [−0.96, −0.47]
−1 −0.9 −0.8 −0.7 −0.6 −0.5 −0.4
Observed Outcome
Mcaud
Estimate [95% CI]
original −0.18 [−0.28, −0.08]
antidepr.med −0.18 [−0.32, −0.03]
antipsych.med −0.17 [−0.28, −0.06]
subtype.binge.purge −0.19 [−0.34, −0.04]
subtype.restrictive −0.18 [−0.31, −0.06]
depr.symp −0.19 [−0.31, −0.07]
subtype.durill −0.19 [−0.31, −0.08]
subtype.tesla −0.14 [−0.26, −0.02]
−0.4 −0.3 −0.2 −0.1 0
Observed Outcome
Mput
Estimate [95% CI]
original −0.44 [−0.59, −0.30]
antidepr.med −0.54 [−0.65, −0.42]
antipsych.med −0.46 [−0.62, −0.30]
subtype.binge.purge −0.48 [−0.60, −0.37]
subtype.restrictive −0.50 [−0.61, −0.38]
depr.symp −0.53 [−0.64, −0.42]
subtype.durill −0.45 [−0.63, −0.27]
subtype.tesla −0.47 [−0.68, −0.27]
−0.7 −0.6 −0.5 −0.4 −0.3 −0.2
Observed Outcome
Mpal
Estimate [95% CI]
original −0.33 [−0.43, −0.23]
antidepr.med −0.32 [−0.50, −0.14]
antipsych.med −0.32 [−0.46, −0.18]
subtype.binge.purge −0.29 [−0.46, −0.12]
subtype.restrictive −0.30 [−0.45, −0.14]
depr.symp −0.32 [−0.49, −0.16]
subtype.durill −0.31 [−0.44, −0.17]
subtype.tesla −0.34 [−0.49, −0.20]
−0.5 −0.4 −0.3 −0.2 −0.1
Observed Outcome
Mhippo
Estimate [95% CI]
original −0.46 [−0.56, −0.36]
antidepr.med −0.49 [−0.62, −0.35]
antipsych.med −0.47 [−0.64, −0.30]
subtype.binge.purge −0.53 [−0.67, −0.40]
subtype.restrictive −0.51 [−0.65, −0.36]
depr.symp −0.49 [−0.62, −0.35]
subtype.durill −0.52 [−0.63, −0.40]
subtype.tesla −0.39 [−0.51, −0.26]
−0.7 −0.6 −0.5 −0.4 −0.3 −0.2
Observed Outcome
Mamyg
Estimate [95% CI]
original −0.48 [−0.60, −0.36]
antidepr.med −0.48 [−0.60, −0.35]
antipsych.med −0.47 [−0.58, −0.37]
subtype.binge.purge −0.48 [−0.63, −0.33]
subtype.restrictive −0.46 [−0.59, −0.34]
depr.symp −0.47 [−0.59, −0.35]
subtype.durill −0.47 [−0.60, −0.34]
subtype.tesla −0.40 [−0.55, −0.25]
−0.7 −0.6 −0.5 −0.4 −0.3 −0.2
Observed Outcome
Maccumb
Estimate [95% CI]
original −0.34 [−0.50, −0.17]
antidepr.med −0.31 [−0.51, −0.12]
antipsych.med −0.31 [−0.47, −0.16]
subtype.binge.purge −0.27 [−0.46, −0.09]
subtype.restrictive −0.28 [−0.41, −0.15]
depr.symp −0.34 [−0.52, −0.15]
subtype.durill −0.31 [−0.47, −0.16]
subtype.tesla −0.31 [−0.53, −0.09]
−0.6 −0.5 −0.4 −0.3 −0.2 −0.1 0
Observed Outcome

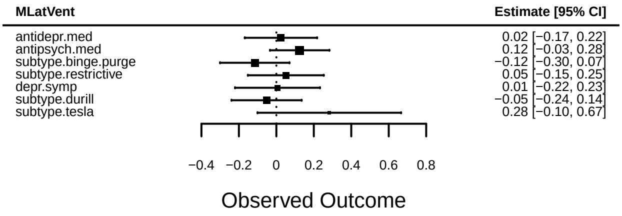
MLatVent
Estimate [95% CI]
antidepr.med 0.02 [−0.17, 0.22]
antipsych.med 0.12 [−0.03, 0.28]
subtype.binge.purge −0.12 [−0.30, 0.07]
subtype.restrictive 0.05 [−0.15, 0.25]
depr.symp 0.01 [−0.22, 0.23]
subtype.durill −0.05 [−0.24, 0.14]
subtype.tesla 0.28 [−0.10, 0.67]
−0.4 −0.2 0 0.2 0.4 0.6 0.8
Observed Outcome

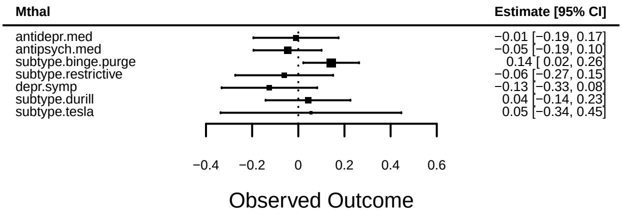
Mthal
Estimate [95% CI]
antidepr.med −0.01 [−0.19, 0.17]
antipsych.med −0.05 [−0.19, 0.10]
subtype.binge.purge 0.14 [ 0.02, 0.26]
subtype.restrictive −0.06 [−0.27, 0.15]
depr.symp −0.13 [−0.33, 0.08]
subtype.durill 0.04 [−0.14, 0.23]
subtype.tesla 0.05 [−0.34, 0.45]
−0.4 −0.2 0 0.2 0.4 0.6
Observed Outcome

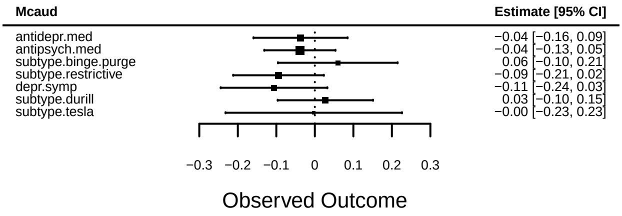
Mcaud
Estimate [95% CI]
antidepr.med −0.04 [−0.16, 0.09]
antipsych.med −0.04 [−0.13, 0.05]
subtype.binge.purge 0.06 [−0.10, 0.21]
subtype.restrictive −0.09 [−0.21, 0.02]
depr.symp −0.11 [−0.24, 0.03]
subtype.durill 0.03 [−0.10, 0.15]
subtype.tesla −0.00 [−0.23, 0.23]
−0.3 −0.2 −0.1 0 0.1 0.2 0.3
Observed Outcome

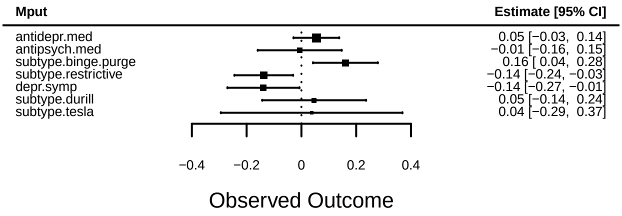
Mput
Estimate [95% CI]
antidepr.med 0.05 [−0.03, 0.14]
antipsych.med −0.01 [−0.16, 0.15]
subtype.binge.purge 0.16 [ 0.04, 0.28]
subtype.restrictive −0.14 [−0.24, −0.03]
depr.symp −0.14 [−0.27, −0.01]
subtype.durill 0.05 [−0.14, 0.24]
subtype.tesla 0.04 [−0.29, 0.37]
−0.4 −0.2 0 0.2 0.4
Observed Outcome

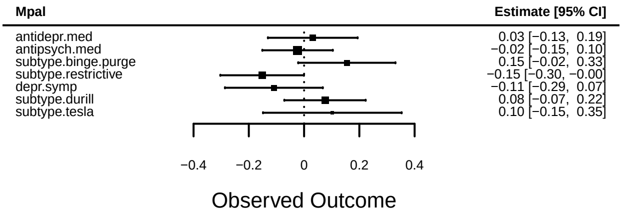
Mpal
Estimate [95% CI]
antidepr.med 0.03 [−0.13, 0.19]
antipsych.med −0.02 [−0.15, 0.10]
subtype.binge.purge 0.15 [−0.02, 0.33]
subtype.restrictive −0.15 [−0.30, −0.00]
depr.symp −0.11 [−0.29, 0.07]
subtype.durill 0.08 [−0.07, 0.22]
subtype.tesla 0.10 [−0.15, 0.35]
−0.4 −0.2 0 0.2 0.4
Observed Outcome

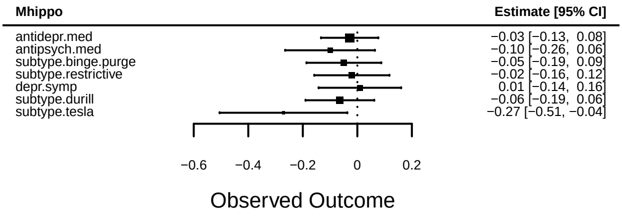
Mhippo
Estimate [95% CI]
antidepr.med −0.03 [−0.13, 0.08]
antipsych.med −0.10 [−0.26, 0.06]
subtype.binge.purge −0.05 [−0.19, 0.09]
subtype.restrictive −0.02 [−0.16, 0.12]
depr.symp 0.01 [−0.14, 0.16]
subtype.durill −0.06 [−0.19, 0.06]
subtype.tesla −0.27 [−0.51, −0.04]
−0.6 −0.4 −0.2 0 0.2
Observed Outcome

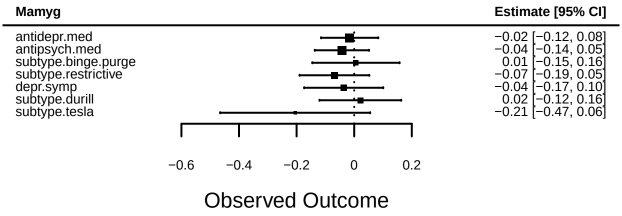
Mamyg
Estimate [95% CI]
antidepr.med −0.02 [−0.12, 0.08]
antipsych.med −0.04 [−0.14, 0.05]
subtype.binge.purge 0.01 [−0.15, 0.16]
subtype.restrictive −0.07 [−0.19, 0.05]
depr.symp −0.04 [−0.17, 0.10]
subtype.durill 0.02 [−0.12, 0.16]
subtype.tesla −0.21 [−0.47, 0.06]
−0.6 −0.4 −0.2 0 0.2
Observed Outcome

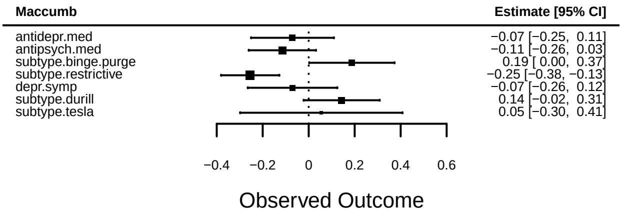
Maccumb
Estimate [95% CI]
antidepr.med −0.07 [−0.25, 0.11]
antipsych.med −0.11 [−0.26, 0.03]
subtype.binge.purge 0.19 [ 0.00, 0.37]
subtype.restrictive −0.25 [−0.38, −0.13]
depr.symp −0.07 [−0.26, 0.12]
subtype.durill 0.14 [−0.02, 0.31]
subtype.tesla 0.05 [−0.30, 0.41]
−0.4 −0.2 0 0.2 0.4 0.6
Observed Outcome

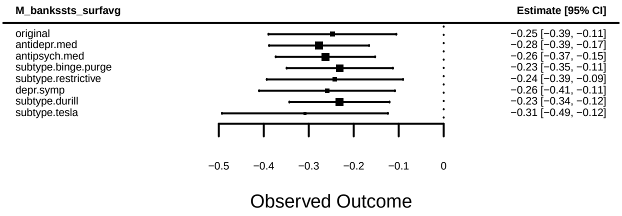
M_bankssts_surfavg
Estimate [95% CI]
original −0.25 [−0.39, −0.11]
antidepr.med −0.28 [−0.39, −0.17]
antipsych.med −0.26 [−0.37, −0.15]
subtype.binge.purge −0.23 [−0.35, −0.11]
subtype.restrictive −0.24 [−0.39, −0.09]
depr.symp −0.26 [−0.41, −0.11]
subtype.durill −0.23 [−0.34, −0.12]
subtype.tesla −0.31 [−0.49, −0.12]
−0.5 −0.4 −0.3 −0.2 −0.1 0
Observed Outcome

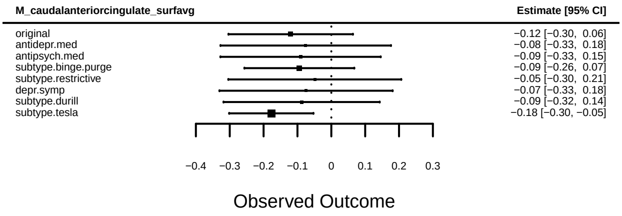
M_caudalanteriorcingulate_surfavg
Estimate [95% CI]
original −0.12 [−0.30, 0.06]
antidepr.med −0.08 [−0.33, 0.18]
antipsych.med −0.09 [−0.33, 0.15]
subtype.binge.purge −0.09 [−0.26, 0.07]
subtype.restrictive −0.05 [−0.30, 0.21]
depr.symp −0.07 [−0.33, 0.18]
subtype.durill −0.09 [−0.32, 0.14]
subtype.tesla −0.18 [−0.30, −0.05]
−0.4 −0.3 −0.2 −0.1 0 0.1 0.2 0.3
Observed Outcome

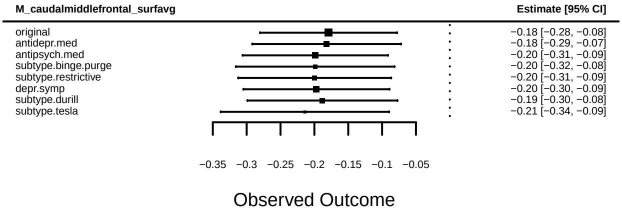
M_caudalmiddlefrontal_surfavg
Estimate [95% CI]
original −0.18 [−0.28, −0.08]
antidepr.med −0.18 [−0.29, −0.07]
antipsych.med −0.20 [−0.31, −0.09]
subtype.binge.purge −0.20 [−0.32, −0.08]
subtype.restrictive −0.20 [−0.31, −0.09]
depr.symp −0.20 [−0.30, −0.09]
subtype.durill −0.19 [−0.30, −0.08]
subtype.tesla −0.21 [−0.34, −0.09]
−0.35 −0.3 −0.25 −0.2 −0.15 −0.1 −0.05
Observed Outcome

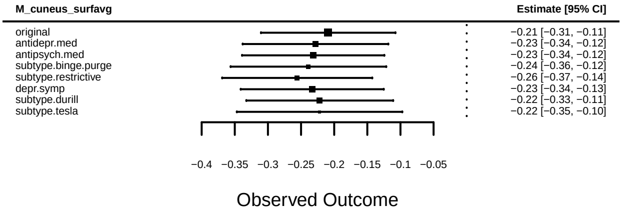
M_cuneus_surfavg
Estimate [95% CI]
original −0.21 [−0.31, −0.11]
antidepr.med −0.23 [−0.34, −0.12]
antipsych.med −0.23 [−0.34, −0.12]
subtype.binge.purge −0.24 [−0.36, −0.12]
subtype.restrictive −0.26 [−0.37, −0.14]
depr.symp −0.23 [−0.34, −0.13]
subtype.durill −0.22 [−0.33, −0.11]
subtype.tesla −0.22 [−0.35, −0.10]
−0.4 −0.35 −0.3 −0.25 −0.2 −0.15 −0.1 −0.05
Observed Outcome

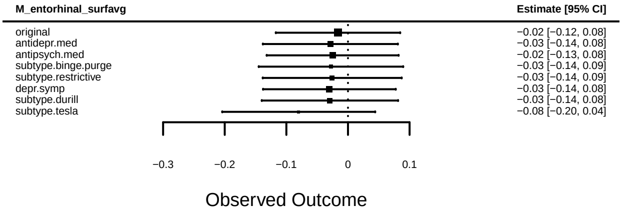
M_entorhinal_surfavg
Estimate [95% CI]
original −0.02 [−0.12, 0.08]
antidepr.med −0.03 [−0.14, 0.08]
antipsych.med −0.02 [−0.13, 0.08]
subtype.binge.purge −0.03 [−0.14, 0.09]
subtype.restrictive −0.03 [−0.14, 0.09]
depr.symp −0.03 [−0.14, 0.08]
subtype.durill −0.03 [−0.14, 0.08]
subtype.tesla −0.08 [−0.20, 0.04]
−0.3 −0.2 −0.1 0 0.1
Observed Outcome

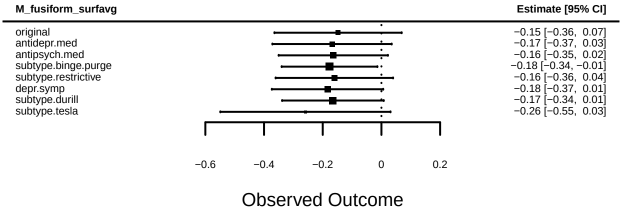
M_fusiform_surfavg
Estimate [95% CI]
original −0.15 [−0.36, 0.07]
antidepr.med −0.17 [−0.37, 0.03]
antipsych.med −0.16 [−0.35, 0.02]
subtype.binge.purge −0.18 [−0.34, −0.01]
subtype.restrictive −0.16 [−0.36, 0.04]
depr.symp −0.18 [−0.37, 0.01]
subtype.durill −0.17 [−0.34, 0.01]
subtype.tesla −0.26 [−0.55, 0.03]
−0.6 −0.4 −0.2 0 0.2
Observed Outcome

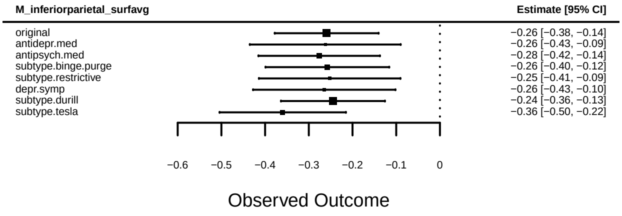
M_inferiorparietal_surfavg
Estimate [95% CI]
original −0.26 [−0.38, −0.14]
antidepr.med −0.26 [−0.43, −0.09]
antipsych.med −0.28 [−0.42, −0.14]
subtype.binge.purge −0.26 [−0.40, −0.12]
subtype.restrictive −0.25 [−0.41, −0.09]
depr.symp −0.26 [−0.43, −0.10]
subtype.durill −0.24 [−0.36, −0.13]
subtype.tesla −0.36 [−0.50, −0.22]
−0.6 −0.5 −0.4 −0.3 −0.2 −0.1 0
Observed Outcome

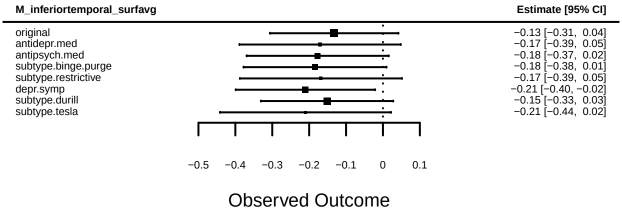
M_inferiortemporal_surfavg
Estimate [95% CI]
original −0.13 [−0.31, 0.04]
antidepr.med −0.17 [−0.39, 0.05]
antipsych.med −0.18 [−0.37, 0.02]
subtype.binge.purge −0.18 [−0.38, 0.01]
subtype.restrictive −0.17 [−0.39, 0.05]
depr.symp −0.21 [−0.40, −0.02]
subtype.durill −0.15 [−0.33, 0.03]
subtype.tesla −0.21 [−0.44, 0.02]
−0.5 −0.4 −0.3 −0.2 −0.1 0 0.1
Observed Outcome

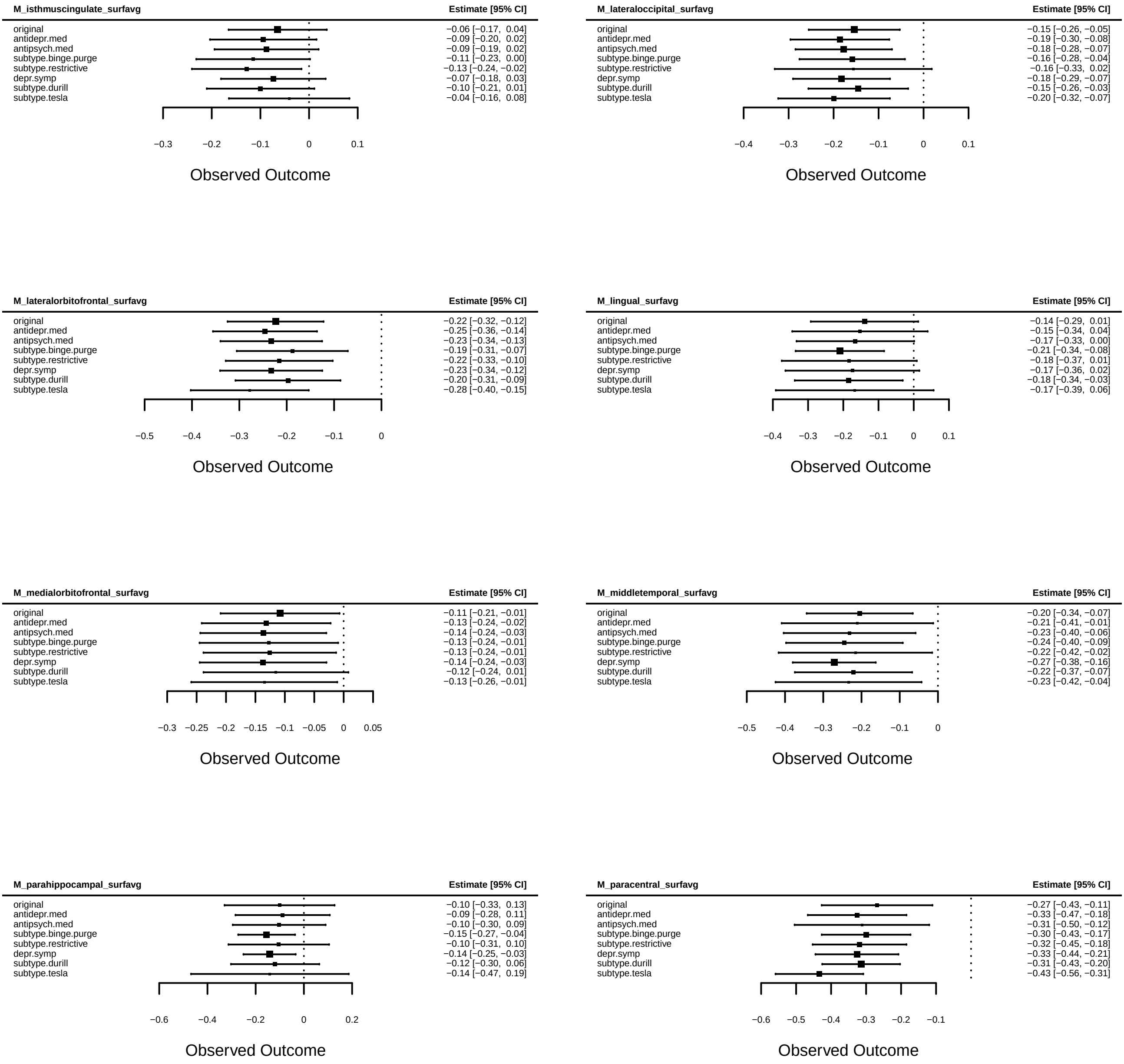
M_isthmuscingulate_surfavg
Estimate [95% CI]
original −0.06 [−0.17, 0.04]
antidepr.med −0.09 [−0.20, 0.02]
antipsych.med −0.09 [−0.19, 0.02]
subtype.binge.purge −0.11 [−0.23, 0.00]
subtype.restrictive −0.13 [−0.24, −0.02]
depr.symp −0.07 [−0.18, 0.03]
subtype.durill −0.10 [−0.21, 0.01]
subtype.tesla −0.04 [−0.16, 0.08]
−0.3 −0.2 −0.1 0 0.1
Observed Outcome
M_lateraloccipital_surfavg
Estimate [95% CI]
original −0.15 [−0.26, −0.05]
antidepr.med −0.19 [−0.30, −0.08]
antipsych.med −0.18 [−0.28, −0.07]
subtype.binge.purge −0.16 [−0.28, −0.04]
subtype.restrictive −0.16 [−0.33, 0.02]
depr.symp −0.18 [−0.29, −0.07]
subtype.durill −0.15 [−0.26, −0.03]
subtype.tesla −0.20 [−0.32, −0.07]
−0.4 −0.3 −0.2 −0.1 0 0.1
Observed Outcome
M_lateralorbitofrontal_surfavg
Estimate [95% CI]
original −0.22 [−0.32, −0.12]
antidepr.med −0.25 [−0.36, −0.14]
antipsych.med −0.23 [−0.34, −0.13]
subtype.binge.purge −0.19 [−0.31, −0.07]
subtype.restrictive −0.22 [−0.33, −0.10]
depr.symp −0.23 [−0.34, −0.12]
subtype.durill −0.20 [−0.31, −0.09]
subtype.tesla −0.28 [−0.40, −0.15]
−0.5 −0.4 −0.3 −0.2 −0.1 0
Observed Outcome
M_lingual_surfavg
Estimate [95% CI]
original −0.14 [−0.29, 0.01]
antidepr.med −0.15 [−0.34, 0.04]
antipsych.med −0.17 [−0.33, 0.00]
subtype.binge.purge −0.21 [−0.34, −0.08]
subtype.restrictive −0.18 [−0.37, 0.01]
depr.symp −0.17 [−0.36, 0.02]
subtype.durill −0.18 [−0.34, −0.03]
subtype.tesla −0.17 [−0.39, 0.06]
−0.4 −0.3 −0.2 −0.1 0 0.1
Observed Outcome
M_medialorbitofrontal_surfavg
Estimate [95% CI]
original −0.11 [−0.21, −0.01]
antidepr.med −0.13 [−0.24, −0.02]
antipsych.med −0.14 [−0.24, −0.03]
subtype.binge.purge −0.13 [−0.24, −0.01]
subtype.restrictive −0.13 [−0.24, −0.01]
depr.symp −0.14 [−0.24, −0.03]
subtype.durill −0.12 [−0.24, 0.01]
subtype.tesla −0.13 [−0.26, −0.01]
−0.3 −0.25 −0.2 −0.15 −0.1 −0.05 0 0.05
Observed Outcome
M_middletemporal_surfavg
Estimate [95% CI]
original −0.20 [−0.34, −0.07]
antidepr.med −0.21 [−0.41, −0.01]
antipsych.med −0.23 [−0.40, −0.06]
subtype.binge.purge −0.24 [−0.40, −0.09]
subtype.restrictive −0.22 [−0.42, −0.02]
depr.symp −0.27 [−0.38, −0.16]
subtype.durill −0.22 [−0.37, −0.07]
subtype.tesla −0.23 [−0.42, −0.04]
−0.5 −0.4 −0.3 −0.2 −0.1 0
Observed Outcome
M_parahippocampal_surfavg
Estimate [95% CI]
original −0.10 [−0.33, 0.13]
antidepr.med −0.09 [−0.28, 0.11]
antipsych.med −0.10 [−0.30, 0.09]
subtype.binge.purge −0.15 [−0.27, −0.04]
subtype.restrictive −0.10 [−0.31, 0.10]
depr.symp −0.14 [−0.25, −0.03]
subtype.durill −0.12 [−0.30, 0.06]
subtype.tesla −0.14 [−0.47, 0.19]
−0.6 −0.4 −0.2 0 0.2
Observed Outcome
M_paracentral_surfavg
Estimate [95% CI]
original −0.27 [−0.43, −0.11]
antidepr.med −0.33 [−0.47, −0.18]
antipsych.med −0.31 [−0.50, −0.12]
subtype.binge.purge −0.30 [−0.43, −0.17]
subtype.restrictive −0.32 [−0.45, −0.18]
depr.symp −0.33 [−0.44, −0.21]
subtype.durill −0.31 [−0.43, −0.20]
subtype.tesla −0.43 [−0.56, −0.31]
−0.6 −0.5 −0.4 −0.3 −0.2 −0.1
Observed Outcome

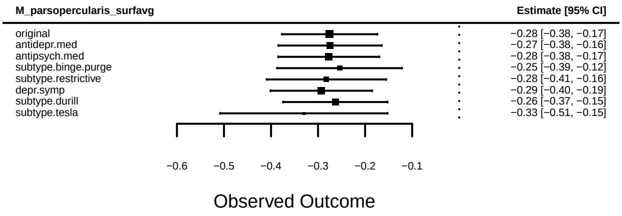
M_parsopercularis_surfavg
Estimate [95% CI]
original −0.28 [−0.38, −0.17]
antidepr.med −0.27 [−0.38, −0.16]
antipsych.med −0.28 [−0.38, −0.17]
subtype.binge.purge −0.25 [−0.39, −0.12]
subtype.restrictive −0.28 [−0.41, −0.16]
depr.symp −0.29 [−0.40, −0.19]
subtype.durill −0.26 [−0.37, −0.15]
subtype.tesla −0.33 [−0.51, −0.15]
−0.6 −0.5 −0.4 −0.3 −0.2 −0.1
Observed Outcome

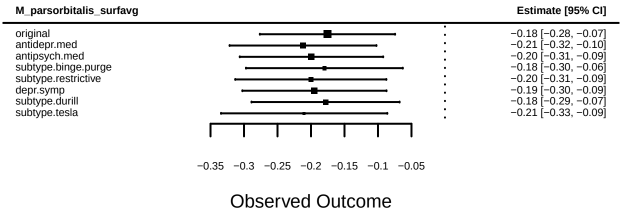
M_parsorbitalis_surfavg
Estimate [95% CI]
original −0.18 [−0.28, −0.07]
antidepr.med −0.21 [−0.32, −0.10]
antipsych.med −0.20 [−0.31, −0.09]
subtype.binge.purge −0.18 [−0.30, −0.06]
subtype.restrictive −0.20 [−0.31, −0.09]
depr.symp −0.19 [−0.30, −0.09]
subtype.durill −0.18 [−0.29, −0.07]
subtype.tesla −0.21 [−0.33, −0.09]
−0.35 −0.3 −0.25 −0.2 −0.15 −0.1 −0.05
Observed Outcome

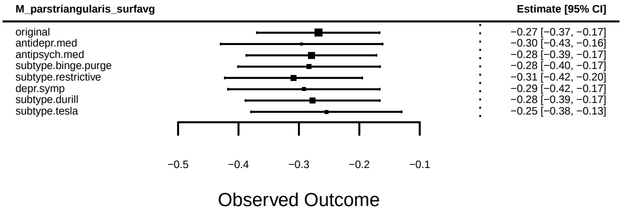
M_parstriangularis_surfavg
Estimate [95% CI]
original −0.27 [−0.37, −0.17]
antidepr.med −0.30 [−0.43, −0.16]
antipsych.med −0.28 [−0.39, −0.17]
subtype.binge.purge −0.28 [−0.40, −0.17]
subtype.restrictive −0.31 [−0.42, −0.20]
depr.symp −0.29 [−0.42, −0.17]
subtype.durill −0.28 [−0.39, −0.17]
subtype.tesla −0.25 [−0.38, −0.13]
−0.5 −0.4 −0.3 −0.2 −0.1
Observed Outcome

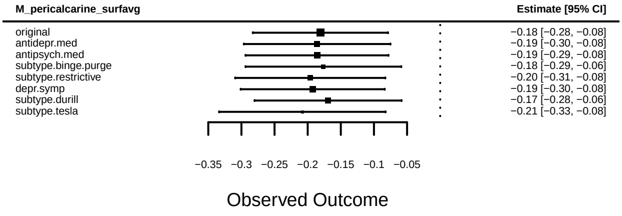
M_pericalcarine_surfavg
Estimate [95% CI]
original −0.18 [−0.28, −0.08]
antidepr.med −0.19 [−0.30, −0.08]
antipsych.med −0.19 [−0.29, −0.08]
subtype.binge.purge −0.18 [−0.29, −0.06]
subtype.restrictive −0.20 [−0.31, −0.08]
depr.symp −0.19 [−0.30, −0.08]
subtype.durill −0.17 [−0.28, −0.06]
subtype.tesla −0.21 [−0.33, −0.08]
−0.35 −0.3 −0.25 −0.2 −0.15 −0.1 −0.05
Observed Outcome

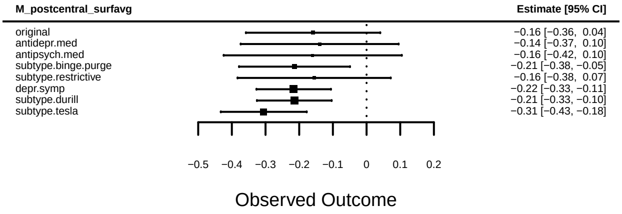
M_postcentral_surfavg
Estimate [95% CI]
original −0.16 [−0.36, 0.04]
antidepr.med −0.14 [−0.37, 0.10]
antipsych.med −0.16 [−0.42, 0.10]
subtype.binge.purge −0.21 [−0.38, −0.05]
subtype.restrictive −0.16 [−0.38, 0.07]
depr.symp −0.22 [−0.33, −0.11]
subtype.durill −0.21 [−0.33, −0.10]
subtype.tesla −0.31 [−0.43, −0.18]
−0.5 −0.4 −0.3 −0.2 −0.1 0 0.1 0.2
Observed Outcome

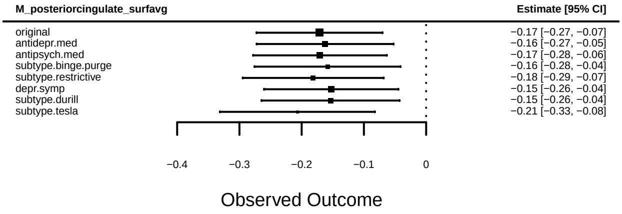
M_posteriorcingulate_surfavg
Estimate [95% CI]
original −0.17 [−0.27, −0.07]
antidepr.med −0.16 [−0.27, −0.05]
antipsych.med −0.17 [−0.28, −0.06]
subtype.binge.purge −0.16 [−0.28, −0.04]
subtype.restrictive −0.18 [−0.29, −0.07]
depr.symp −0.15 [−0.26, −0.04]
subtype.durill −0.15 [−0.26, −0.04]
subtype.tesla −0.21 [−0.33, −0.08]
−0.4 −0.3 −0.2 −0.1 0
Observed Outcome

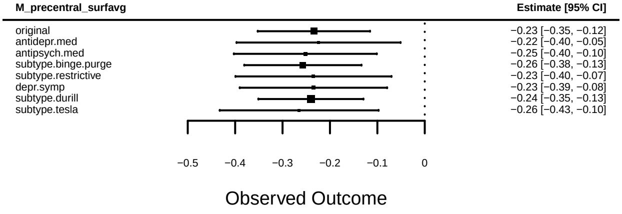
M_precentral_surfavg
Estimate [95% CI]
original −0.23 [−0.35, −0.12]
antidepr.med −0.22 [−0.40, −0.05]
antipsych.med −0.25 [−0.40, −0.10]
subtype.binge.purge −0.26 [−0.38, −0.13]
subtype.restrictive −0.23 [−0.40, −0.07]
depr.symp −0.23 [−0.39, −0.08]
subtype.durill −0.24 [−0.35, −0.13]
subtype.tesla −0.26 [−0.43, −0.10]
−0.5 −0.4 −0.3 −0.2 −0.1 0
Observed Outcome

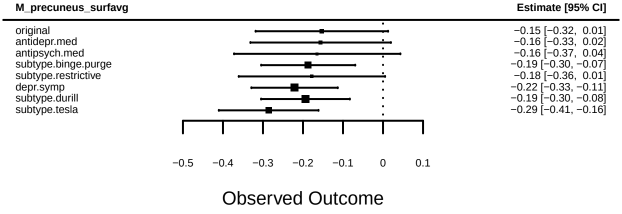
M_precuneus_surfavg
Estimate [95% CI]
original −0.15 [−0.32, 0.01]
antidepr.med −0.16 [−0.33, 0.02]
antipsych.med −0.16 [−0.37, 0.04]
subtype.binge.purge −0.19 [−0.30, −0.07]
subtype.restrictive −0.18 [−0.36, 0.01]
depr.symp −0.22 [−0.33, −0.11]
subtype.durill −0.19 [−0.30, −0.08]
subtype.tesla −0.29 [−0.41, −0.16]
−0.5 −0.4 −0.3 −0.2 −0.1 0 0.1
Observed Outcome

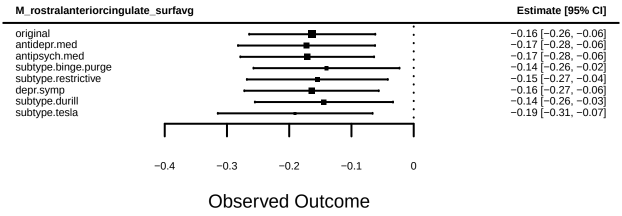
M_rostralanteriorcingulate_surfavg
Estimate [95% CI]
original −0.16 [−0.26, −0.06]
antidepr.med −0.17 [−0.28, −0.06]
antipsych.med −0.17 [−0.28, −0.06]
subtype.binge.purge −0.14 [−0.26, −0.02]
subtype.restrictive −0.15 [−0.27, −0.04]
depr.symp −0.16 [−0.27, −0.06]
subtype.durill −0.14 [−0.26, −0.03]
subtype.tesla −0.19 [−0.31, −0.07]
−0.4 −0.3 −0.2 −0.1 0
Observed Outcome

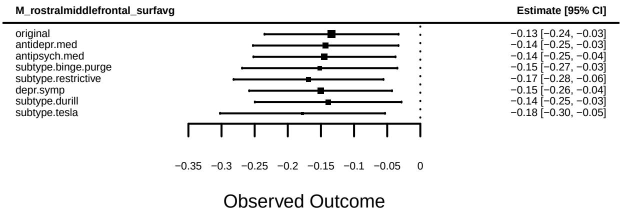
M_rostralmiddlefrontal_surfavg
Estimate [95% CI]
original −0.13 [−0.24, −0.03]
antidepr.med −0.14 [−0.25, −0.03]
antipsych.med −0.14 [−0.25, −0.04]
subtype.binge.purge −0.15 [−0.27, −0.03]
subtype.restrictive −0.17 [−0.28, −0.06]
depr.symp −0.15 [−0.26, −0.04]
subtype.durill −0.14 [−0.25, −0.03]
subtype.tesla −0.18 [−0.30, −0.05]
−0.35 −0.3 −0.25 −0.2 −0.15 −0.1 −0.05 0
Observed Outcome

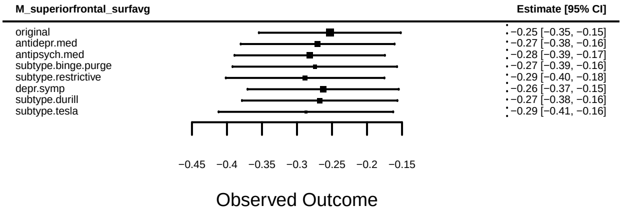
M_superiorfrontal_surfavg
Estimate [95% CI]
original −0.25 [−0.35, −0.15]
antidepr.med −0.27 [−0.38, −0.16]
antipsych.med −0.28 [−0.39, −0.17]
subtype.binge.purge −0.27 [−0.39, −0.16]
subtype.restrictive −0.29 [−0.40, −0.18]
depr.symp −0.26 [−0.37, −0.15]
subtype.durill −0.27 [−0.38, −0.16]
subtype.tesla −0.29 [−0.41, −0.16]
−0.45 −0.4 −0.35 −0.3 −0.25 −0.2 −0.15
Observed Outcome

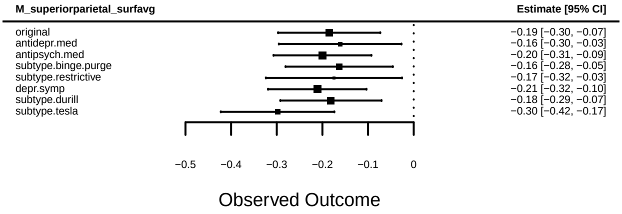
M_superiorparietal_surfavg
Estimate [95% CI]
original −0.19 [−0.30, −0.07]
antidepr.med −0.16 [−0.30, −0.03]
antipsych.med −0.20 [−0.31, −0.09]
subtype.binge.purge −0.16 [−0.28, −0.05]
subtype.restrictive −0.17 [−0.32, −0.03]
depr.symp −0.21 [−0.32, −0.10]
subtype.durill −0.18 [−0.29, −0.07]
subtype.tesla −0.30 [−0.42, −0.17]
−0.5 −0.4 −0.3 −0.2 −0.1 0
Observed Outcome

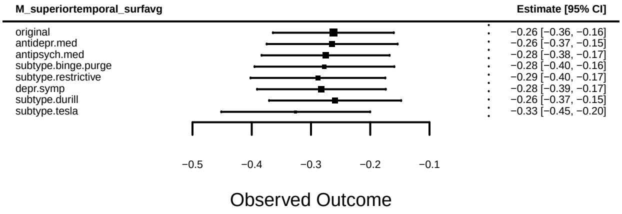
M_superiortemporal_surfavg
Estimate [95% CI]
original −0.26 [−0.36, −0.16]
antidepr.med −0.26 [−0.37, −0.15]
antipsych.med −0.28 [−0.38, −0.17]
subtype.binge.purge −0.28 [−0.40, −0.16]
subtype.restrictive −0.29 [−0.40, −0.17]
depr.symp −0.28 [−0.39, −0.17]
subtype.durill −0.26 [−0.37, −0.15]
subtype.tesla −0.33 [−0.45, −0.20]
−0.5 −0.4 −0.3 −0.2 −0.1
Observed Outcome

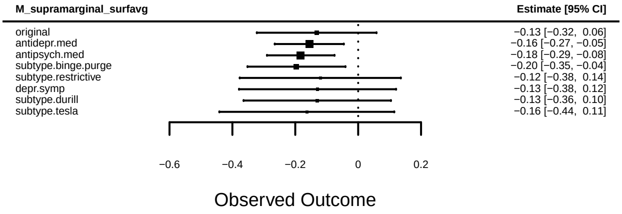
M_supramarginal_surfavg
Estimate [95% CI]
original −0.13 [−0.32, 0.06]
antidepr.med −0.16 [−0.27, −0.05]
antipsych.med −0.18 [−0.29, −0.08]
subtype.binge.purge −0.20 [−0.35, −0.04]
subtype.restrictive −0.12 [−0.38, 0.14]
depr.symp −0.13 [−0.38, 0.12]
subtype.durill −0.13 [−0.36, 0.10]
subtype.tesla −0.16 [−0.44, 0.11]
−0.6 −0.4 −0.2 0 0.2
Observed Outcome

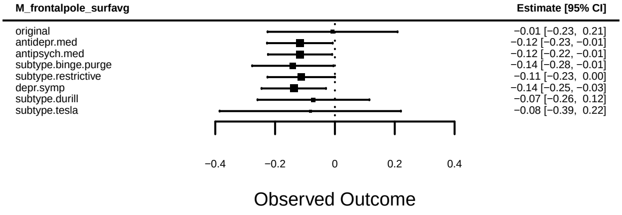
M_frontalpole_surfavg
Estimate [95% CI]
original −0.01 [−0.23, 0.21]
antidepr.med −0.12 [−0.23, −0.01]
antipsych.med −0.12 [−0.22, −0.01]
subtype.binge.purge −0.14 [−0.28, −0.01]
subtype.restrictive −0.11 [−0.23, 0.00]
depr.symp −0.14 [−0.25, −0.03]
subtype.durill −0.07 [−0.26, 0.12]
subtype.tesla −0.08 [−0.39, 0.22]
−0.4 −0.2 0 0.2 0.4
Observed Outcome

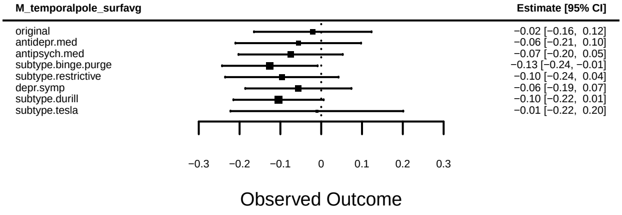
M_temporalpole_surfavg
Estimate [95% CI]
original −0.02 [−0.16, 0.12]
antidepr.med −0.06 [−0.21, 0.10]
antipsych.med −0.07 [−0.20, 0.05]
subtype.binge.purge −0.13 [−0.24, −0.01]
subtype.restrictive −0.10 [−0.24, 0.04]
depr.symp −0.06 [−0.19, 0.07]
subtype.durill −0.10 [−0.22, 0.01]
subtype.tesla −0.01 [−0.22, 0.20]
−0.3 −0.2 −0.1 0 0.1 0.2 0.3
Observed Outcome

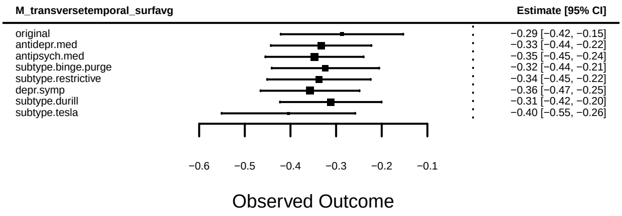
M_transversetemporal_surfavg
Estimate [95% CI]
original −0.29 [−0.42, −0.15]
antidepr.med −0.33 [−0.44, −0.22]
antipsych.med −0.35 [−0.45, −0.24]
subtype.binge.purge −0.32 [−0.44, −0.21]
subtype.restrictive −0.34 [−0.45, −0.22]
depr.symp −0.36 [−0.47, −0.25]
subtype.durill −0.31 [−0.42, −0.20]
subtype.tesla −0.40 [−0.55, −0.26]
−0.6 −0.5 −0.4 −0.3 −0.2 −0.1
Observed Outcome

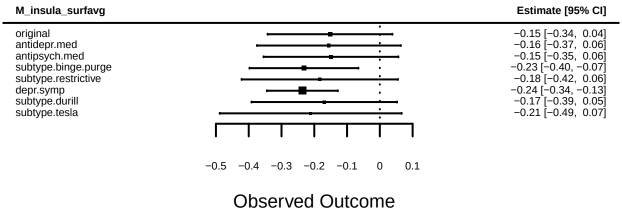
M_insula_surfavg
Estimate [95% CI]
original −0.15 [−0.34, 0.04]
antidepr.med −0.16 [−0.37, 0.06]
antipsych.med −0.15 [−0.35, 0.06]
subtype.binge.purge −0.23 [−0.40, −0.07]
subtype.restrictive −0.18 [−0.42, 0.06]
depr.symp −0.24 [−0.34, −0.13]
subtype.durill −0.17 [−0.39, 0.05]
subtype.tesla −0.21 [−0.49, 0.07]
−0.5 −0.4 −0.3 −0.2 −0.1 0 0.1
Observed Outcome

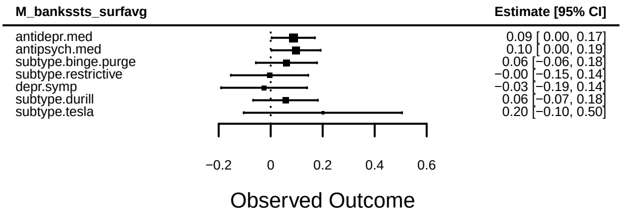
M_bankssts_surfavg
Estimate [95% CI]
antidepr.med 0.09 [ 0.00, 0.17]
antipsych.med 0.10 [ 0.00, 0.19]
subtype.binge.purge 0.06 [−0.06, 0.18]
subtype.restrictive −0.00 [−0.15, 0.14]
depr.symp −0.03 [−0.19, 0.14]
subtype.durill 0.06 [−0.07, 0.18]
subtype.tesla 0.20 [−0.10, 0.50]
−0.2 0 0.2 0.4 0.6
Observed Outcome

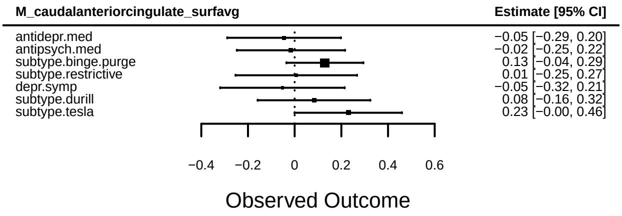
M_caudalanteriorcingulate_surfavg
Estimate [95% CI]
antidepr.med −0.05 [−0.29, 0.20]
antipsych.med −0.02 [−0.25, 0.22]
subtype.binge.purge 0.13 [−0.04, 0.29]
subtype.restrictive 0.01 [−0.25, 0.27]
depr.symp −0.05 [−0.32, 0.21]
subtype.durill 0.08 [−0.16, 0.32]
subtype.tesla 0.23 [−0.00, 0.46]
−0.4 −0.2 0 0.2 0.4 0.6
Observed Outcome

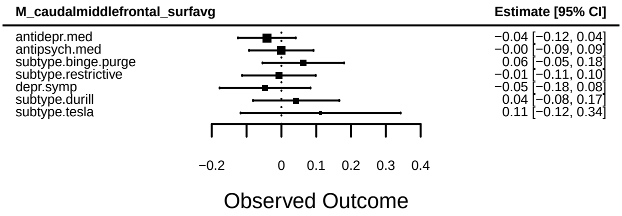
M_caudalmiddlefrontal_surfavg
Estimate [95% CI]
antidepr.med −0.04 [−0.12, 0.04]
antipsych.med −0.00 [−0.09, 0.09]
subtype.binge.purge 0.06 [−0.05, 0.18]
subtype.restrictive −0.01 [−0.11, 0.10]
depr.symp −0.05 [−0.18, 0.08]
subtype.durill 0.04 [−0.08, 0.17]
subtype.tesla 0.11 [−0.12, 0.34]
−0.2 0 0.1 0.2 0.3 0.4
Observed Outcome

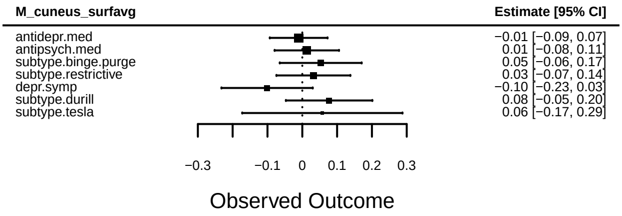
M_cuneus_surfavg
Estimate [95% CI]
antidepr.med −0.01 [−0.09, 0.07]
antipsych.med 0.01 [−0.08, 0.11]
subtype.binge.purge 0.05 [−0.06, 0.17]
subtype.restrictive 0.03 [−0.07, 0.14]
depr.symp −0.10 [−0.23, 0.03]
subtype.durill 0.08 [−0.05, 0.20]
subtype.tesla 0.06 [−0.17, 0.29]
−0.3 −0.1 0 0.1 0.2 0.3
Observed Outcome

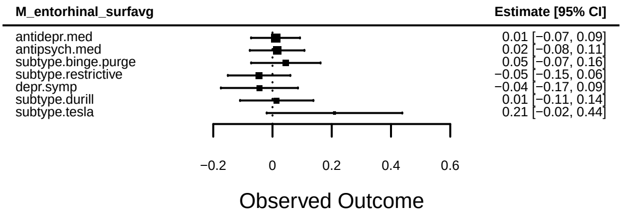
M_entorhinal_surfavg
Estimate [95% CI]
antidepr.med 0.01 [−0.07, 0.09]
antipsych.med 0.02 [−0.08, 0.11]
subtype.binge.purge 0.05 [−0.07, 0.16]
subtype.restrictive −0.05 [−0.15, 0.06]
depr.symp −0.04 [−0.17, 0.09]
subtype.durill 0.01 [−0.11, 0.14]
subtype.tesla 0.21 [−0.02, 0.44]
−0.2 0 0.2 0.4 0.6
Observed Outcome

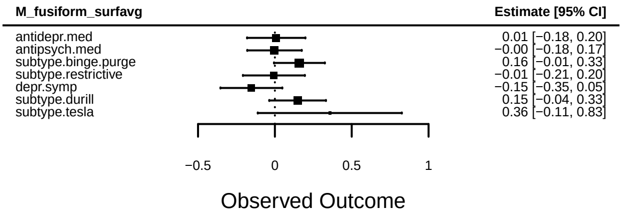
M_fusiform_surfavg
Estimate [95% CI]
antidepr.med 0.01 [−0.18, 0.20]
antipsych.med −0.00 [−0.18, 0.17]
subtype.binge.purge 0.16 [−0.01, 0.33]
subtype.restrictive −0.01 [−0.21, 0.20]
depr.symp −0.15 [−0.35, 0.05]
subtype.durill 0.15 [−0.04, 0.33]
subtype.tesla 0.36 [−0.11, 0.83]
−0.5 0 0.5 1
Observed Outcome

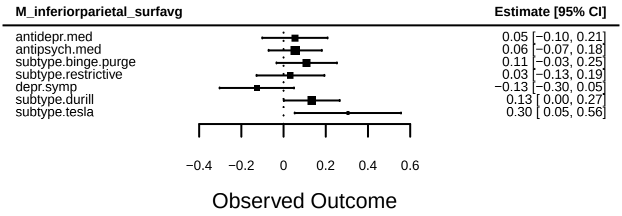
M_inferiorparietal_surfavg
Estimate [95% CI]
antidepr.med 0.05 [−0.10, 0.21]
antipsych.med 0.06 [−0.07, 0.18]
subtype.binge.purge 0.11 [−0.03, 0.25]
subtype.restrictive 0.03 [−0.13, 0.19]
depr.symp −0.13 [−0.30, 0.05]
subtype.durill 0.13 [ 0.00, 0.27]
subtype.tesla 0.30 [ 0.05, 0.56]
−0.4 −0.2 0 0.2 0.4 0.6
Observed Outcome

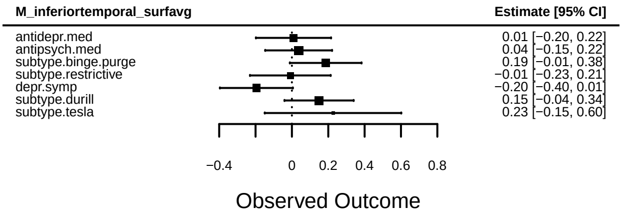
M_inferiortemporal_surfavg
Estimate [95% CI]
antidepr.med 0.01 [−0.20, 0.22]
antipsych.med 0.04 [−0.15, 0.22]
subtype.binge.purge 0.19 [−0.01, 0.38]
subtype.restrictive −0.01 [−0.23, 0.21]
depr.symp −0.20 [−0.40, 0.01]
subtype.durill 0.15 [−0.04, 0.34]
subtype.tesla 0.23 [−0.15, 0.60]
−0.4 0 0.2 0.4 0.6 0.8
Observed Outcome

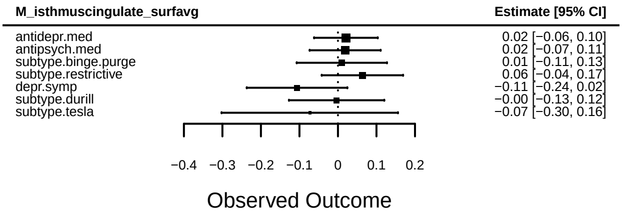
M_isthmuscingulate_surfavg
Estimate [95% CI]
antidepr.med 0.02 [−0.06, 0.10]
antipsych.med 0.02 [−0.07, 0.11]
subtype.binge.purge 0.01 [−0.11, 0.13]
subtype.restrictive 0.06 [−0.04, 0.17]
depr.symp −0.11 [−0.24, 0.02]
subtype.durill −0.00 [−0.13, 0.12]
subtype.tesla −0.07 [−0.30, 0.16]
−0.4 −0.3 −0.2 −0.1 0 0.1 0.2
Observed Outcome

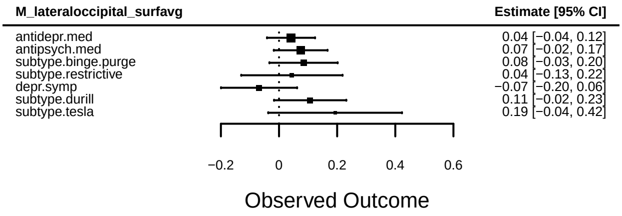
M_lateraloccipital_surfavg
Estimate [95% CI]
antidepr.med 0.04 [−0.04, 0.12]
antipsych.med 0.07 [−0.02, 0.17]
subtype.binge.purge 0.08 [−0.03, 0.20]
subtype.restrictive 0.04 [−0.13, 0.22]
depr.symp −0.07 [−0.20, 0.06]
subtype.durill 0.11 [−0.02, 0.23]
subtype.tesla 0.19 [−0.04, 0.42]
−0.2 0 0.2 0.4 0.6
Observed Outcome

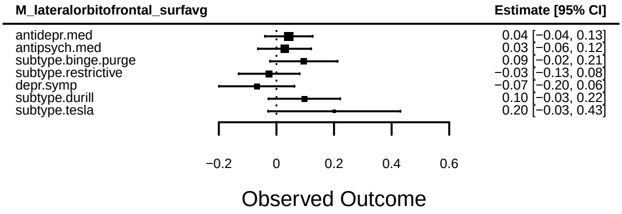
M_lateralorbitofrontal_surfavg
Estimate [95% CI]
antidepr.med 0.04 [−0.04, 0.13]
antipsych.med 0.03 [−0.06, 0.12]
subtype.binge.purge 0.09 [−0.02, 0.21]
subtype.restrictive −0.03 [−0.13, 0.08]
depr.symp −0.07 [−0.20, 0.06]
subtype.durill 0.10 [−0.03, 0.22]
subtype.tesla 0.20 [−0.03, 0.43]
−0.2 0 0.2 0.4 0.6
Observed Outcome

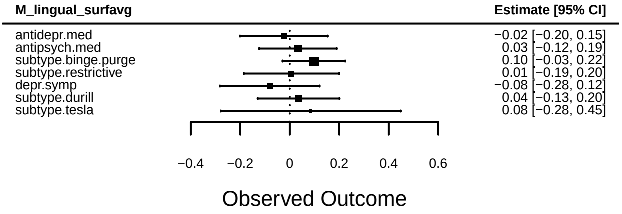
M_lingual_surfavg
Estimate [95% CI]
antidepr.med −0.02 [−0.20, 0.15]
antipsych.med 0.03 [−0.12, 0.19]
subtype.binge.purge 0.10 [−0.03, 0.22]
subtype.restrictive 0.01 [−0.19, 0.20]
depr.symp −0.08 [−0.28, 0.12]
subtype.durill 0.04 [−0.13, 0.20]
subtype.tesla 0.08 [−0.28, 0.45]
−0.4 −0.2 0 0.2 0.4 0.6
Observed Outcome

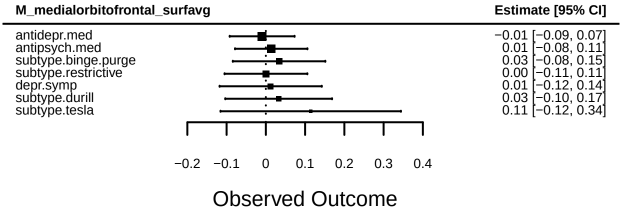
M_medialorbitofrontal_surfavg
Estimate [95% CI]
antidepr.med −0.01 [−0.09, 0.07]
antipsych.med 0.01 [−0.08, 0.11]
subtype.binge.purge 0.03 [−0.08, 0.15]
subtype.restrictive 0.00 [−0.11, 0.11]
depr.symp 0.01 [−0.12, 0.14]
subtype.durill 0.03 [−0.10, 0.17]
subtype.tesla 0.11 [−0.12, 0.34]
−0.2 −0.1 0 0.1 0.2 0.3 0.4
Observed Outcome

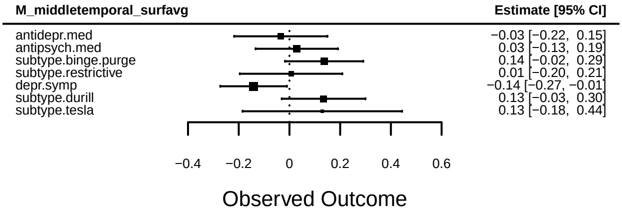
M_middletemporal_surfavg
Estimate [95% CI]
antidepr.med −0.03 [−0.22, 0.15]
antipsych.med 0.03 [−0.13, 0.19]
subtype.binge.purge 0.14 [−0.02, 0.29]
subtype.restrictive 0.01 [−0.20, 0.21]
depr.symp −0.14 [−0.27, −0.01]
subtype.durill 0.13 [−0.03, 0.30]
subtype.tesla 0.13 [−0.18, 0.44]
−0.4 −0.2 0 0.2 0.4 0.6
Observed Outcome

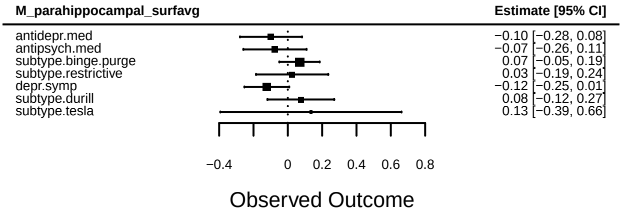
M_parahippocampal_surfavg
Estimate [95% CI]
antidepr.med −0.10 [−0.28, 0.08]
antipsych.med −0.07 [−0.26, 0.11]
subtype.binge.purge 0.07 [−0.05, 0.19]
subtype.restrictive 0.03 [−0.19, 0.24]
depr.symp −0.12 [−0.25, 0.01]
subtype.durill 0.08 [−0.12, 0.27]
subtype.tesla 0.13 [−0.39, 0.66]
−0.4 0 0.2 0.4 0.6 0.8
Observed Outcome

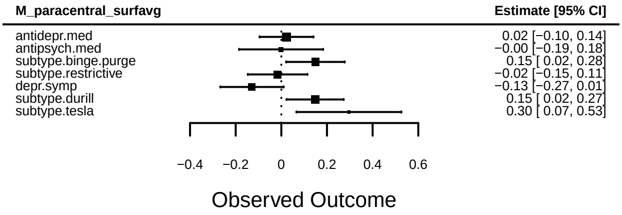
M_paracentral_surfavg
Estimate [95% CI]
antidepr.med 0.02 [−0.10, 0.14]
antipsych.med −0.00 [−0.19, 0.18]
subtype.binge.purge 0.15 [ 0.02, 0.28]
subtype.restrictive −0.02 [−0.15, 0.11]
depr.symp −0.13 [−0.27, 0.01]
subtype.durill 0.15 [ 0.02, 0.27]
subtype.tesla 0.30 [ 0.07, 0.53]
−0.4 −0.2 0 0.2 0.4 0.6
Observed Outcome

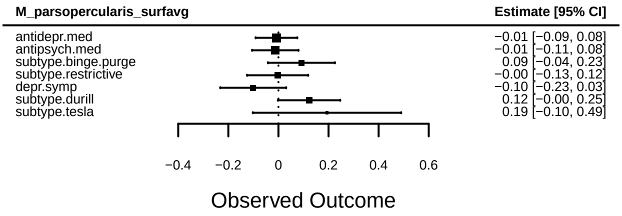
M_parsopercularis_surfavg
Estimate [95% CI]
antidepr.med −0.01 [−0.09, 0.08]
antipsych.med −0.01 [−0.11, 0.08]
subtype.binge.purge 0.09 [−0.04, 0.23]
subtype.restrictive −0.00 [−0.13, 0.12]
depr.symp −0.10 [−0.23, 0.03]
subtype.durill 0.12 [−0.00, 0.25]
subtype.tesla 0.19 [−0.10, 0.49]
−0.4 −0.2 0 0.2 0.4 0.6
Observed Outcome

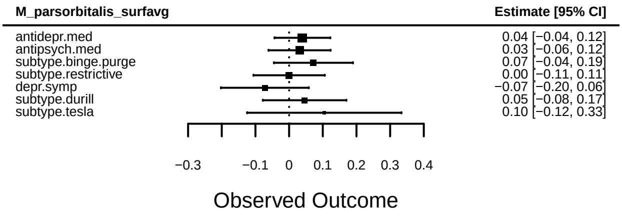
M_parsorbitalis_surfavg
Estimate [95% CI]
antidepr.med 0.04 [−0.04, 0.12]
antipsych.med 0.03 [−0.06, 0.12]
subtype.binge.purge 0.07 [−0.04, 0.19]
subtype.restrictive 0.00 [−0.11, 0.11]
depr.symp −0.07 [−0.20, 0.06]
subtype.durill 0.05 [−0.08, 0.17]
subtype.tesla 0.10 [−0.12, 0.33]
−0.3 −0.1 0 0.1 0.2 0.3 0.4
Observed Outcome

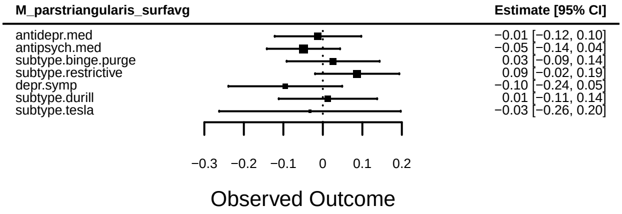
M_parstriangularis_surfavg
Estimate [95% CI]
antidepr.med −0.01 [−0.12, 0.10]
antipsych.med −0.05 [−0.14, 0.04]
subtype.binge.purge 0.03 [−0.09, 0.14]
subtype.restrictive 0.09 [−0.02, 0.19]
depr.symp −0.10 [−0.24, 0.05]
subtype.durill 0.01 [−0.11, 0.14]
subtype.tesla −0.03 [−0.26, 0.20]
−0.3 −0.2 −0.1 0 0.1 0.2
Observed Outcome

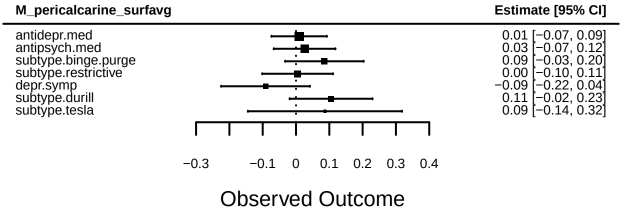
M_pericalcarine_surfavg
Estimate [95% CI]
antidepr.med 0.01 [−0.07, 0.09]
antipsych.med 0.03 [−0.07, 0.12]
subtype.binge.purge 0.09 [−0.03, 0.20]
subtype.restrictive 0.00 [−0.10, 0.11]
depr.symp −0.09 [−0.22, 0.04]
subtype.durill 0.11 [−0.02, 0.23]
subtype.tesla 0.09 [−0.14, 0.32]
−0.3 −0.1 0 0.1 0.2 0.3 0.4
Observed Outcome

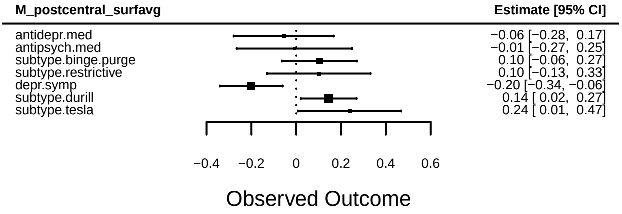
M_postcentral_surfavg
Estimate [95% CI]
antidepr.med −0.06 [−0.28, 0.17]
antipsych.med −0.01 [−0.27, 0.25]
subtype.binge.purge 0.10 [−0.06, 0.27]
subtype.restrictive 0.10 [−0.13, 0.33]
depr.symp −0.20 [−0.34, −0.06]
subtype.durill 0.14 [ 0.02, 0.27]
subtype.tesla 0.24 [ 0.01, 0.47]
−0.4 −0.2 0 0.2 0.4 0.6
Observed Outcome

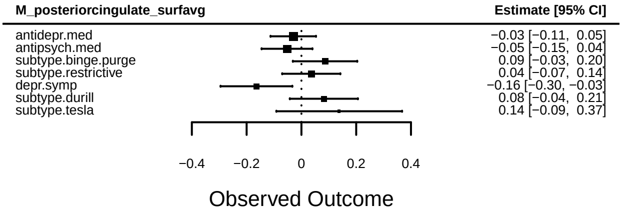
M_posteriorcingulate_surfavg
Estimate [95% CI]
antidepr.med −0.03 [−0.11, 0.05]
antipsych.med −0.05 [−0.15, 0.04]
subtype.binge.purge 0.09 [−0.03, 0.20]
subtype.restrictive 0.04 [−0.07, 0.14]
depr.symp −0.16 [−0.30, −0.03]
subtype.durill 0.08 [−0.04, 0.21]
subtype.tesla 0.14 [−0.09, 0.37]
−0.4 −0.2 0 0.2 0.4
Observed Outcome

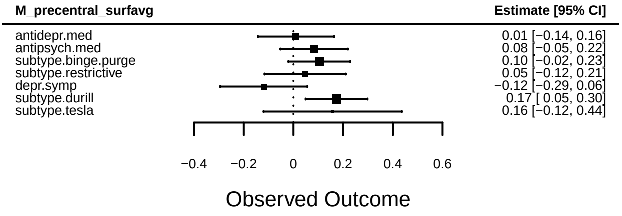
M_precentral_surfavg
Estimate [95% CI]
antidepr.med 0.01 [−0.14, 0.16]
antipsych.med 0.08 [−0.05, 0.22]
subtype.binge.purge 0.10 [−0.02, 0.23]
subtype.restrictive 0.05 [−0.12, 0.21]
depr.symp −0.12 [−0.29, 0.06]
subtype.durill 0.17 [ 0.05, 0.30]
subtype.tesla 0.16 [−0.12, 0.44]
−0.4 −0.2 0 0.2 0.4 0.6
Observed Outcome

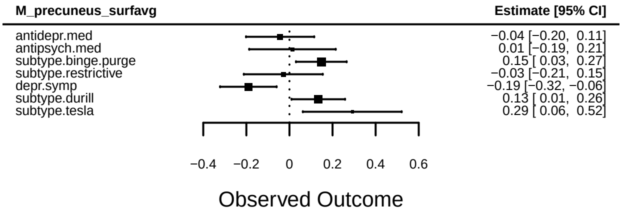
M_precuneus_surfavg
Estimate [95% CI]
antidepr.med −0.04 [−0.20, 0.11]
antipsych.med 0.01 [−0.19, 0.21]
subtype.binge.purge 0.15 [ 0.03, 0.27]
subtype.restrictive −0.03 [−0.21, 0.15]
depr.symp −0.19 [−0.32, −0.06]
subtype.durill 0.13 [ 0.01, 0.26]
subtype.tesla 0.29 [ 0.06, 0.52]
−0.4 −0.2 0 0.2 0.4 0.6
Observed Outcome

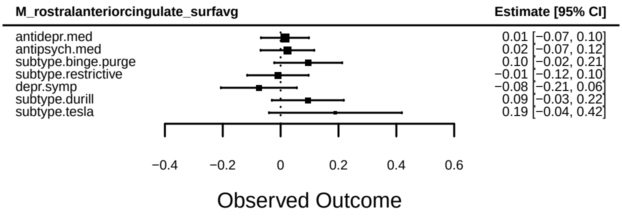
M_rostralanteriorcingulate_surfavg
Estimate [95% CI]
antidepr.med 0.01 [−0.07, 0.10]
antipsych.med 0.02 [−0.07, 0.12]
subtype.binge.purge 0.10 [−0.02, 0.21]
subtype.restrictive −0.01 [−0.12, 0.10]
depr.symp −0.08 [−0.21, 0.06]
subtype.durill 0.09 [−0.03, 0.22]
subtype.tesla 0.19 [−0.04, 0.42]
−0.4 −0.2 0 0.2 0.4 0.6
Observed Outcome

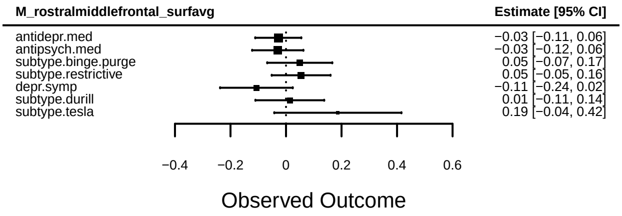
M_rostralmiddlefrontal_surfavg
Estimate [95% CI]
antidepr.med −0.03 [−0.11, 0.06]
antipsych.med −0.03 [−0.12, 0.06]
subtype.binge.purge 0.05 [−0.07, 0.17]
subtype.restrictive 0.05 [−0.05, 0.16]
depr.symp −0.11 [−0.24, 0.02]
subtype.durill 0.01 [−0.11, 0.14]
subtype.tesla 0.19 [−0.04, 0.42]
−0.4 −0.2 0 0.2 0.4 0.6
Observed Outcome

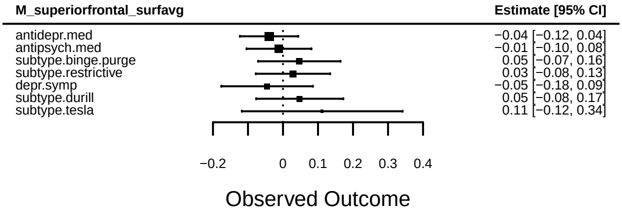
M_superiorfrontal_surfavg
Estimate [95% CI]
antidepr.med −0.04 [−0.12, 0.04]
antipsych.med −0.01 [−0.10, 0.08]
subtype.binge.purge 0.05 [−0.07, 0.16]
subtype.restrictive 0.03 [−0.08, 0.13]
depr.symp −0.05 [−0.18, 0.09]
subtype.durill 0.05 [−0.08, 0.17]
subtype.tesla 0.11 [−0.12, 0.34]
−0.2 0 0.1 0.2 0.3 0.4
Observed Outcome

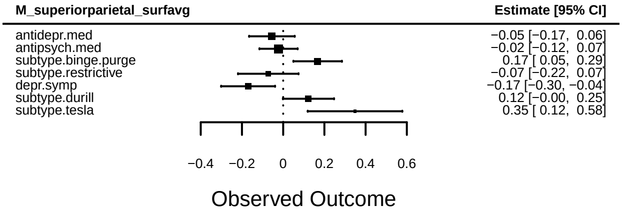
M_superiorparietal_surfavg
Estimate [95% CI]
antidepr.med −0.05 [−0.17, 0.06]
antipsych.med −0.02 [−0.12, 0.07]
subtype.binge.purge 0.17 [ 0.05, 0.29]
subtype.restrictive −0.07 [−0.22, 0.07]
depr.symp −0.17 [−0.30, −0.04]
subtype.durill 0.12 [−0.00, 0.25]
subtype.tesla 0.35 [ 0.12, 0.58]
−0.4 −0.2 0 0.2 0.4 0.6
Observed Outcome

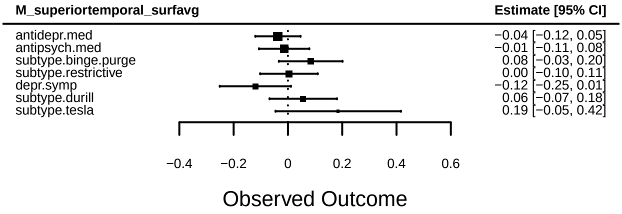
M_superiortemporal_surfavg
Estimate [95% CI]
antidepr.med −0.04 [−0.12, 0.05]
antipsych.med −0.01 [−0.11, 0.08]
subtype.binge.purge 0.08 [−0.03, 0.20]
subtype.restrictive 0.00 [−0.10, 0.11]
depr.symp −0.12 [−0.25, 0.01]
subtype.durill 0.06 [−0.07, 0.18]
subtype.tesla 0.19 [−0.05, 0.42]
−0.4 −0.2 0 0.2 0.4 0.6
Observed Outcome

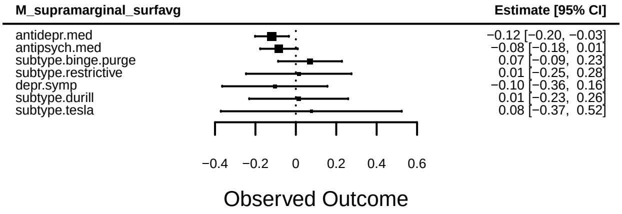
M_supramarginal_surfavg
Estimate [95% CI]
antidepr.med −0.12 [−0.20, −0.03]
antipsych.med −0.08 [−0.18, 0.01]
subtype.binge.purge 0.07 [−0.09, 0.23]
subtype.restrictive 0.01 [−0.25, 0.28]
depr.symp −0.10 [−0.36, 0.16]
subtype.durill 0.01 [−0.23, 0.26]
subtype.tesla 0.08 [−0.37, 0.52]
−0.4 −0.2 0 0.2 0.4 0.6
Observed Outcome

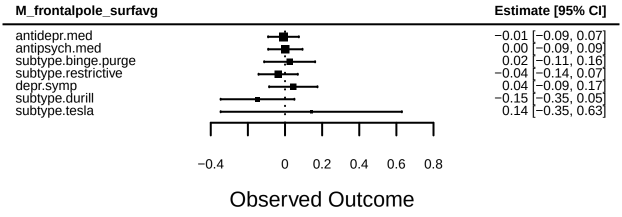
M_frontalpole_surfavg
Estimate [95% CI]
antidepr.med −0.01 [−0.09, 0.07]
antipsych.med 0.00 [−0.09, 0.09]
subtype.binge.purge 0.02 [−0.11, 0.16]
subtype.restrictive −0.04 [−0.14, 0.07]
depr.symp 0.04 [−0.09, 0.17]
subtype.durill −0.15 [−0.35, 0.05]
subtype.tesla 0.14 [−0.35, 0.63]
−0.4 0 0.2 0.4 0.6 0.8
Observed Outcome

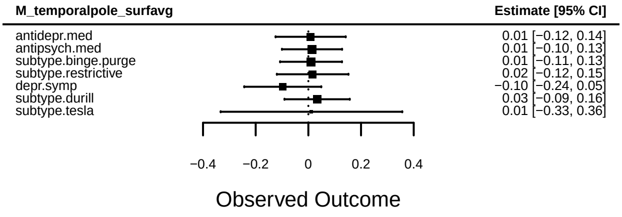
M_temporalpole_surfavg
Estimate [95% CI]
antidepr.med 0.01 [−0.12, 0.14]
antipsych.med 0.01 [−0.10, 0.13]
subtype.binge.purge 0.01 [−0.11, 0.13]
subtype.restrictive 0.02 [−0.12, 0.15]
depr.symp −0.10 [−0.24, 0.05]
subtype.durill 0.03 [−0.09, 0.16]
subtype.tesla 0.01 [−0.33, 0.36]
−0.4 −0.2 0 0.2 0.4
Observed Outcome

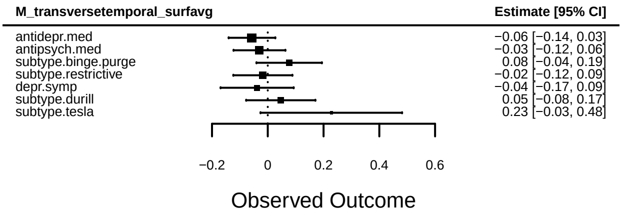
M_transversetemporal_surfavg
Estimate [95% CI]
antidepr.med −0.06 [−0.14, 0.03]
antipsych.med −0.03 [−0.12, 0.06]
subtype.binge.purge 0.08 [−0.04, 0.19]
subtype.restrictive −0.02 [−0.12, 0.09]
depr.symp −0.04 [−0.17, 0.09]
subtype.durill 0.05 [−0.08, 0.17]
subtype.tesla 0.23 [−0.03, 0.48]
−0.2
0
0.2
0.4
0.6
Observed Outcome

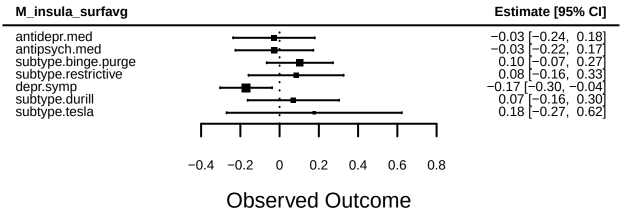
M_insula_surfavg
Estimate [95% CI]
antidepr.med −0.03 [−0.24, 0.18]
antipsych.med −0.03 [−0.22, 0.17]
subtype.binge.purge 0.10 [−0.07, 0.27]
subtype.restrictive 0.08 [−0.16, 0.33]
depr.symp −0.17 [−0.30, −0.04]
subtype.durill 0.07 [−0.16, 0.30]
subtype.tesla 0.18 [−0.27, 0.62]
−0.4
−0.2
0
0.2
0.4
0.6
0.8
Observed Outcome

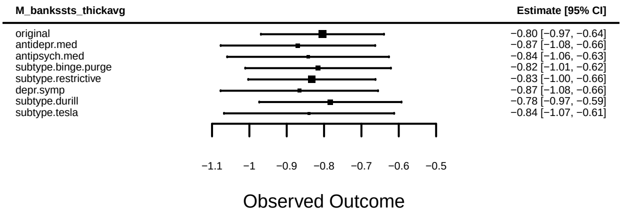
M_bankssts_thickavg
Estimate [95% CI]
original −0.80 [−0.97, −0.64]
antidepr.med −0.87 [−1.08, −0.66]
antipsych.med −0.84 [−1.06, −0.63]
subtype.binge.purge −0.82 [−1.01, −0.62]
subtype.restrictive −0.83 [−1.00, −0.66]
depr.symp −0.87 [−1.08, −0.66]
subtype.durill −0.78 [−0.97, −0.59]
subtype.tesla −0.84 [−1.07, −0.61]
−1.1 −1 −0.9 −0.8 −0.7 −0.6 −0.5
Observed Outcome

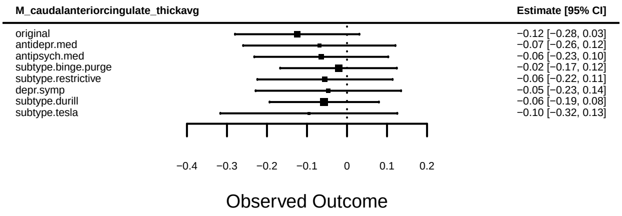
M_caudalanteriorcingulate_thickavg
Estimate [95% CI]
original −0.12 [−0.28, 0.03]
antidepr.med −0.07 [−0.26, 0.12]
antipsych.med −0.06 [−0.23, 0.10]
subtype.binge.purge −0.02 [−0.17, 0.12]
subtype.restrictive −0.06 [−0.22, 0.11]
depr.symp −0.05 [−0.23, 0.14]
subtype.durill −0.06 [−0.19, 0.08]
subtype.tesla −0.10 [−0.32, 0.13]
−0.4 −0.3 −0.2 −0.1 0 0.1 0.2
Observed Outcome

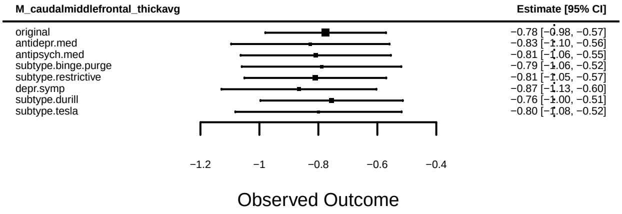
M_caudalmiddlefrontal_thickavg
Estimate [95% CI]
original −0.78 [−0.98, −0.57]
antidepr.med −0.83 [−1.10, −0.56]
antipsych.med −0.81 [−1.06, −0.55]
subtype.binge.purge −0.79 [−1.06, −0.52]
subtype.restrictive −0.81 [−1.05, −0.57]
depr.symp −0.87 [−1.13, −0.60]
subtype.durill −0.76 [−1.00, −0.51]
subtype.tesla −0.80 [−1.08, −0.52]
−1.2 −1 −0.8 −0.6 −0.4
Observed Outcome

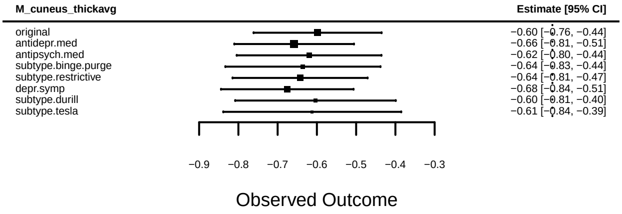
M_cuneus_thickavg
Estimate [95% CI]
original −0.60 [−0.76, −0.44]
antidepr.med −0.66 [−0.81, −0.51]
antipsych.med −0.62 [−0.80, −0.44]
subtype.binge.purge −0.64 [−0.83, −0.44]
subtype.restrictive −0.64 [−0.81, −0.47]
depr.symp −0.68 [−0.84, −0.51]
subtype.durill −0.60 [−0.81, −0.40]
subtype.tesla −0.61 [−0.84, −0.39]
−0.9 −0.8 −0.7 −0.6 −0.5 −0.4 −0.3
Observed Outcome

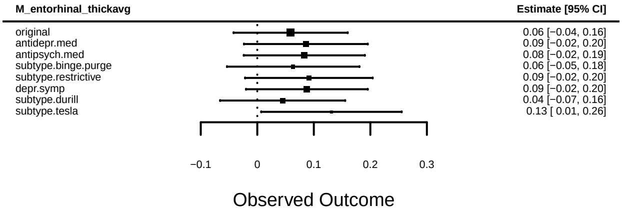
M_entorhinal_thickavg
Estimate [95% CI]
original 0.06 [−0.04, 0.16]
antidepr.med 0.09 [−0.02, 0.20]
antipsych.med 0.08 [−0.02, 0.19]
subtype.binge.purge 0.06 [−0.05, 0.18]
subtype.restrictive 0.09 [−0.02, 0.20]
depr.symp 0.09 [−0.02, 0.20]
subtype.durill 0.04 [−0.07, 0.16]
subtype.tesla 0.13 [ 0.01, 0.26]
−0.1 0 0.1 0.2 0.3
Observed Outcome

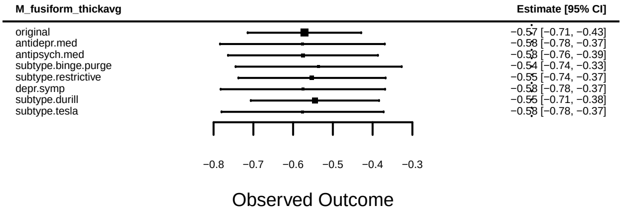
M_fusiform_thickavg
Estimate [95% CI]
original −0.57 [−0.71, −0.43]
antidepr.med −0.58 [−0.78, −0.37]
antipsych.med −0.58 [−0.76, −0.39]
subtype.binge.purge −0.54 [−0.74, −0.33]
subtype.restrictive −0.55 [−0.74, −0.37]
depr.symp −0.58 [−0.78, −0.37]
subtype.durill −0.55 [−0.71, −0.38]
subtype.tesla −0.58 [−0.78, −0.37]
−0.8 −0.7 −0.6 −0.5 −0.4 −0.3
Observed Outcome

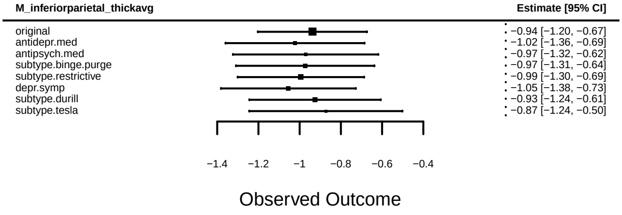
M_inferiorparietal_thickavg
Estimate [95% CI]
original −0.94 [−1.20, −0.67]
antidepr.med −1.02 [−1.36, −0.69]
antipsych.med −0.97 [−1.32, −0.62]
subtype.binge.purge −0.97 [−1.31, −0.64]
subtype.restrictive −0.99 [−1.30, −0.69]
depr.symp −1.05 [−1.38, −0.73]
subtype.durill −0.93 [−1.24, −0.61]
subtype.tesla −0.87 [−1.24, −0.50]
−1.4 −1.2 −1 −0.8 −0.6 −0.4
Observed Outcome

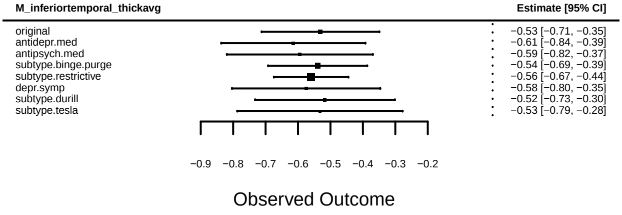
M_inferiortemporal_thickavg
Estimate [95% CI]
original −0.53 [−0.71, −0.35]
antidepr.med −0.61 [−0.84, −0.39]
antipsych.med −0.59 [−0.82, −0.37]
subtype.binge.purge −0.54 [−0.69, −0.39]
subtype.restrictive −0.56 [−0.67, −0.44]
depr.symp −0.58 [−0.80, −0.35]
subtype.durill −0.52 [−0.73, −0.30]
subtype.tesla −0.53 [−0.79, −0.28]
−0.9 −0.8 −0.7 −0.6 −0.5 −0.4 −0.3 −0.2
Observed Outcome

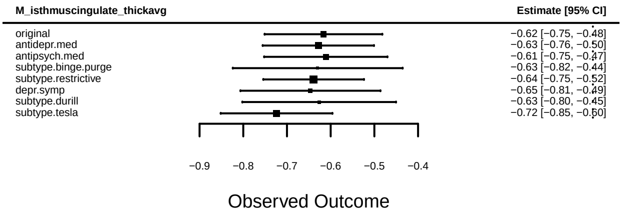
M_isthmuscingulate_thickavg
Estimate [95% CI]
original −0.62 [−0.75, −0.48]
antidepr.med −0.63 [−0.76, −0.50]
antipsych.med −0.61 [−0.75, −0.47]
subtype.binge.purge −0.63 [−0.82, −0.44]
subtype.restrictive −0.64 [−0.75, −0.52]
depr.symp −0.65 [−0.81, −0.49]
subtype.durill −0.63 [−0.80, −0.45]
subtype.tesla −0.72 [−0.85, −0.60]
−0.9 −0.8 −0.7 −0.6 −0.5 −0.4
Observed Outcome

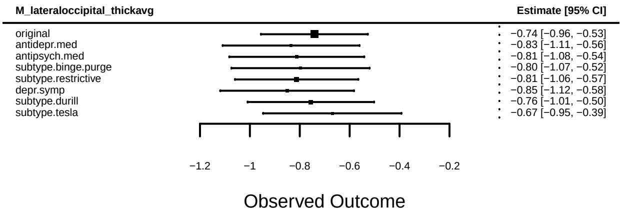
M_lateraloccipital_thickavg
Estimate [95% CI]
original −0.74 [−0.96, −0.53]
antidepr.med −0.83 [−1.11, −0.56]
antipsych.med −0.81 [−1.08, −0.54]
subtype.binge.purge −0.80 [−1.07, −0.52]
subtype.restrictive −0.81 [−1.06, −0.57]
depr.symp −0.85 [−1.12, −0.58]
subtype.durill −0.76 [−1.01, −0.50]
subtype.tesla −0.67 [−0.95, −0.39]
−1.2 −1 −0.8 −0.6 −0.4 −0.2
Observed Outcome

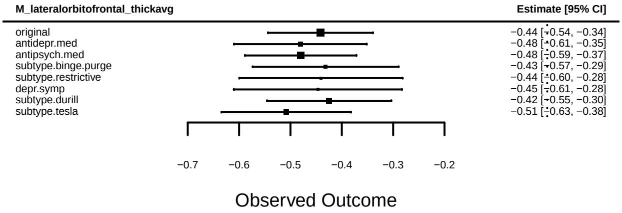
M_lateralorbitofrontal_thickavg
Estimate [95% CI]
original −0.44 [−0.54, −0.34]
antidepr.med −0.48 [−0.61, −0.35]
antipsych.med −0.48 [−0.59, −0.37]
subtype.binge.purge −0.43 [−0.57, −0.29]
subtype.restrictive −0.44 [−0.60, −0.28]
depr.symp −0.45 [−0.61, −0.28]
subtype.durill −0.42 [−0.55, −0.30]
subtype.tesla −0.51 [−0.63, −0.38]
−0.7 −0.6 −0.5 −0.4 −0.3 −0.2
Observed Outcome

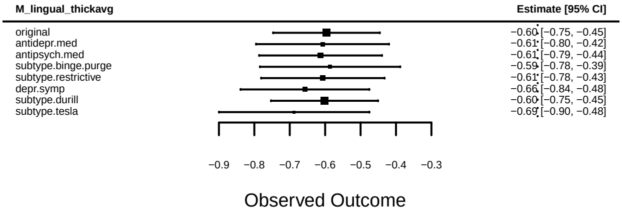
M_lingual_thickavg
Estimate [95% CI]
original −0.60 [−0.75, −0.45]
antidepr.med −0.61 [−0.80, −0.42]
antipsych.med −0.61 [−0.79, −0.44]
subtype.binge.purge −0.59 [−0.78, −0.39]
subtype.restrictive −0.61 [−0.78, −0.43]
depr.symp −0.66 [−0.84, −0.48]
subtype.durill −0.60 [−0.75, −0.45]
subtype.tesla −0.69 [−0.90, −0.48]
−0.9 −0.8 −0.7 −0.6 −0.5 −0.4 −0.3
Observed Outcome

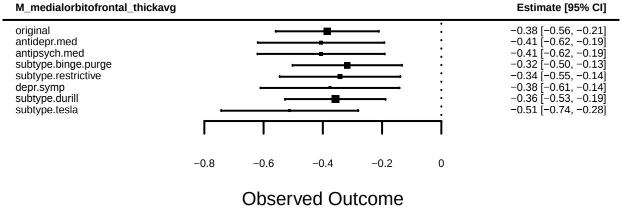
M_medialorbitofrontal_thickavg
Estimate [95% CI]
original −0.38 [−0.56, −0.21]
antidepr.med −0.41 [−0.62, −0.19]
antipsych.med −0.41 [−0.62, −0.19]
subtype.binge.purge −0.32 [−0.50, −0.13]
subtype.restrictive −0.34 [−0.55, −0.14]
depr.symp −0.38 [−0.61, −0.14]
subtype.durill −0.36 [−0.53, −0.19]
subtype.tesla −0.51 [−0.74, −0.28]
−0.8 −0.6 −0.4 −0.2 0
Observed Outcome

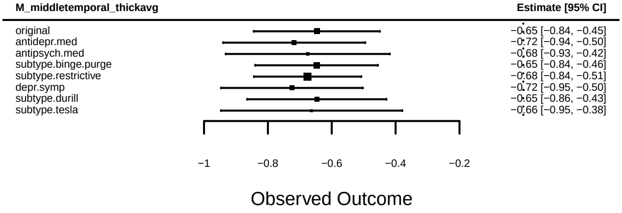
M_middletemporal_thickavg
Estimate [95% CI]
original −0.65 [−0.84, −0.45]
antidepr.med −0.72 [−0.94, −0.50]
antipsych.med −0.68 [−0.93, −0.42]
subtype.binge.purge −0.65 [−0.84, −0.46]
subtype.restrictive −0.68 [−0.84, −0.51]
depr.symp −0.72 [−0.95, −0.50]
subtype.durill −0.65 [−0.86, −0.43]
subtype.tesla −0.66 [−0.95, −0.38]
−1 −0.8 −0.6 −0.4 −0.2
Observed Outcome

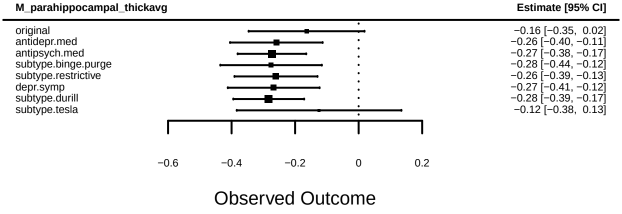
M_parahippocampal_thickavg
Estimate [95% CI]
original −0.16 [−0.35, 0.02]
antidepr.med −0.26 [−0.40, −0.11]
antipsych.med −0.27 [−0.38, −0.17]
subtype.binge.purge −0.28 [−0.44, −0.12]
subtype.restrictive −0.26 [−0.39, −0.13]
depr.symp −0.27 [−0.41, −0.12]
subtype.durill −0.28 [−0.39, −0.17]
subtype.tesla −0.12 [−0.38, 0.13]
−0.6 −0.4 −0.2 0 0.2
Observed Outcome

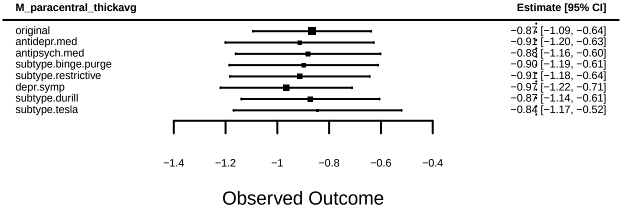
M_paracentral_thickavg
Estimate [95% CI]
original −0.87 [−1.09, −0.64]
antidepr.med −0.91 [−1.20, −0.63]
antipsych.med −0.88 [−1.16, −0.60]
subtype.binge.purge −0.90 [−1.19, −0.61]
subtype.restrictive −0.91 [−1.18, −0.64]
depr.symp −0.97 [−1.22, −0.71]
subtype.durill −0.87 [−1.14, −0.61]
subtype.tesla −0.84 [−1.17, −0.52]
−1.4 −1.2 −1 −0.8 −0.6 −0.4
Observed Outcome

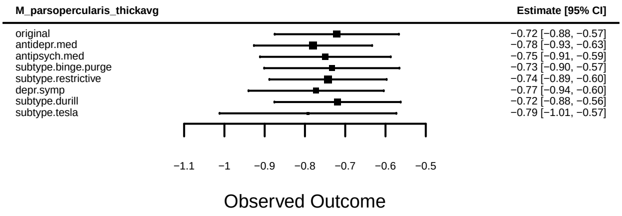
M_parsopercularis_thickavg
Estimate [95% CI]
original −0.72 [−0.88, −0.57]
antidepr.med −0.78 [−0.93, −0.63]
antipsych.med −0.75 [−0.91, −0.59]
subtype.binge.purge −0.73 [−0.90, −0.57]
subtype.restrictive −0.74 [−0.89, −0.60]
depr.symp −0.77 [−0.94, −0.60]
subtype.durill −0.72 [−0.88, −0.56]
subtype.tesla −0.79 [−1.01, −0.57]
−1.1 −1 −0.9 −0.8 −0.7 −0.6 −0.5
Observed Outcome

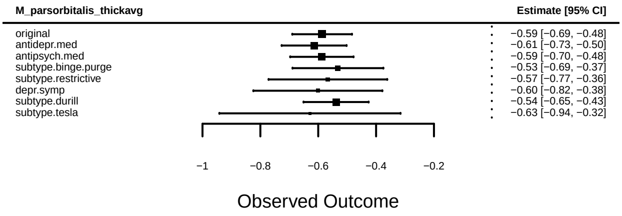
M_parsorbitalis_thickavg
Estimate [95% CI]
original −0.59 [−0.69, −0.48]
antidepr.med −0.61 [−0.73, −0.50]
antipsych.med −0.59 [−0.70, −0.48]
subtype.binge.purge −0.53 [−0.69, −0.37]
subtype.restrictive −0.57 [−0.77, −0.36]
depr.symp −0.60 [−0.82, −0.38]
subtype.durill −0.54 [−0.65, −0.43]
subtype.tesla −0.63 [−0.94, −0.32]
−1 −0.8 −0.6 −0.4 −0.2
Observed Outcome

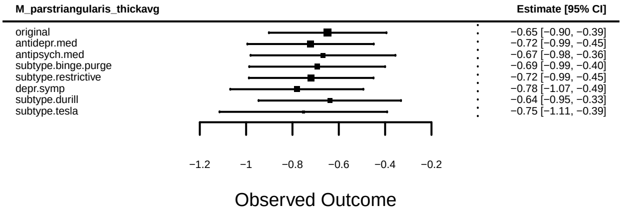
M_parstriangularis_thickavg
Estimate [95% CI]
original −0.65 [−0.90, −0.39]
antidepr.med −0.72 [−0.99, −0.45]
antipsych.med −0.67 [−0.98, −0.36]
subtype.binge.purge −0.69 [−0.99, −0.40]
subtype.restrictive −0.72 [−0.99, −0.45]
depr.symp −0.78 [−1.07, −0.49]
subtype.durill −0.64 [−0.95, −0.33]
subtype.tesla −0.75 [−1.11, −0.39]
−1.2 −1 −0.8 −0.6 −0.4 −0.2
Observed Outcome

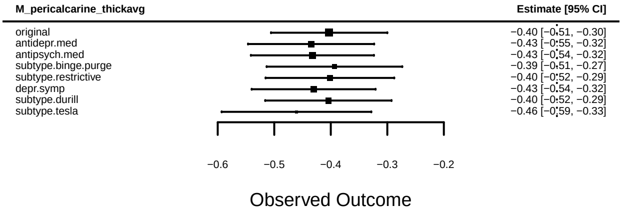
M_pericalcarine_thickavg
Estimate [95% CI]
original −0.40 [−0.51, −0.30]
antidepr.med −0.43 [−0.55, −0.32]
antipsych.med −0.43 [−0.54, −0.32]
subtype.binge.purge −0.39 [−0.51, −0.27]
subtype.restrictive −0.40 [−0.52, −0.29]
depr.symp −0.43 [−0.54, −0.32]
subtype.durill −0.40 [−0.52, −0.29]
subtype.tesla −0.46 [−0.59, −0.33]
−0.6 −0.5 −0.4 −0.3 −0.2
Observed Outcome

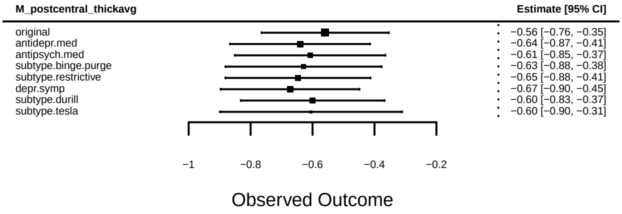
M_postcentral_thickavg
Estimate [95% CI]
original −0.56 [−0.76, −0.35]
antidepr.med −0.64 [−0.87, −0.41]
antipsych.med −0.61 [−0.85, −0.37]
subtype.binge.purge −0.63 [−0.88, −0.38]
subtype.restrictive −0.65 [−0.88, −0.41]
depr.symp −0.67 [−0.90, −0.45]
subtype.durill −0.60 [−0.83, −0.37]
subtype.tesla −0.60 [−0.90, −0.31]
−1 −0.8 −0.6 −0.4 −0.2
Observed Outcome

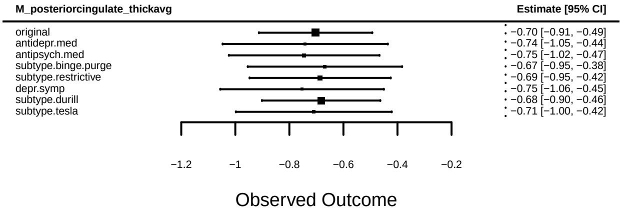
M_posteriorcingulate_thickavg
Estimate [95% CI]
original −0.70 [−0.91, −0.49]
antidepr.med −0.74 [−1.05, −0.44]
antipsych.med −0.75 [−1.02, −0.47]
subtype.binge.purge −0.67 [−0.95, −0.38]
subtype.restrictive −0.69 [−0.95, −0.42]
depr.symp −0.75 [−1.06, −0.45]
subtype.durill −0.68 [−0.90, −0.46]
subtype.tesla −0.71 [−1.00, −0.42]
−1.2 −1 −0.8 −0.6 −0.4 −0.2
Observed Outcome

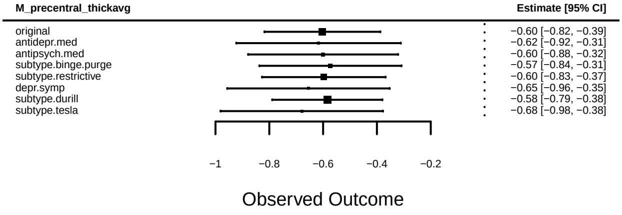
M_precentral_thickavg
Estimate [95% CI]
original −0.60 [−0.82, −0.39]
antidepr.med −0.62 [−0.92, −0.31]
antipsych.med −0.60 [−0.88, −0.32]
subtype.binge.purge −0.57 [−0.84, −0.31]
subtype.restrictive −0.60 [−0.83, −0.37]
depr.symp −0.65 [−0.96, −0.35]
subtype.durill −0.58 [−0.79, −0.38]
subtype.tesla −0.68 [−0.98, −0.38]
−1 −0.8 −0.6 −0.4 −0.2
Observed Outcome

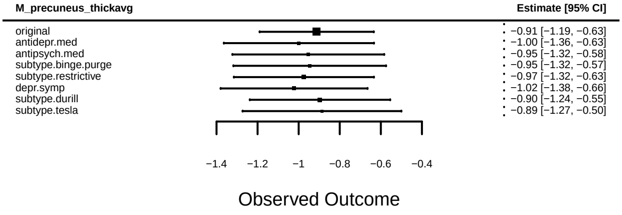
M_precuneus_thickavg
Estimate [95% CI]
original −0.91 [−1.19, −0.63]
antidepr.med −1.00 [−1.36, −0.63]
antipsych.med −0.95 [−1.32, −0.58]
subtype.binge.purge −0.95 [−1.32, −0.57]
subtype.restrictive −0.97 [−1.32, −0.63]
depr.symp −1.02 [−1.38, −0.66]
subtype.durill −0.90 [−1.24, −0.55]
subtype.tesla −0.89 [−1.27, −0.50]
−1.4 −1.2 −1 −0.8 −0.6 −0.4
Observed Outcome

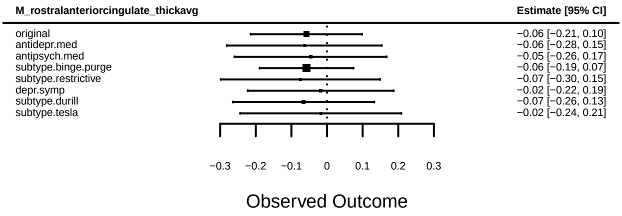
M_rostralanteriorcingulate_thickavg
Estimate [95% CI]
original −0.06 [−0.21, 0.10]
antidepr.med −0.06 [−0.28, 0.15]
antipsych.med −0.05 [−0.26, 0.17]
subtype.binge.purge −0.06 [−0.19, 0.07]
subtype.restrictive −0.07 [−0.30, 0.15]
depr.symp −0.02 [−0.22, 0.19]
subtype.durill −0.07 [−0.26, 0.13]
subtype.tesla −0.02 [−0.24, 0.21]
−0.3 −0.2 −0.1 0 0.1 0.2 0.3
Observed Outcome

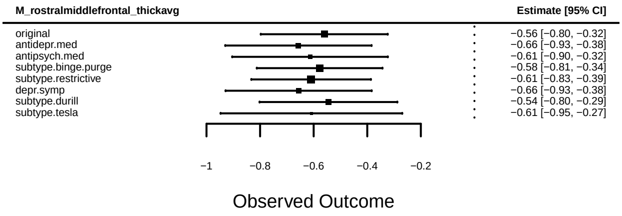
M_rostralmiddlefrontal_thickavg
Estimate [95% CI]
original −0.56 [−0.80, −0.32]
antidepr.med −0.66 [−0.93, −0.38]
antipsych.med −0.61 [−0.90, −0.32]
subtype.binge.purge −0.58 [−0.81, −0.34]
subtype.restrictive −0.61 [−0.83, −0.39]
depr.symp −0.66 [−0.93, −0.38]
subtype.durill −0.54 [−0.80, −0.29]
subtype.tesla −0.61 [−0.95, −0.27]
−1 −0.8 −0.6 −0.4 −0.2
Observed Outcome

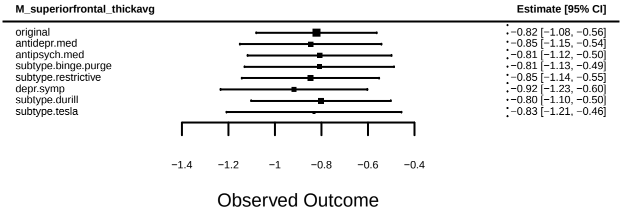
M_superiorfrontal_thickavg
Estimate [95% CI]
original −0.82 [−1.08, −0.56]
antidepr.med −0.85 [−1.15, −0.54]
antipsych.med −0.81 [−1.12, −0.50]
subtype.binge.purge −0.81 [−1.13, −0.49]
subtype.restrictive −0.85 [−1.14, −0.55]
depr.symp −0.92 [−1.23, −0.60]
subtype.durill −0.80 [−1.10, −0.50]
subtype.tesla −0.83 [−1.21, −0.46]
−1.4 −1.2 −1 −0.8 −0.6 −0.4
Observed Outcome

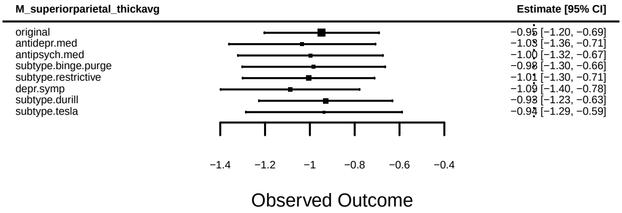
M_superiorparietal_thickavg
Estimate [95% CI]
original −0.95 [−1.20, −0.69]
antidepr.med −1.03 [−1.36, −0.71]
antipsych.med −1.00 [−1.32, −0.67]
subtype.binge.purge −0.98 [−1.30, −0.66]
subtype.restrictive −1.01 [−1.30, −0.71]
depr.symp −1.09 [−1.40, −0.78]
subtype.durill −0.93 [−1.23, −0.63]
subtype.tesla −0.94 [−1.29, −0.59]
−1.4 −1.2 −1 −0.8 −0.6 −0.4
Observed Outcome

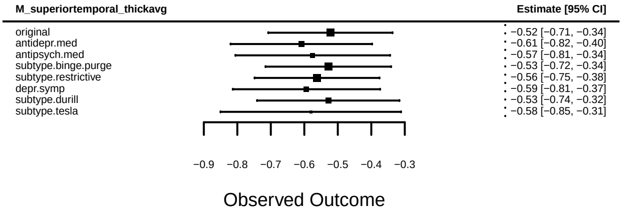
M_superiortemporal_thickavg
Estimate [95% CI]
original −0.52 [−0.71, −0.34]
antidepr.med −0.61 [−0.82, −0.40]
antipsych.med −0.57 [−0.81, −0.34]
subtype.binge.purge −0.53 [−0.72, −0.34]
subtype.restrictive −0.56 [−0.75, −0.38]
depr.symp −0.59 [−0.81, −0.37]
subtype.durill −0.53 [−0.74, −0.32]
subtype.tesla −0.58 [−0.85, −0.31]
−0.9 −0.8 −0.7 −0.6 −0.5 −0.4 −0.3
Observed Outcome

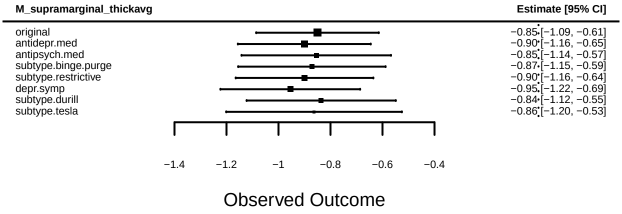
M_supramarginal_thickavg
Estimate [95% CI]
original −0.85 [−1.09, −0.61]
antidepr.med −0.90 [−1.16, −0.65]
antipsych.med −0.85 [−1.14, −0.57]
subtype.binge.purge −0.87 [−1.15, −0.59]
subtype.restrictive −0.90 [−1.16, −0.64]
depr.symp −0.95 [−1.22, −0.69]
subtype.durill −0.84 [−1.12, −0.55]
subtype.tesla −0.86 [−1.20, −0.53]
−1.4 −1.2 −1 −0.8 −0.6 −0.4
Observed Outcome

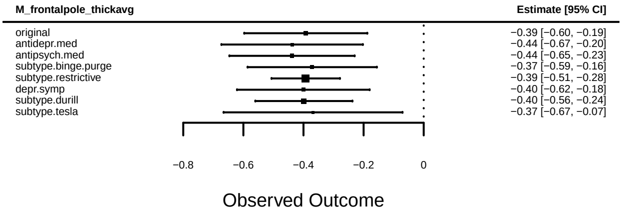
M_frontalpole_thickavg
Estimate [95% CI]
original −0.39 [−0.60, −0.19]
antidepr.med −0.44 [−0.67, −0.20]
antipsych.med −0.44 [−0.65, −0.23]
subtype.binge.purge −0.37 [−0.59, −0.16]
subtype.restrictive −0.39 [−0.51, −0.28]
depr.symp −0.40 [−0.62, −0.18]
subtype.durill −0.40 [−0.56, −0.24]
subtype.tesla −0.37 [−0.67, −0.07]
−0.8 −0.6 −0.4 −0.2 0
Observed Outcome

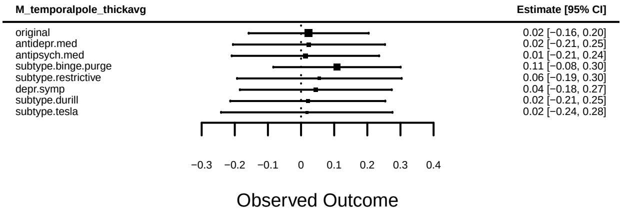
M_temporalpole_thickavg
Estimate [95% CI]
original 0.02 [−0.16, 0.20]
antidepr.med 0.02 [−0.21, 0.25]
antipsych.med 0.01 [−0.21, 0.24]
subtype.binge.purge 0.11 [−0.08, 0.30]
subtype.restrictive 0.06 [−0.19, 0.30]
depr.symp 0.04 [−0.18, 0.27]
subtype.durill 0.02 [−0.21, 0.25]
subtype.tesla 0.02 [−0.24, 0.28]
−0.3 −0.2 −0.1 0 0.1 0.2 0.3 0.4
Observed Outcome

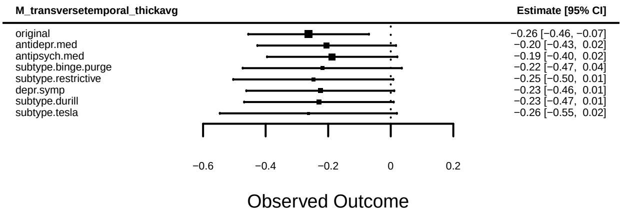
M_transversetemporal_thickavg
Estimate [95% CI]
original −0.26 [−0.46, −0.07]
antidepr.med −0.20 [−0.43, 0.02]
antipsych.med −0.19 [−0.40, 0.02]
subtype.binge.purge −0.22 [−0.47, 0.04]
subtype.restrictive −0.25 [−0.50, 0.01]
depr.symp −0.23 [−0.46, 0.01]
subtype.durill −0.23 [−0.47, 0.01]
subtype.tesla −0.26 [−0.55, 0.02]
−0.6 −0.4 −0.2 0 0.2
Observed Outcome

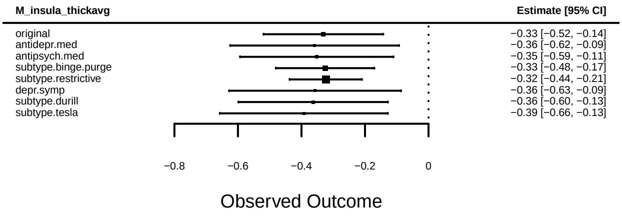
M_insula_thickavg
Estimate [95% CI]
original −0.33 [−0.52, −0.14]
antidepr.med −0.36 [−0.62, −0.09]
antipsych.med −0.35 [−0.59, −0.11]
subtype.binge.purge −0.33 [−0.48, −0.17]
subtype.restrictive −0.32 [−0.44, −0.21]
depr.symp −0.36 [−0.63, −0.09]
subtype.durill −0.36 [−0.60, −0.13]
subtype.tesla −0.39 [−0.66, −0.13]
−0.8 −0.6 −0.4 −0.2 0
Observed Outcome

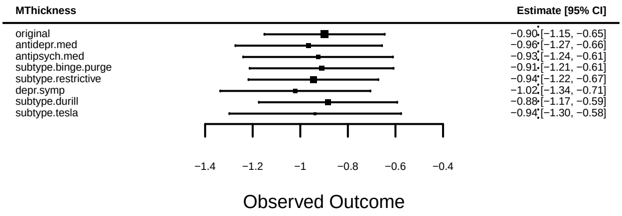
MThickness
Estimate [95% CI]
original −0.90 [−1.15, −0.65]
antidepr.med −0.96 [−1.27, −0.66]
antipsych.med −0.93 [−1.24, −0.61]
subtype.binge.purge −0.91 [−1.21, −0.61]
subtype.restrictive −0.94 [−1.22, −0.67]
depr.symp −1.02 [−1.34, −0.71]
subtype.durill −0.88 [−1.17, −0.59]
subtype.tesla −0.94 [−1.30, −0.58]
−1.4 −1.2 −1 −0.8 −0.6 −0.4
Observed Outcome

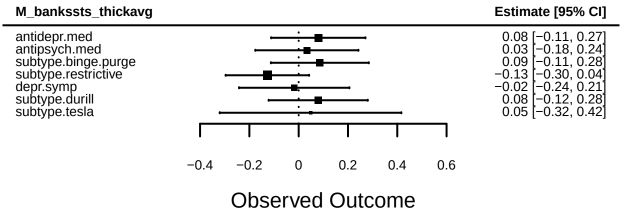
M_bankssts_thickavg
Estimate [95% CI]
antidepr.med 0.08 [−0.11, 0.27]
antipsych.med 0.03 [−0.18, 0.24]
subtype.binge.purge 0.09 [−0.11, 0.28]
subtype.restrictive −0.13 [−0.30, 0.04]
depr.symp −0.02 [−0.24, 0.21]
subtype.durill 0.08 [−0.12, 0.28]
subtype.tesla 0.05 [−0.32, 0.42]
−0.4 −0.2 0 0.2 0.4 0.6
Observed Outcome

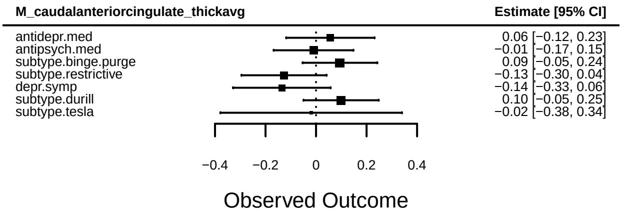
M_caudalanteriorcingulate_thickavg
Estimate [95% CI]
antidepr.med 0.06 [−0.12, 0.23]
antipsych.med −0.01 [−0.17, 0.15]
subtype.binge.purge 0.09 [−0.05, 0.24]
subtype.restrictive −0.13 [−0.30, 0.04]
depr.symp −0.14 [−0.33, 0.06]
subtype.durill 0.10 [−0.05, 0.25]
subtype.tesla −0.02 [−0.38, 0.34]
−0.4 −0.2 0 0.2 0.4
Observed Outcome

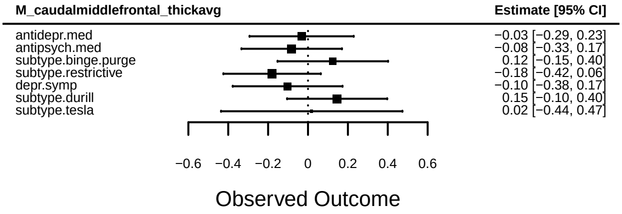
M_caudalmiddlefrontal_thickavg
Estimate [95% CI]
antidepr.med −0.03 [−0.29, 0.23]
antipsych.med −0.08 [−0.33, 0.17]
subtype.binge.purge 0.12 [−0.15, 0.40]
subtype.restrictive −0.18 [−0.42, 0.06]
depr.symp −0.10 [−0.38, 0.17]
subtype.durill 0.15 [−0.10, 0.40]
subtype.tesla 0.02 [−0.44, 0.47]
−0.6 −0.4 −0.2 0 0.2 0.4 0.6
Observed Outcome

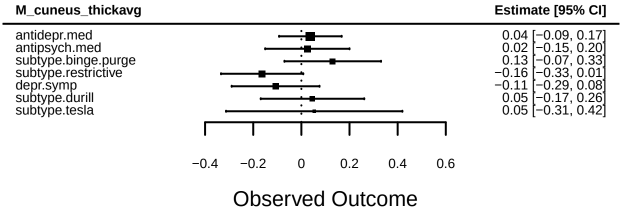
M_cuneus_thickavg
Estimate [95% CI]
antidepr.med 0.04 [−0.09, 0.17]
antipsych.med 0.02 [−0.15, 0.20]
subtype.binge.purge 0.13 [−0.07, 0.33]
subtype.restrictive −0.16 [−0.33, 0.01]
depr.symp −0.11 [−0.29, 0.08]
subtype.durill 0.05 [−0.17, 0.26]
subtype.tesla 0.05 [−0.31, 0.42]
−0.4 −0.2 0 0.2 0.4 0.6
Observed Outcome

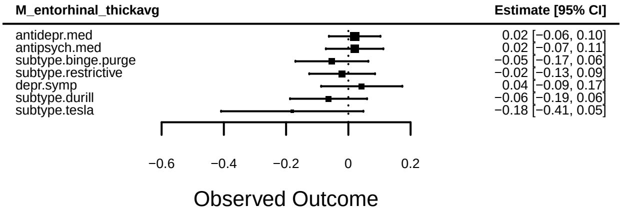
M_entorhinal_thickavg
Estimate [95% CI]
antidepr.med 0.02 [−0.06, 0.10]
antipsych.med 0.02 [−0.07, 0.11]
subtype.binge.purge −0.05 [−0.17, 0.06]
subtype.restrictive −0.02 [−0.13, 0.09]
depr.symp 0.04 [−0.09, 0.17]
subtype.durill −0.06 [−0.19, 0.06]
subtype.tesla −0.18 [−0.41, 0.05]
−0.6 −0.4 −0.2 0 0.2
Observed Outcome

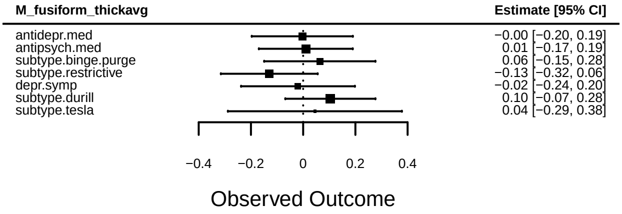
M_fusiform_thickavg
Estimate [95% CI]
antidepr.med −0.00 [−0.20, 0.19]
antipsych.med 0.01 [−0.17, 0.19]
subtype.binge.purge 0.06 [−0.15, 0.28]
subtype.restrictive −0.13 [−0.32, 0.06]
depr.symp −0.02 [−0.24, 0.20]
subtype.durill 0.10 [−0.07, 0.28]
subtype.tesla 0.04 [−0.29, 0.38]
−0.4 −0.2 0 0.2 0.4
Observed Outcome

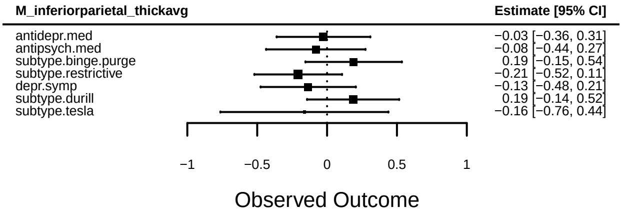
M_inferiorparietal_thickavg
Estimate [95% CI]
antidepr.med −0.03 [−0.36, 0.31]
antipsych.med −0.08 [−0.44, 0.27]
subtype.binge.purge 0.19 [−0.15, 0.54]
subtype.restrictive −0.21 [−0.52, 0.11]
depr.symp −0.13 [−0.48, 0.21]
subtype.durill 0.19 [−0.14, 0.52]
subtype.tesla −0.16 [−0.76, 0.44]
−1 −0.5 0 0.5 1
Observed Outcome

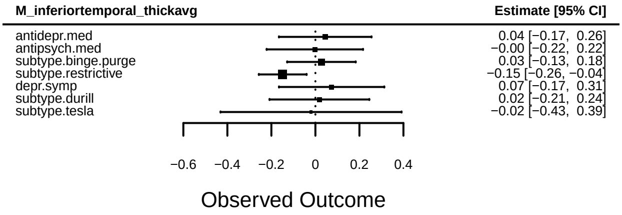
M_inferiortemporal_thickavg
Estimate [95% CI]
antidepr.med 0.04 [−0.17, 0.26]
antipsych.med −0.00 [−0.22, 0.22]
subtype.binge.purge 0.03 [−0.13, 0.18]
subtype.restrictive −0.15 [−0.26, −0.04]
depr.symp 0.07 [−0.17, 0.31]
subtype.durill 0.02 [−0.21, 0.24]
subtype.tesla −0.02 [−0.43, 0.39]
−0.6 −0.4 −0.2 0 0.2 0.4
Observed Outcome

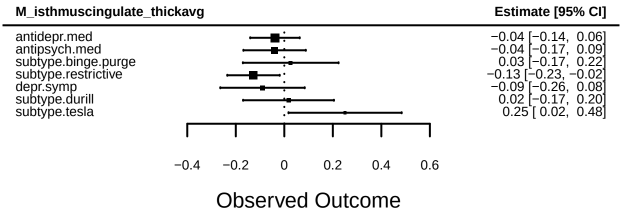
M_isthmuscingulate_thickavg
Estimate [95% CI]
antidepr.med −0.04 [−0.14, 0.06]
antipsych.med −0.04 [−0.17, 0.09]
subtype.binge.purge 0.03 [−0.17, 0.22]
subtype.restrictive −0.13 [−0.23, −0.02]
depr.symp −0.09 [−0.26, 0.08]
subtype.durill 0.02 [−0.17, 0.20]
subtype.tesla 0.25 [ 0.02, 0.48]
−0.4 −0.2 0 0.2 0.4 0.6
Observed Outcome

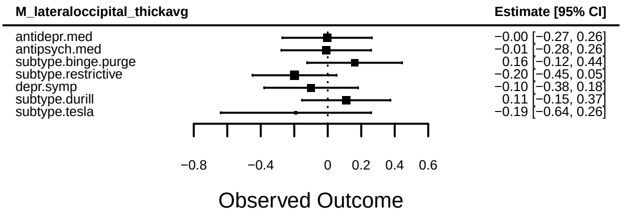
M_lateraloccipital_thickavg
Estimate [95% CI]
antidepr.med −0.00 [−0.27, 0.26]
antipsych.med −0.01 [−0.28, 0.26]
subtype.binge.purge 0.16 [−0.12, 0.44]
subtype.restrictive −0.20 [−0.45, 0.05]
depr.symp −0.10 [−0.38, 0.18]
subtype.durill 0.11 [−0.15, 0.37]
subtype.tesla −0.19 [−0.64, 0.26]
−0.8 −0.4 0 0.2 0.4 0.6
Observed Outcome

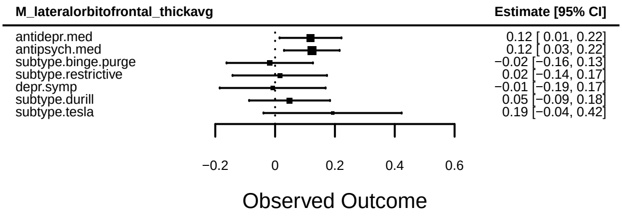
M_lateralorbitofrontal_thickavg
Estimate [95% CI]
antidepr.med 0.12 [ 0.01, 0.22]
antipsych.med 0.12 [ 0.03, 0.22]
subtype.binge.purge −0.02 [−0.16, 0.13]
subtype.restrictive 0.02 [−0.14, 0.17]
depr.symp −0.01 [−0.19, 0.17]
subtype.durill 0.05 [−0.09, 0.18]
subtype.tesla 0.19 [−0.04, 0.42]
−0.2 0 0.2 0.4 0.6
Observed Outcome

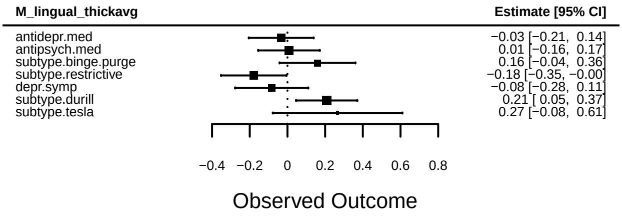
M_lingual_thickavg
Estimate [95% CI]
antidepr.med −0.03 [−0.21, 0.14]
antipsych.med 0.01 [−0.16, 0.17]
subtype.binge.purge 0.16 [−0.04, 0.36]
subtype.restrictive −0.18 [−0.35, −0.00]
depr.symp −0.08 [−0.28, 0.11]
subtype.durill 0.21 [ 0.05, 0.37]
subtype.tesla 0.27 [−0.08, 0.61]
−0.4 −0.2 0 0.2 0.4 0.6 0.8
Observed Outcome

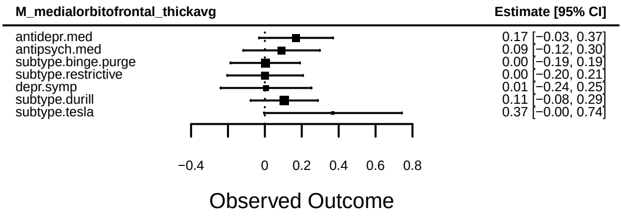
M_medialorbitofrontal_thickavg
Estimate [95% CI]
antidepr.med 0.17 [−0.03, 0.37]
antipsych.med 0.09 [−0.12, 0.30]
subtype.binge.purge 0.00 [−0.19, 0.19]
subtype.restrictive 0.00 [−0.20, 0.21]
depr.symp 0.01 [−0.24, 0.25]
subtype.durill 0.11 [−0.08, 0.29]
subtype.tesla 0.37 [−0.00, 0.74]
−0.4 0 0.2 0.4 0.6 0.8
Observed Outcome

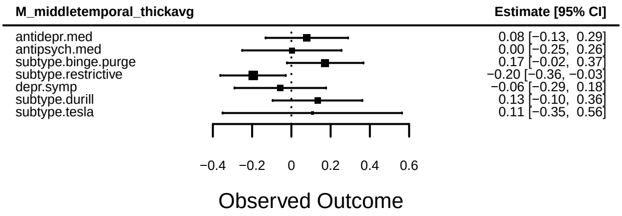
M_middletemporal_thickavg
Estimate [95% CI]
antidepr.med 0.08 [−0.13, 0.29]
antipsych.med 0.00 [−0.25, 0.26]
subtype.binge.purge 0.17 [−0.02, 0.37]
subtype.restrictive −0.20 [−0.36, −0.03]
depr.symp −0.06 [−0.29, 0.18]
subtype.durill 0.13 [−0.10, 0.36]
subtype.tesla 0.11 [−0.35, 0.56]
−0.4 −0.2 0 0.2 0.4 0.6
Observed Outcome

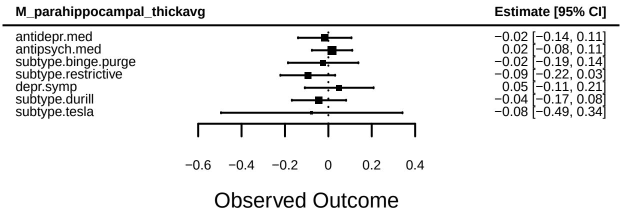
M_parahippocampal_thickavg
Estimate [95% CI]
antidepr.med −0.02 [−0.14, 0.11]
antipsych.med 0.02 [−0.08, 0.11]
subtype.binge.purge −0.02 [−0.19, 0.14]
subtype.restrictive −0.09 [−0.22, 0.03]
depr.symp 0.05 [−0.11, 0.21]
subtype.durill −0.04 [−0.17, 0.08]
subtype.tesla −0.08 [−0.49, 0.34]
−0.6 −0.4 −0.2 0 0.2 0.4
Observed Outcome

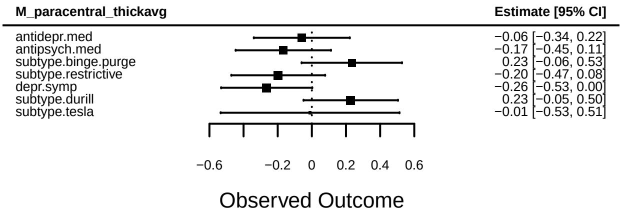
M_paracentral_thickavg
Estimate [95% CI]
antidepr.med −0.06 [−0.34, 0.22]
antipsych.med −0.17 [−0.45, 0.11]
subtype.binge.purge 0.23 [−0.06, 0.53]
subtype.restrictive −0.20 [−0.47, 0.08]
depr.symp −0.26 [−0.53, 0.00]
subtype.durill 0.23 [−0.05, 0.50]
subtype.tesla −0.01 [−0.53, 0.51]
−0.6 −0.2 0 0.2 0.4 0.6
Observed Outcome

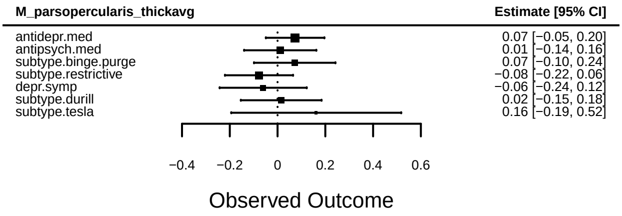
M_parsopercularis_thickavg
Estimate [95% CI]
antidepr.med 0.07 [−0.05, 0.20]
antipsych.med 0.01 [−0.14, 0.16]
subtype.binge.purge 0.07 [−0.10, 0.24]
subtype.restrictive −0.08 [−0.22, 0.06]
depr.symp −0.06 [−0.24, 0.12]
subtype.durill 0.02 [−0.15, 0.18]
subtype.tesla 0.16 [−0.19, 0.52]
−0.4 −0.2 0 0.2 0.4 0.6
Observed Outcome

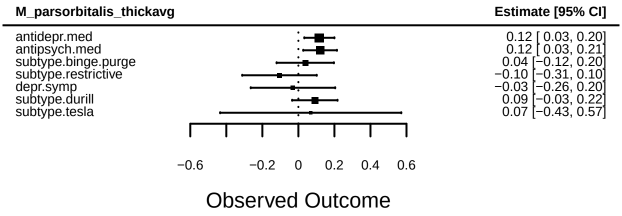
M_parsorbitalis_thickavg
Estimate [95% CI]
antidepr.med 0.12 [ 0.03, 0.20]
antipsych.med 0.12 [ 0.03, 0.21]
subtype.binge.purge 0.04 [−0.12, 0.20]
subtype.restrictive −0.10 [−0.31, 0.10]
depr.symp −0.03 [−0.26, 0.20]
subtype.durill 0.09 [−0.03, 0.22]
subtype.tesla 0.07 [−0.43, 0.57]
−0.6 −0.2 0 0.2 0.4 0.6
Observed Outcome

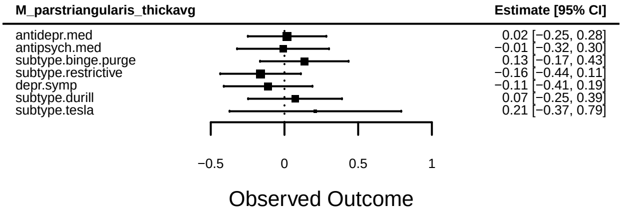
M_parstriangularis_thickavg
Estimate [95% CI]
antidepr.med 0.02 [−0.25, 0.28]
antipsych.med −0.01 [−0.32, 0.30]
subtype.binge.purge 0.13 [−0.17, 0.43]
subtype.restrictive −0.16 [−0.44, 0.11]
depr.symp −0.11 [−0.41, 0.19]
subtype.durill 0.07 [−0.25, 0.39]
subtype.tesla 0.21 [−0.37, 0.79]
−0.5 0 0.5 1
Observed Outcome

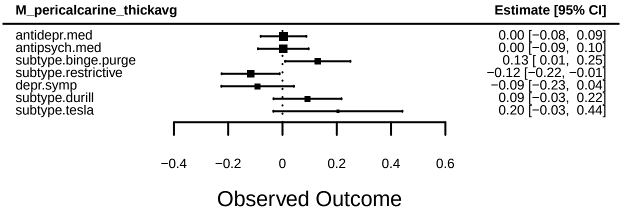
M_pericalcarine_thickavg
Estimate [95% CI]
antidepr.med 0.00 [−0.08, 0.09]
antipsych.med 0.00 [−0.09, 0.10]
subtype.binge.purge 0.13 [ 0.01, 0.25]
subtype.restrictive −0.12 [−0.22, −0.01]
depr.symp −0.09 [−0.23, 0.04]
subtype.durill 0.09 [−0.03, 0.22]
subtype.tesla 0.20 [−0.03, 0.44]
−0.4 −0.2 0 0.2 0.4 0.6
Observed Outcome

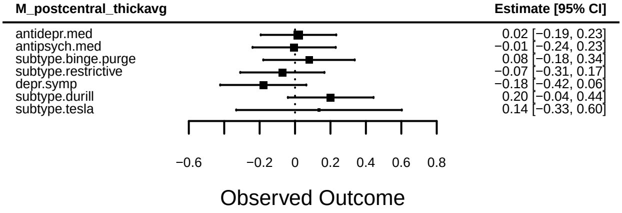
M_postcentral_thickavg
Estimate [95% CI]
antidepr.med 0.02 [−0.19, 0.23]
antipsych.med −0.01 [−0.24, 0.23]
subtype.binge.purge 0.08 [−0.18, 0.34]
subtype.restrictive −0.07 [−0.31, 0.17]
depr.symp −0.18 [−0.42, 0.06]
subtype.durill 0.20 [−0.04, 0.44]
subtype.tesla 0.14 [−0.33, 0.60]
−0.6 −0.2 0 0.2 0.4 0.6 0.8
Observed Outcome

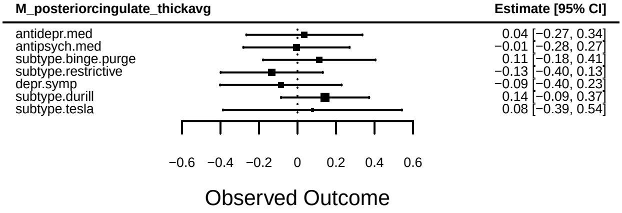
M_posteriorcingulate_thickavg
Estimate [95% CI]
antidepr.med 0.04 [−0.27, 0.34]
antipsych.med −0.01 [−0.28, 0.27]
subtype.binge.purge 0.11 [−0.18, 0.41]
subtype.restrictive −0.13 [−0.40, 0.13]
depr.symp −0.09 [−0.40, 0.23]
subtype.durill 0.14 [−0.09, 0.37]
subtype.tesla 0.08 [−0.39, 0.54]
−0.6 −0.4 −0.2 0 0.2 0.4 0.6
Observed Outcome

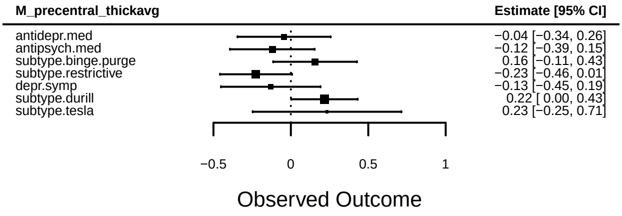
M_precentral_thickavg
Estimate [95% CI]
antidepr.med −0.04 [−0.34, 0.26]
antipsych.med −0.12 [−0.39, 0.15]
subtype.binge.purge 0.16 [−0.11, 0.43]
subtype.restrictive −0.23 [−0.46, 0.01]
depr.symp −0.13 [−0.45, 0.19]
subtype.durill 0.22 [ 0.00, 0.43]
subtype.tesla 0.23 [−0.25, 0.71]
−0.5 0 0.5 1
Observed Outcome

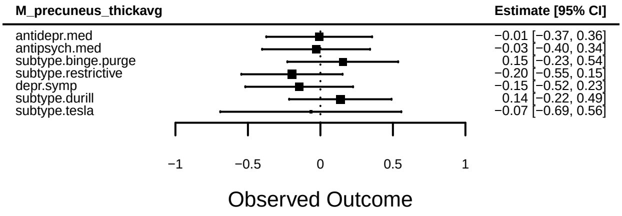
M_precuneus_thickavg
Estimate [95% CI]
antidepr.med −0.01 [−0.37, 0.36]
antipsych.med −0.03 [−0.40, 0.34]
subtype.binge.purge 0.15 [−0.23, 0.54]
subtype.restrictive −0.20 [−0.55, 0.15]
depr.symp −0.15 [−0.52, 0.23]
subtype.durill 0.14 [−0.22, 0.49]
subtype.tesla −0.07 [−0.69, 0.56]
−1 −0.5 0 0.5 1
Observed Outcome

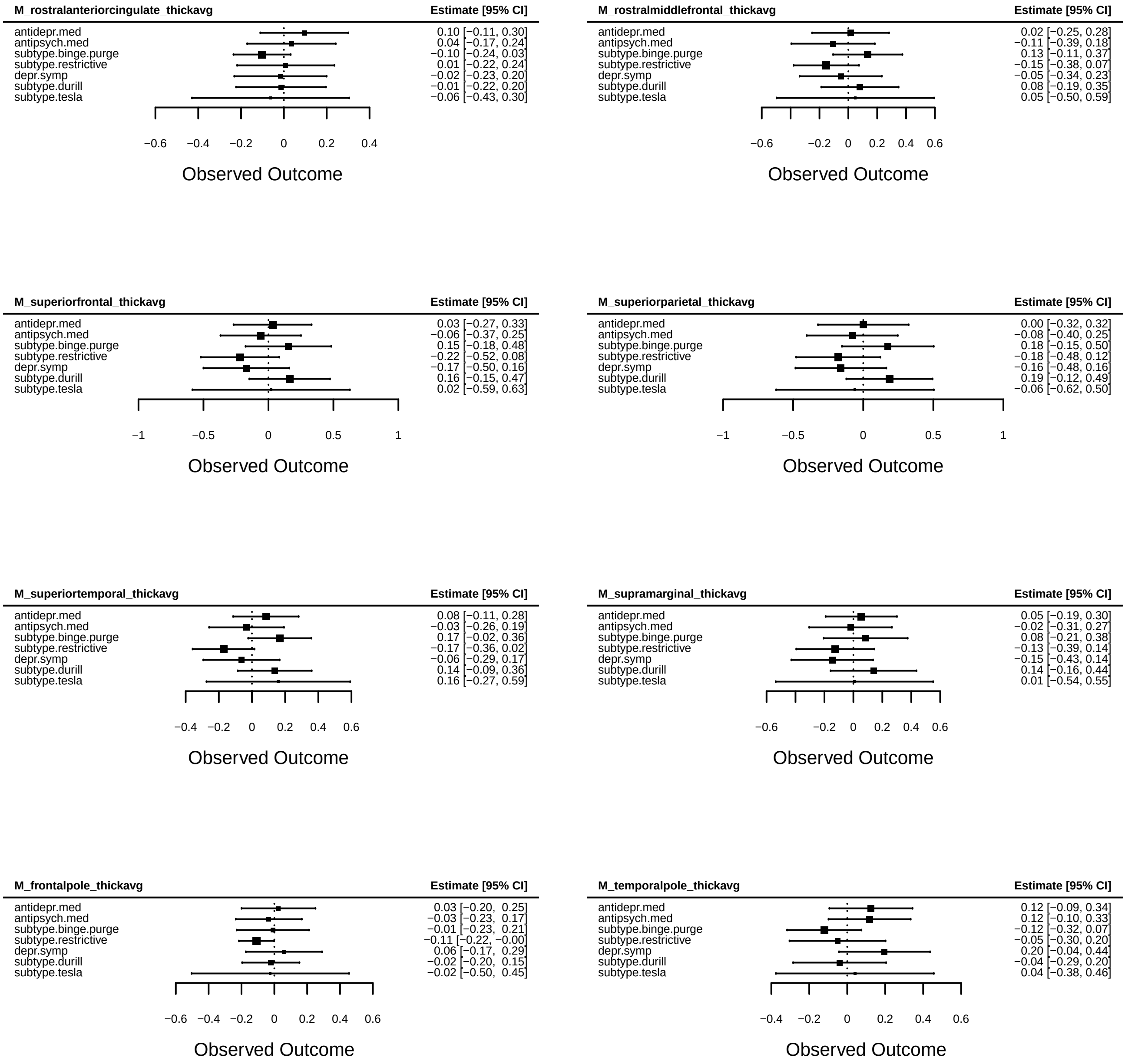
M_rostralanteriorcingulate_thickavg
Estimate [95% CI]
antidepr.med 0.10 [−0.11, 0.30]
antipsych.med 0.04 [−0.17, 0.24]
subtype.binge.purge −0.10 [−0.24, 0.03]
subtype.restrictive 0.01 [−0.22, 0.24]
depr.symp −0.02 [−0.23, 0.20]
subtype.durill −0.01 [−0.22, 0.20]
subtype.tesla −0.06 [−0.43, 0.30]
−0.6 −0.4 −0.2 0 0.2 0.4
Observed Outcome
M_rostralmiddlefrontal_thickavg
Estimate [95% CI]
antidepr.med 0.02 [−0.25, 0.28]
antipsych.med −0.11 [−0.39, 0.18]
subtype.binge.purge 0.13 [−0.11, 0.37]
subtype.restrictive −0.15 [−0.38, 0.07]
depr.symp −0.05 [−0.34, 0.23]
subtype.durill 0.08 [−0.19, 0.35]
subtype.tesla 0.05 [−0.50, 0.59]
−0.6 −0.2 0 0.2 0.4 0.6
Observed Outcome
M_superiorfrontal_thickavg
Estimate [95% CI]
antidepr.med 0.03 [−0.27, 0.33]
antipsych.med −0.06 [−0.37, 0.25]
subtype.binge.purge 0.15 [−0.18, 0.48]
subtype.restrictive −0.22 [−0.52, 0.08]
depr.symp −0.17 [−0.50, 0.16]
subtype.durill 0.16 [−0.15, 0.47]
subtype.tesla 0.02 [−0.59, 0.63]
−1 −0.5 0 0.5 1
Observed Outcome
M_superiorparietal_thickavg
Estimate [95% CI]
antidepr.med 0.00 [−0.32, 0.32]
antipsych.med −0.08 [−0.40, 0.25]
subtype.binge.purge 0.18 [−0.15, 0.50]
subtype.restrictive −0.18 [−0.48, 0.12]
depr.symp −0.16 [−0.48, 0.16]
subtype.durill 0.19 [−0.12, 0.49]
subtype.tesla −0.06 [−0.62, 0.50]
−1 −0.5 0 0.5 1
Observed Outcome
M_superiortemporal_thickavg
Estimate [95% CI]
antidepr.med 0.08 [−0.11, 0.28]
antipsych.med −0.03 [−0.26, 0.19]
subtype.binge.purge 0.17 [−0.02, 0.36]
subtype.restrictive −0.17 [−0.36, 0.02]
depr.symp −0.06 [−0.29, 0.17]
subtype.durill 0.14 [−0.09, 0.36]
subtype.tesla 0.16 [−0.27, 0.59]
−0.4 −0.2 0 0.2 0.4 0.6
Observed Outcome
M_supramarginal_thickavg
Estimate [95% CI]
antidepr.med 0.05 [−0.19, 0.30]
antipsych.med −0.02 [−0.31, 0.27]
subtype.binge.purge 0.08 [−0.21, 0.38]
subtype.restrictive −0.13 [−0.39, 0.14]
depr.symp −0.15 [−0.43, 0.14]
subtype.durill 0.14 [−0.16, 0.44]
subtype.tesla 0.01 [−0.54, 0.55]
−0.6 −0.2 0 0.2 0.4 0.6
Observed Outcome
M_frontalpole_thickavg
Estimate [95% CI]
antidepr.med 0.03 [−0.20, 0.25]
antipsych.med −0.03 [−0.23, 0.17]
subtype.binge.purge −0.01 [−0.23, 0.21]
subtype.restrictive −0.11 [−0.22, −0.00]
depr.symp 0.06 [−0.17, 0.29]
subtype.durill −0.02 [−0.20, 0.15]
subtype.tesla −0.02 [−0.50, 0.45]
−0.6 −0.4 −0.2 0 0.2 0.4 0.6
Observed Outcome
M_temporalpole_thickavg
Estimate [95% CI]
antidepr.med 0.12 [−0.09, 0.34]
antipsych.med 0.12 [−0.10, 0.33]
subtype.binge.purge −0.12 [−0.32, 0.07]
subtype.restrictive −0.05 [−0.30, 0.20]
depr.symp 0.20 [−0.04, 0.44]
subtype.durill −0.04 [−0.29, 0.20]
subtype.tesla 0.04 [−0.38, 0.46]
−0.4 −0.2 0 0.2 0.4 0.6
Observed Outcome

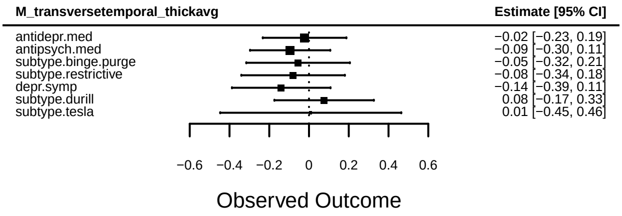
M_transversetemporal_thickavg
Estimate [95% CI]
antidepr.med
antipsych.med
subtype.binge.purge
subtype.restrictive
depr.symp
subtype.durill
subtype.tesla
−0.02 [−0.23, 0.19]
−0.09 [−0.30, 0.11]
−0.05 [−0.32, 0.21]
−0.08 [−0.34, 0.18]
−0.14 [−0.39, 0.11]
0.08 [−0.17, 0.33]
0.01 [−0.45, 0.46]
−0.6
−0.4
−0.2
0
0.2
0.4
0.6
Observed Outcome

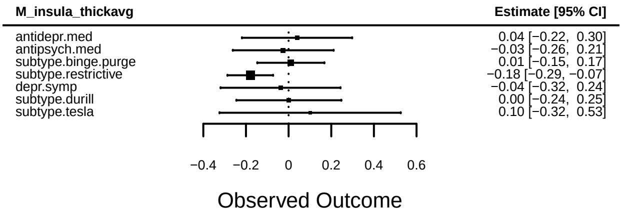
M_insula_thickavg
Estimate [95% CI]
antidepr.med
antipsych.med
subtype.binge.purge
subtype.restrictive
depr.symp
subtype.durill
subtype.tesla
0.04 [−0.22, 0.30]
−0.03 [−0.26, 0.21]
0.01 [−0.15, 0.17]
−0.18 [−0.29, −0.07]
−0.04 [−0.32, 0.24]
0.00 [−0.24, 0.25]
0.10 [−0.32, 0.53]
−0.4
−0.2
0
0.2
0.4
0.6
Observed Outcome

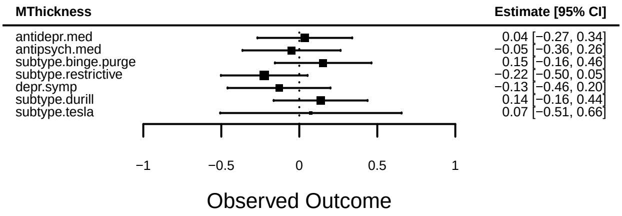
MThickness
Estimate [95% CI]
antidepr.med
antipsych.med
subtype.binge.purge
subtype.restrictive
depr.symp
subtype.durill
subtype.tesla
0.04 [−0.27, 0.34]
−0.05 [−0.36, 0.26]
0.15 [−0.16, 0.46]
−0.22 [−0.50, 0.05]
−0.13 [−0.46, 0.20]
0.14 [−0.16, 0.44]
0.07 [−0.51, 0.66]
−1
−0.5
0
0.5
1
Observed Outcome
